# Psychometric archetypes reveal biological signatures of vulnerability, resilience, and future mental health

**DOI:** 10.1101/2024.09.01.24312906

**Authors:** Niels Mørch, Andrés Barrena Calderón, Timo Lehmann Kvamme, Julie G. Donskov, Ditte Kim Secher, Júlia Díaz-i-Calvete, Dimitrios Pediotidis-Maniatis, Blanka Zana, Simon Durand, Bianka Rumi, Jovana Bjekic, Maro Machizawa, Makiko Yamada, Filip Ottosson, Jonas Bybjerg-Grauholm, Madeleine Ernst, Naveed Ur Rehman, Jakob Grove, Anders D. Børglum, Kristian Sandberg, Per Qvist

**Author notes:** These authors contributed equally to this work (Niels Mørch, Andrés Barrena Calderón).

## Abstract

**Background:** Mental health comprises emotional, psychological, and social dimensions, extending beyond the mere absence of illness. Shaped by a complex interplay of hereditary factors and life experiences, mental health can deteriorate into clinical conditions necessitating intervention. However, ambiguity between pathological and non-pathological states underscores the need for a dimensional approach to early risk detection and stratified psychiatry.

**Methods:** We analyzed multimodal data from approximately 300 young adults, including psychometric assessments, structural brain imaging, genomic data, and fasting-state plasma metabolomics. Using a psychometry-based soft-clustering approach (archetyping), we stratified participants based on cognitive, emotional, and behavioral traits. We evaluated associations between archetypes, biological features, and mental health outcomes both cross-sectionally and at five-year follow-up.

**Results:** We identified five psychometric archetypes, capturing diverse psychological profiles that span a continuum from vulnerability to resilience. Archetypes at the vulnerable end, which were marked by emotional dysregulation and high neuroticism, were associated with elevated polygenic risk for psychiatric disorders, altered cortical structures in emotion-related regions, and metabolomic profiles previously linked to psychopathology. By contrast, resilient archetypes were characterized by emotional stability and adaptive functioning. Archetype scores were prospectively associated with symptom burden, and models guided by archetype-associated biological features outperformed agnostic models in predicting clinical outcomes, supporting their clinical relevance.

**Conclusions:** This study demonstrates that data-driven psychometric archetypes reflect biologically grounded variation in mental health and can inform prospective risk stratification. This approach offers a framework for understanding mental health heterogeneity and holds promise for advancing early screening and targeted interventions in the young population.

## INTRODUCTION

Mental health is a dynamic and multifaceted trait that extends beyond the mere absence of mental illness (MDx). It encompasses emotional, psychological, and social dimensions, affecting how individuals perceive, feel, and navigate life’s challenges and rewards. Shaped by both genetic predispositions and life experiences^1^, mental health can deteriorate into clinical conditions necessitating intervention. However, the boundary between pathological and non-pathological states is often ambiguous, and overlapping clinical profiles make diagnostic categories difficult to define^2^. Compelling evidence for interconnected etiologies exists, as genetic risk variants are typically nonspecifically associated with a range of MDx^3–5^, and many patients transition between diagnostic categories or develop mental co-morbidities over the course of life. Collectively, there is compelling evidence^6,7^, and an emerging consensus among clinicians^8^, that MDx exists on a continuum that includes mental health-related traits descriptive of the general population^9,10^, challenging the validity of their current categorical diagnostic classification. A dimensional approach aimed at identifying prodromal, intermediate, and transdiagnostic mental health risk signatures is therefore essential for advancing stratified and precision psychiatry^11,12^.

Despite growing consensus around dimensional models of mental health, much psychiatric research remains anchored in traditional case-control designs. While informative, such studies typically focus on diagnostic extremes and exclude large segments of the population experiencing mixed symptoms or subclinical distress^13^. This narrow focus limits our understanding of the full spectrum of mental health variation and the underlying mechanisms that span diagnostic boundaries^14^. In contrast, transdiagnostic approaches consider broader psychometric profiles and acknowledge the phenotypic and etiological overlap between diagnostic categories, offering a more nuanced framework for identifying the processes that drive vulnerability and resilience across the mental health continuum^15^.

Building on this dimensional perspective, recent years have witnessed major advances in elucidating the genetic and biological underpinnings of mental health, paving the way for integrated biomarker research that complements psychometric stratification. In particular, genome-wide association studies (GWASs) have provided insights into the genetic architecture of both mental health-related traits and psychiatric conditions^16–20^. As ascertainment strategies have shifted toward more comprehensively phenotyped cohorts^21^, the pleiotropic effects and broader phenotypic correlates of genetic risk factors have begun to emerge^22–25^. It is now well established that genetic predisposition to mental disorders intersects with genetic variation associated with diverse behavioral, neuroimaging, and mental health-related traits in the general population^26,27^ (e.g., personality^6^, cognitive ability^28^, impulsivity^29^, sleep patterns^30^, and musicality^31^). Although polygenic scores (PGS) rarely explain more than 5% of the variance in mental health-related traits, leveraging the shared genetic architecture between mental disorders and related traits can substantially improve prediction of individual risk, recurrence, and psychiatric comorbidity^32,33^. Nevertheless, as clinical manifestation is attributable to a combination of heritable, biological, and environmental factors^16^, genetic risk alone will always only provide limited insight into an individual’s current mental health status, clinical profile, and trajectory^34^. Peripheral biomarkers may offer a promising solution by capturing both transient and persistent signatures of mental health and aiding in the prediction of mental health outcomes^35,36^. In this context, blood-based biomarkers have been identified that reflect dynamic changes in the course of mental illness^36,37^, and non-invasive multimodal neuroimaging has revealed subtle anatomical and functional brain changes even at prodromal stages^38^. Critically, combining polygenic scores with other biomarkers has been shown to explain more variance in complex traits than using PGSs alone^39,40^.

Here, we utilize a novel psychometry-based archetyping approach in a representative and deeply phenotyped sample of young adults to identify biologically grounded mental health profiles. By integrating psychometric data with genomic, neuroimaging, and blood-based metabolomic features, we characterize archetype-specific biological signatures and evaluate their relevance to participants’ mental health status. Finally, we assess whether these profiles can prospectively predict psychiatric symptom burden and future mental health outcomes over a five-year period, providing a proof-of-concept framework for dimensional, biologically informed risk stratification.

## MATERIALS AND METHODS

### Study population and procedures

The data used in this study were collected as part of the EU COST Action CA18106 and originate from Aarhus University, Denmark. These data are part of a larger dataset focused on the Neural Architecture of Consciousness and have also been used in other studies with different aims^41–43^. Some methodological descriptions have been adapted from these previous publications.

Participants were recruited from the Center for Functionally Integrative Neuroscience’s participant pool at Aarhus University and through local advertisements. Inclusion criteria were: (a) anatomically normal brain (no known abnormalities, brain damage, or brain surgery); (b) age 18-50 years; (c) good physical health; (d) normal or corrected-to-normal vision; and (e) normal hearing. Exclusion criteria were: (a) MRI contraindications; (b) use of neuropharmacological or other medications affecting neural states; (c) body size incompatible with MRI scanning; (d) pregnancy; and (e) dermatological conditions interfering with MRI procedures. Participants received monetary compensation. A total of 351 (210 female) participants were included (**Table S1** for details). The study was approved by the local ethics committee (De Videnskabsetiske Komitéer for Region Midtjylland, Denmark) and conducted in accordance with the Declaration of Helsinki and all other relevant ethical guidelines and regulations.

At enrollment, participants completed the NEO-PI-3 personality assessment online, as well as an extensive questionnaire covering psychological and behavioral domains, including awareness, mindfulness, perception, cognition, emotional regulation, anhedonia, depression, sleep patterns, impulsiveness, and stress (see **Table S2** for the full list). Participants also provided basic demographic information, including employment status, education level and field of study, civil status, and personal and family history of mental health conditions.

Approximately one to two weeks after completing the online sessions, participants underwent a one-hour MRI session comprising diffusion-weighted imaging (DWI), multi-parameter mapping (MPM), and resting-state functional MRI (fMRI). In a separate session, typically within two weeks of the MRI, participants completed an IQ assessment (WAIS-IV). Buccal epithelial cells were collected from 304 participants using SK-1S Isohelix™ buccal swabs, and DNA was purified using the Isohelix™ Buccal-Prep Plus DNA Isolation Kit (YouDoBio, Rødovre, Denmark). Fasting-state blood plasma collected from 198 participants within two weeks of the MRI session. DNA and plasma samples were stored at −80°C until further processing.

### Psychometry-based archetyping

Psychometric archetypes were identified using an unsupervised soft-clustering approach implemented with the R package ‘archetypes’^44^ and its ‘robustArchetypes’^45^ function. Archetype analysis detects extreme profiles within a multivariate dataset and represents each individual as a convex combination of these extremes, thereby capturing the spectrum of phenotypic variation. To determine the optimal number of archetypes (*k)*, the minimized residual sum of squares (RSS), a standard metric in archetypal analysis, was evaluated across varying numbers of archetypes using a scree plot analysis. The Calinski-Harabasz Index (CHI) was then applied to determine the most suitable cutoff for distinguishing individuals with clear categorical archetype membership. Participants who did not meet this requirement were classified as having a ‘mixed’ profile. Cluster validity was assessed *post hoc* using a Silhouette analysis based on a standard distance matrix, implemented with the R packages ‘factoextra’^46^ and ‘cluster’^47^ (**Figure S2-3**).

### Genotyping, relatedness pruning, and removal of ancestry outliers

Genotyping was performed at Statens Serum Institut (SSI, Copenhagen, Denmark) using the Global Screening Array v2 with a multi-disease drop-in (Illumina, San Diego, California) according to the manufacturer’s instructions. Genotype calling was performed using GenTrain V3. Quality control (QC) was conducted at the marker level. Variants were retained if they met the following criteria: call rate ≥ 0.98, missing difference ≤ 0.02 between cases and controls, minor allele frequency (MAF) ≥ 0.005, and Hardy-Weinberg equilibrium (HWE) p ≥ 1×10⁻⁷ (See https://sites.google.com/a/broadinstitute.org/ricopili/preimputation-qc for further details).

Phasing of the merged genotype set performed using EAGLE (v2.4.1)^48^ and imputed with the EUR population of the 1000g-phase-3-v5 (hg19) reference panel using Minimac4 (v1.0.0)^49^. Imputed SNPs with an imputation quality score (R^2^) <0.3 were excluded.

Principal components were calculated using LDAK, and 21 ancestry outliers were identified by visual inspection of the PCA plot and removed (**Figure S4**). Principal components were then recalculated on the remaining individuals and used as covariates in subsequent analyses.

### Polygenic scores, genetic prediction, and genetic archetypes

Polygenic scores (PGSs) were derived from a comprehensive library of publicly available GWAS summary statistics for traits broadly related to mental health, including behavioral, cognitive, and neuroimaging phenotypes^50–95^ (**Table S3).** Summary statistics files were harmonized with the software program process_sumstats^96^, and PGSs were computed using MegaPRS^97^, which has demonstrated strong predictive performance for psychiatric traits compared to other advanced PGS methods^98^. MegaPRS was run with standard settings recommended by the authors (https://dougspeed.com/megaprs/) using the BLD-LDAK heritability model and Elastic-SS to construct the prediction model. In the heritability model, the α parameter was set to 0.25, and high linkage disequilibrium regions were excluded. Model parameters were selected using pseudo cross-validation. The 1000 Genomes from the non-Finnish European subpopulation were used as a reference to estimate SNP-SNP correlations. Only SNPs with a minor allele frequency ≥1% and an imputation score ≥0.9 (when available) were included for PGS construction.

### Magnetic resonance imaging (MRI)

Imaging was performed on a Siemens Magnetom Prisma-fit 3T MRI scanner. The procedure commenced with preliminary scouting scans. This was followed by two sequences of resting-state fMRI, lasting 12 and 6 minutes, respectively. The session also included quantitative multi-parameter mapping (approximately 20 minutes) to facilitate the synthetic generation of T1-weighted images. Additionally, high-angular resolution diffusion imaging (HARDI) was conducted over approximately 10 minutes, all within a single session lasting about one hour.

For every participant, 1500 functional volumes were acquired, with a repetition time (TR) of 700 ms and an echo time (TE) of 30 ms. The parameters set were: a voxel size of 2.5 mm^3^, a field of view (FOV) of 200 mm, and a flip angle of 53°.

The MPM protocol was implemented using the Siemens vendor-provided 3D multi-echo spoiled gradient echo (FLASH) sequences, with separate acquisitions optimized for magnetization transfer (MT), proton density (PD), and T1 contrast weighting. TRs were 18 ms for the PD- and T1-weighted scans and 37 ms for the MT-weighted scan. All sequences used six equidistant echoes with TE ranging from 2.46 to 14.76 ms. Flip angles were 6° (MT), 4° (PD), and 25° (T1). All images were acquired at 1 mm isotropic resolution, with a field of view of 224 × 256 × 176 mm, and with GRAPPA parallel imaging acceleration (factor 2) in the anterior–posterior (AP) phase-encoding direction. To estimate the RF receive field sensitivity (B1⁻), additional low-resolution 3D spoiled gradient echo volumes were acquired using both the head and body coils. These volumes had a resolution of 4 mm³ isotropic, TE ∼2 ms, flip angle of 6°, and no parallel imaging. Volume pairs (head/body coil) were acquired prior to each of the MTw, PDw, and T1w scans to compute relative B1⁻ maps.

The HARDI sequence incorporated multiple diffusion directions: 75 at b = 2500 s/mm^2^, 60 at b = 1500 s/mm^2^, 21 at b = 1200 s/mm^2^, 30 at b = 1000 s/mm^2^, 15 at b = 700 s/mm^2^, and 10 at b = 5 s/mm^2^. These different b-shells were acquired in a single series with a flip angle of 90°, a TR/TE of 2850/71 ms, a voxel size of 2 mm^3^, a matrix size of 100 x 100, and 84 slices in total. The primary phase-encoding direction was from anterior to posterior (AP), with an additional acquisition in the opposite phase-encoding direction (PA) at b = 0 s/mm^2^ for EPI distortion correction.

To create synthetic T1-weighted images, high-resolution longitudinal relaxation rate (R1) and effective proton density (PD) maps were utilized and obtained through the MPM sequence protocol^99^. Initially, these maps underwent thresholding to align with FreeSurfer’s required units. The R1 map was transformed into a T1 map by inverting its values and applying a zero threshold, followed by a multiplication of 1000. Similarly, the PD map was zero-thresholded and scaled up by a factor of 100. These adjustments were carried out using FSL maths commands. The FreeSurfer’s "mri_synthesize" command was then employed to generate a synthetic FLASH image, using the modified T1 (derived from the adjusted R1 map) and PD maps. Optional arguments were used to enhance the contrast between gray and white matter, with parameters set at 20, 30, and 2.5. In the final step, the synthetic T1-weighted image was reduced by a quarter to meet FreeSurfer’s expected scale (see Keller et al.^100^ and Kvamme et al.^41^ for further details). The cerebral cortex was mapped onto the Desikan-Killiany atlas. Of note, structural brain metrics were not corrected for total brain volume or intracranial volume.

### Metabolomics

Plasma samples (100 μL each) were randomly distributed across three 96-well plates (batches). Before sample preparation, a batch of plasma was set aside as external control (EC) samples and stored at −80 °C. These EC samples, along with plate-specific pools of all plasma samples, were used for quality control purposes (**Supplementary Material** under the *Supplementary Methods and Materials* section). Sample preparation involved extraction in cold methanol/acetonitrile (50/50), incubation for 15 minutes, freezing, and centrifuging. The resulting supernatant was evaporated under nitrogen and reconstituted in 95% solvent A (99.8% water, 0.2% formic acid) and 5% solvent B (49.9% methanol, 49.9% acetonitrile, 0.2% formic acid). Mass spectrometry analysis was performed using a timsTOF Pro mass spectrometer coupled to a UHPLC Elute LC system, Bruker Daltonics (Billerica, MA, US). The analytical separation was performed on an Acquity HSS T3 (100 Å, 2.1 mm x 100 mm, 1.8 µm) column (Waters, Milford, MA, US). The analysis started with 99% solvent A for 1.5 min; thereafter, a linear gradient to 95% solvent B for 8.5 min followed by an isocratic condition at 95% mobile phase B for 2.5 min before going back to 99% mobile phase A and equilibration for 2.4 min. Metabolomics preprocessing was done using the Ion Identity Network workflow in MZmine^101–103^(version 3.3.9). Before statistical analysis, metabolite features present in less than 25% of the samples were removed, and features present in fewer than 75% were treated as binary variables (present or absent). This resulted in a final dataset with a total of 1076 metabolite features measured, among which 433 features were continuous and 643 were binary variables. Missing values for metabolite features with continuous measurements were further subjected to imputation using missForest^104^, and subsequent batch correction was performed by centering and univariate scaling each metabolite per batch. Annotation of metabolite features was performed using mass spectral molecular networking through the GNPS Platform^105,106^ and, *in silico* annotation using Sirius+CSI:FingerID^107^ and, deep neural networks in CANOPUS^108^. Detailed descriptions of sample preparation, mass spectrometry analysis, preprocessing, annotation, and quality control procedures can be found in the **Supplementary Material** under the *Supplementary Methods and Materials* section.

### Integration of multimodal features with archetypes using correlation analysis

Integration of polygenic scores (PGSs), plasma metabolomics, and MRI features with archetype scores was performed using a correlation-based visualization strategy. This approach was used to represent cross-modal associations, without implying formal network analysis.

For each data modality, the five features most strongly associated with A1 and A5 archetype scores were selected based on absolute Spearman correlation coefficients. Features were retained only if they showed sufficient variance and data coverage.

Pairwise Spearman correlations were then calculated between (1) archetype scores and features within each modality and (2) features across different modalities. P-values were adjusted for multiple testing using the Benjamini–Hochberg false discovery rate (FDR) procedure (α = 0.05).

Visualization parameters were standardized: edges were colored by correlation direction (positive or negative), edge thickness was proportional to correlation strength, and node size reflected the number of connections.

### Participant Recontact and Data Collection

Approximately five years after baseline participation, all individuals originally enrolled in the study were invited to take part in a follow-up assessment. Participants were recontacted via the secure Danish digital mailbox system (e-Boks), and questionnaires were administered using the SurveyXact platform (Rambøll, Denmark). The follow-up included the Mental Health Questionnaire 2 (MHQ2), a self-report instrument developed for large-scale mental health surveillance in the UK Biobank cohort^109^. MHQ2 covers both current and lifetime psychiatric symptoms, diagnoses, and treatment experiences though symptom severity scales (Depression (PHQ-9); Anxiety (GAD-7); Alcohol use (AUDIT); Loneliness (UCLA Loneliness Scale); Resilience (Brief Resilience Scale); as well as Binary diagnostic and experience-based outcomes (including: Self-reported psychiatric diagnoses; Trauma exposure; Self-harm and suicidality; Bipolar spectrum and psychosis-like experiences). Symptom scale scores were calculated according to published scoring algorithms. Binary mental health outcomes were derived based on algorithmic case definitions and self-report items included in MHQ2.

### Machine learning

We aimed to build predictive models for psychiatric diagnosis at follow-up and to test whether archetype-guided feature selection improves performance compared to agnostic approaches. Given the high-dimensional but relatively small sample size, we employed an automated machine learning platform (JADBio), which systematically optimizes preprocessing, feature selection, and model training^110,111^. Preprocessing steps include normalization, mean or mode imputation for missing values, and removal of constant or near-constant features. All preprocessing and feature selection steps are nested within the cross-validation process to prevent data leakage and overfitting.

Two feature selection strategies were applied. In the first feature selection strategy, models were trained using predefined feature sets based on pairwise Spearman correlations with the outcome. Specifically, an archetype-guided feature selection was employed, where the five features from a specific modality most strongly associated with A1 or A1 and A5 archetype scores were selected. These predefined sets were used directly for model training, without JADBio’s internal feature selection. In the second feature selection strategy, JADBio’s feature selection was used. It applies and compares two different methods of feature selection, namely Lasso^112^ and Statistically Equivalent Signatures (SES)^113^. Both Lasso and SES identify minimal, non-redundant sets of features with strong predictive value, as well as univariate feature selection with Benjamini-Hochberg correction. For each strategy, the models were trained on three different data modality combinations: one using psychometric data only, one using biological data only (PGSs, brain imaging measures, and plasma metabolites), and one combining both.

Classification algorithms included Ridge Logistic Regression, Decision Trees, Random Forests, and Support Vector Machines (SVMs) with linear, polynomial, and Gaussian kernels.

Model performance was estimated using Bootstrap Bias-Corrected Cross-Validation (BBC-CV)^111,114^, which accounts for the anti-conservative bias introduced by testing multiple configurations, conceptually equivalent to adjusting p-values in hypothesis testing. To ensure robustness, repeated 10-fold cross-validation was performed, and the tuning effort was set to “extensive”. Model performance was evaluated by maximizing the area under the precision-recall curve (PR AUC), as our dataset was skewed due to class imbalance.

To estimate the contribution of individual features, *JADBio* quantified the change in model performance when each feature was excluded, providing an interpretable measure of feature importance. The final outputs included a trained predictive model, a minimal feature set (biosignature), and confidence intervals for out-of-sample performance estimates.

### Statistical analysis

While the machine learning framework evaluated the predictive value of psychometric and biological features for future psychiatric outcomes, statistical analyses were conducted to establish direct associations between archetype scores and individual biological measures. Ordinary least squares regression was applied to test associations between archetype scores and plasma metabolites, PGSs, or brain imaging metrics. Prior to modeling, each feature X(ᵢ, ⱼ) (for participant *i* and feature *j*) and age were standardized to mean = 0 and SD = 1; gender was encoded as a binary variable. For each archetype score *k* and each feature *j*, we fitted:

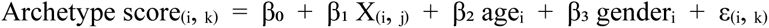

Models were run separately for PGSs, metabolite abundances, and brain imaging metrics, including volumetric measures, cortical curvatures, cortical thickness, vertex count, folding measures, and cortical surface area.

From each model, the regression coefficient β₁, its standard error, the 95% confidence interval, and the raw p-value testing the null hypothesis H₀: β₁ = 0 were extracted. Within each data modality (PGSs, metabolomics, brain imaging metrics), raw p-values were corrected for multiple testing using the Benjamini-Hochberg FDR to yield q-values. All tests were two-sided, and significance was set at FDR q < 0.05.

All analyses of the CA18106 sample were performed at the secured national GenomeDK^115^ high-performance computing cluster in Denmark. Relevant statistical analyses and plotting were performed in the R Statistical Computing environment v4.3.1 (https://www.r-project.org/).

## RESULTS

### Deep psychometric profiling reveals five distinct archetypes

Psychometric data were collected from 351 research volunteers (∼60% female) sampled from the young Danish population (mean age 24.4 years; **Figure S1**). The cohort predominantly consisted of university and college students (approximately 80%), with smaller proportions employed full-time (approximately 8%) or unemployed (approximately 8%). Approximately 38% of participants reported being single (**Table S1)**. Notably, approximately 13% of participants reported a history of mental disorders, including about 6% with multiple diagnoses. The most prevalent conditions were depression (MDD), anxiety and phobia, attention deficit hyperactivity disorder (ADHD), and stress-related disorders (**Figure S5**), thus roughly reflecting the Danish population average for this age group^116^.

Recognizing the continuous nature of mental health-related traits, we employed a soft-clustering method, archetypal analysis^117^, on 258 individuals with complete psychometric data to stratify the sample based on their broadly characterized cognitive, emotional, and behavioral patterns (**Table S2**). Archetypal analysis is a data-driven clustering method that identifies extreme, representative profiles (archetypes) that, within our dataset, define the outer boundaries of variation in psychological traits. This dimensional framework enables a more flexible and realistic representation of mental health diversity, particularly in community-based populations where overlapping symptom patterns are common. By identifying extreme psychometric profiles on individual scales and positioning individuals within the phenotypic spectrum as convex combinations of these extremes, our analysis revealed five stable psychometric archetypes (A1-5) (**Figure 1A** and **Figure S2-3**). Each participant was assigned quantitative archetype scores, reflecting their degree of membership in each archetype. These scores can be interpreted as the probability of belonging to a specific archetype and provide a dimensional view of mental health-related traits. In addition, to facilitate group-based comparisons and to more directly characterize individuals at the extreme of the archetype spectrum, we defined categorical archetypes as those individuals with a membership greater than 0.5 to any archetype. Most individuals (55%) were mainly affiliated with one dominant archetype, whereas 45% were located in the middle of the phenotype distribution with moderate contributions from two or more archetypes (mixed group) (**Figure 1B**). Although a gender bias was seen for individuals with the categorical A4 archetype, differences in gender and age did not otherwise appear to define the archetypes (**Figure S6**). By focusing on categorical archetypes, we highlighted the psychometric features that most clearly characterize each archetype. Unsurprisingly, differences in facets of personality were a significant contributing factor (**Figure 1C** and **Table S4**). Specifically, A1 (22 members) was characterized by high neuroticism as well as low extraversion and conscientiousness scores; A2 (24 members) by high openness score; A3 (26 members) by low agreeableness scores; A4 (39 members) by low openness scores; and A5 (30 members), being the antithesis of A1, with low neuroticism scores as well as high extraversion and conscientiousness scores (**Figure 1C** and **Table S4**). Variation in other psychometrics that were to some degree correlated with personality measures (**Figure S7** - e.g., perception, cognition, impulsiveness, perceived stress, sleep patterns, mindfulness, emotional regulation, hedonic tone, and interoceptive awareness) further significantly shaped the categorical archetypes (**Figure 1C** and **Table S4**). In alignment with the marked differences in neuroticism score between archetypes, individuals with a dominant A1 profile scored significantly higher than the rest of the sample in the commonly applied depression screening tool^118^, the CES-D depression scale (**Figure 1C-D**; Mann-Whitney U test: p = 0.012). This trend was also evident across several other psychometric scales related to emotional regulation, impulse control, and cognitive function, which are often associated with mental health risks^119–121^ (**Figure 1C**). Importantly, analyses based on the dimensional archetype scores across the entire sample yielded similar associations, demonstrating that the observed patterns are not restricted to individuals with clear categorical membership but reflect broader, continuous variation in mental health traits. (**Figure S8**). Consequently, in subsequent analyses, we used both categorical archetypes and dimensional archetype scores to enhance interpretability and the statistical power for discovery.

**Figure 1.**
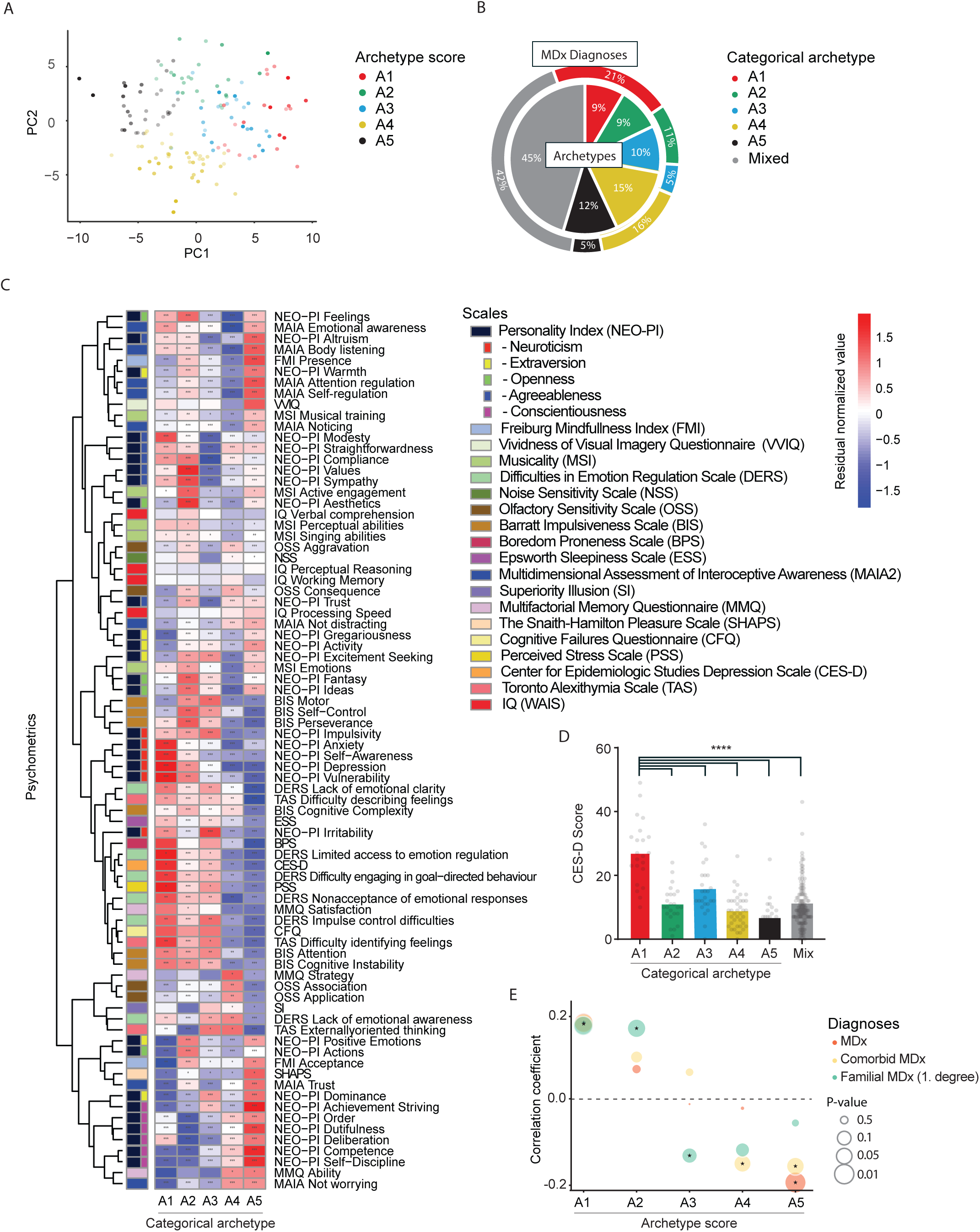
Identification and characterization of psychometric archetypes. **A)** PCA plot representing the archetype scores for the five archetypes (A1-5) projected in dimensions following principal component analysis. Each dot represents an individual, with color codes representing each of the five archetypes. Strength of colors represents the level of archetype affiliation. Only individuals with a dominating archetype affiliation are shown (membership > 0.5). **B)** Pie chart showing the percentage of individuals with a dominating affiliation to one of the five categorical archetypes and the mixed archetype group, as well as the percentage of individuals with a self-reported MDx in each archetype out of all individuals with a self-reported MDx. **C)** Heatmap showing the psychometric characteristics of each of the five categorical archetypes. All variables were rank-normally transformed, and test of significance of differences between individuals in a given archetype and all other individuals indicated (Mann-Whitney U test). Statistical significance level indicated by ∗p < 0.05, ∗∗p < 0.01, and ∗∗∗p < 0.001. Row-wise clustering was performed with the complete linkage method, as shown by the lines on the left. **D)** Plot of CES-D scores for each of the five categorical archetypes and the mix group. **E)** Correlations between archetype scores and any self-reported MDx diagnoses, more than one diagnosis (comorbid MDx), as well as MDx diagnoses among first-degree relatives, respectively. Statistical significance level indicated by size of the outer circle as specified in the legend on the plot.

### Psychometric archetypes are associated with differences in the prevalence of mental health diagnoses

In order to evaluate the clinical relevance of the archetypes, we assessed the distribution of participants’ self-reported diagnoses across the categorical archetypes. While having a dominant archetype on its own did not significantly increase the likelihood of having a diagnosis (**Table 1**, Kruskal-Wallis test: p = 0.052), individual categorical archetypes were associated with either increased or decreased prevalence of MDx diagnoses (**Figure 1B**). In line with its characteristic high neuroticism and depression scores, the prevalence of MDx diagnoses was significantly higher in the A1 group compared to the average among participants, with nearly one-third reporting one or more MDx diagnoses (**Figure 1B** and **Table 1**; prevalence ratio (PR) = 2.4, 95% CI 1.2-4.8, p = 0.012). Notably, reported diagnoses were not restricted to specific diagnostic categories but broadly represented the most common neurodevelopmental, anxiety, mood, personality, stress, eating, and substance-related disorders (**Table 1**). In contrast, while not statistically significant, individuals with a dominant A5 archetype profile appeared protective against MDx, with only one self-reported case in this group (**Figure 1B** and **Table 1**; PR = 0.25, 95% CI 0.04-1.78, p = 0.17).

**Table 1.** Distribution of self-reported diagnoses across discrete archetypes at baseline.

|  | Discrete archetypes |  |  |  |  | Mixed |
| --- | --- | --- | --- | --- | --- | --- |
|  | A1 | A2 | A3 | A4 | A5 |  |
| **# of individuals** | 22 | 24 | 26 | 39 | 30 | 117 |
| with any MDx diagnosis | 7 (32%) | 5 (21%) | 3 (12%) | 6 (15%) | 1 (3%) | 12 (10%) |
| with comorbid MDx diagnoses | 3 (14%) | 3 (13%) | 2 (8%) | 3 (8%) | 0 (0%) | 4 (3%) |
| with first-degree relatives with MDx diagnosis | 10 (45%) | 10 (42%) | 4 (15%) | 6 (15%) | 8 (27%) | 27 (23%) |
| **Diagnostic entities:** |  |  |  |  |  |  |
| ADHD | 2 | 2 | 2 | 0 | 0 | 2 |
| Anxiety or phobia | 2 | 1 | 0 | 3 | 1 | 3 |
| Depression | 4 | 2 | 2 | 5 | 0 | 8 |
| Substance use disorder | 1 | 0 | 1 | 0 | 0 | 1 |
| Personality disorder | 1 | 0 | 0 | 1 | 0 | 0 |
| Self-harm | 0 | 1 | 1 | 0 | 0 | 0 |
| Eating disorder | 2 | 1 | 0 | 0 | 0 | 0 |
| Stress-related disorder | 3 | 1 | 2 | 0 | 0 | 3 |

We then analyzed the correlation between individual dimensional archetype scores and the prevalence of MDx diagnoses, encompassing both single and comorbid conditions, as well as family history of MDx (**Table S5**). The A1 score exhibited a significant positive correlation with both diagnosed MDx (**Figure 1E**; Pearson’s correlation coefficient = 0.172; p = 0.0085) and psychiatric comorbidity (**Figure 1E**; Pearson’s correlation coefficient = 0.170; p = 0.0096), thus highlighting the A1 score as a significant marker of psychiatric risk. The significant correlation between A1 scores and MDx diagnoses among first-degree relatives (**Figure 1E**; Pearson’s correlation coefficient = 0.165; p = 0.0118) further suggests that individuals with a strong affiliation to the A1 archetype have an inherited predisposition to mental health problems. Whereas, on average, one-quarter of individuals in the cohort reported having first-degree relatives with diagnosed MDx, the distribution was not even across categorical archetypes (**Table 1** and **Table S5**; Kruskal-Wallis test: p = 0.024), with the highest proportion (45%) seen in the A1 archetype group.

While an elevated familial risk was observed for the dimensional A2 score (**Figure 1E**; Pearson’s correlation coefficient = 0.162; p = 0.0128), categorical A2 archetype affiliation was not significantly associated with an increased risk of MDx diagnosis, nor did dimensional A2 scores show a significant correlation with MDx diagnoses (**Figure 1E** and **Table 1**). For the A3 score, a significant negative correlation was observed with familial risk of MDx (**Figure 1E**; Pearson’s correlation coefficient = −0.131; p = 0.0458), and a similar trend was seen for the A4 and A5 scores (**Figure 1E**), which both displayed significant negative correlations with psychiatric comorbidity (A4: Pearson’s correlation coefficient = - 0.150; p = 0.0225. A5: Pearson’s correlation coefficient = −0.156; p = 0.0176). The A5 scores were further significantly negatively correlated with having any MDx diagnosis (**Figure 1E**; Pearson’s correlation coefficient = −0.194; p = 0.0031).

### Genetic associations with psychometric archetypes

To assess whether psychometric archetypes represent distinct, biologically grounded entities shaped by heritable components, we aimed to quantify the genetic contribution to the archetypes. Given the limitations of our sample size, we opted for an alternative to direct heritability analysis. Instead, we tested whether variation in archetype scores could be accounted for by PGSs derived from a wide range of mental health-related traits, including behavioral, cognitive, and neuroimaging phenotypes (**Table S3**). Notably, up to 7% of the variance in archetype scores could be explained by a PGS model based on miserableness (**Table S6**). Complementing these findings, analyses using categorical archetype membership as the outcome revealed the strongest associations with PGSs for brain imaging traits, particularly those related to resting-state fMRI, where predictive power reached Nagelkerke’s pseudo-R² of 0.08 (≈ 8%) (**Table S6**). Strikingly, individual archetype scores appeared to be driven by distinct sets of genetic influences, with few PGSs explaining a meaningful proportion of variance across more than one archetype score (**Figure 2** and **Table S7**). For the A1 archetype score, significant contributions were observed from PGSs for brain imaging traits as well as cognitive, behavioral, and clinical phenotypes (**Figure 2A-B** and **Table S7**). These included MDD and depression-related measures, neuroticism (R² = 0.02), autism spectrum disorder (ASD) (A1: R² = 0.03; A5: R² = 0.024), the well-being spectrum (A1: R² = 0.024), and bipolar disorder (BP) (A2: R² = 0.03) (**Figure 2B**). For the A2-4 archetypes scores, PGSs associated with cognitive traits consistently explained a significant proportion of the variance (**Figure 2B**). Additional contributions came from personality traits such as extraversion (A2 and A4), risk tolerance, and clinical traits including subjective well-being, BP, and panic disorder (A2), as well as depression- and anxiety-related measures (A3 and A4). PGSs related to resting-state fMRI connectivity and other brain measures also significantly influenced A2– A4 scores (**Figure 2A-B**). For the resilient A5 archetype, cognitive, behavioral, and clinical PGSs accounted for a substantial portion of the variance. This included multiple measures indexing broad genetic disposition to psychopathology, as captured by cross-disorder GWASs, as well as depression, miserableness, suicide attempt, ASD, ADHD, and anxiety (**Figure 2B**). PGSs for brain imaging traits, such as rostral middle frontal cortical thickness, weighted-mean ISOVF in the left medial lemniscus tract, and left thalamus nuclei AV volume, together with several resting-state fMRI connectivity measures, also contributed significantly to A5 scores (**Figure 2A**). Consistent with the observed prevalence of MDx diagnoses in the categorical A1 archetype group and reduced prevalence in the categorical A5 group, cross-disorder PGS and other clinical PGSs relating to psychiatric disorders all correlated positively with A1 archetype scores but negatively with A5 archetype scores (**Figure 2C-D** and **Figure S9**). For the neuroimaging traits, the direction of the correlation did, in some instances, differentiate between archetype scores, i.e., PGS for cortical average thickness in the caudate anterior cingulate region displayed a negative correlation to the A3 score but a positive correlation to the A5 score (**Figure S9**). Similarly, PGSs for cortical surface area in the Isthmus Cingulate were positively correlated with the A2 score, while negatively correlated with the A4 score (**Figure S9**).

**Figure 2.**
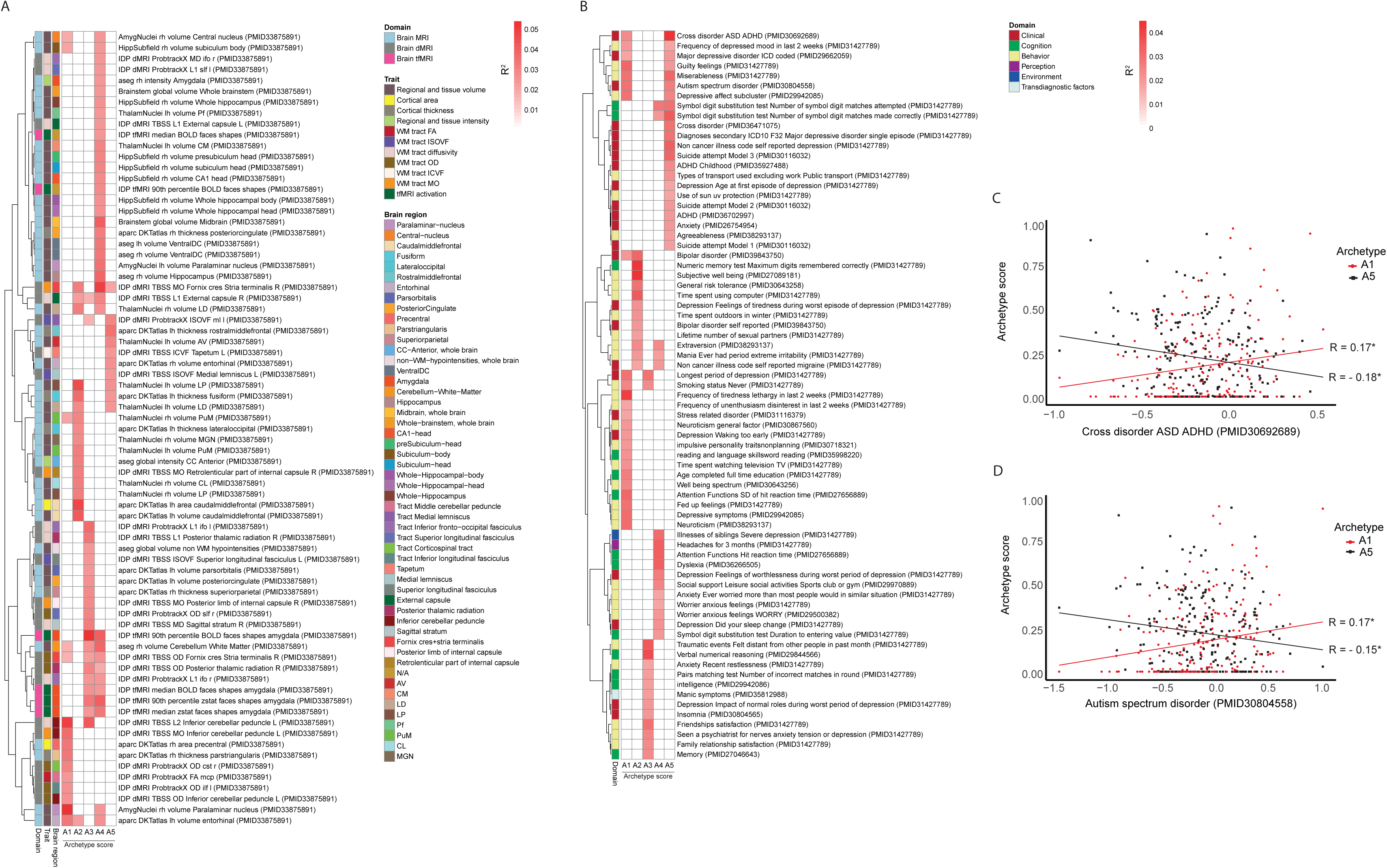
Genetic contribution to psychometric archetypes. Variance in archetype scores explained by polygenic scores (PGS). R² values are shown for significant models only (p<0.05). Top PGS predictors for each archetype, grouped by trait domain: **A)** brain imaging; and **B)** psychiatric, cognitive, and behavioral traits. Row-wise clustering was performed with the complete linkage method, as shown by the lines on the left of each heatmap. **C-D)** Archetype scores for A1 and A5 were positively and negatively associated, respectively, with cross-disorder and autism spectrum disorder PGSs, consistent with a vulnerability-resilience continuum. Statistical significance level indicated by ∗p < 0.05.

While no associations remained significant after FDR correction (q < 0.05), 693 nominally significant associations (p < 0.05) were observed (**Figure S9** and **Table S8**), revealing consistent bidirectional patterns across several PGSs, such as ASD^122^ and rfMRIconnectivity_ICA100edge230^70^. Opposing patterns between the A1 and A5 archetype scores reinforce the notion that A1 and A5 may represent extremes along a latent vulnerability-resilience continuum and support the biological plausibility of the archetypal framework.

### Neuroarchitectural features define individual archetypes

To comprehensively examine the neurobiological correlates of the identified archetypes, we assessed a wide array of structural brain imaging measures across participants, including cortical thickness, surface area, curvature metrics, vertex count, and gray matter volume, spanning both cortical and subcortical regions.

Across the five archetypes, we identified widespread and significant associations between archetype scores and multiple MRI-derived features (**Figure 3A-E** and **Table S9**). Interestingly, most associations were observed for scores of the intermediate archetypes A2-A4, rather than the two extremes (A1 and A5). These associations spanned multiple structural domains, including widespread changes in cortical curvature and surface area (A2), pronounced global cortical thinning (A3), and cortical and subcortical volume reductions (A4).

**Figure 3.**
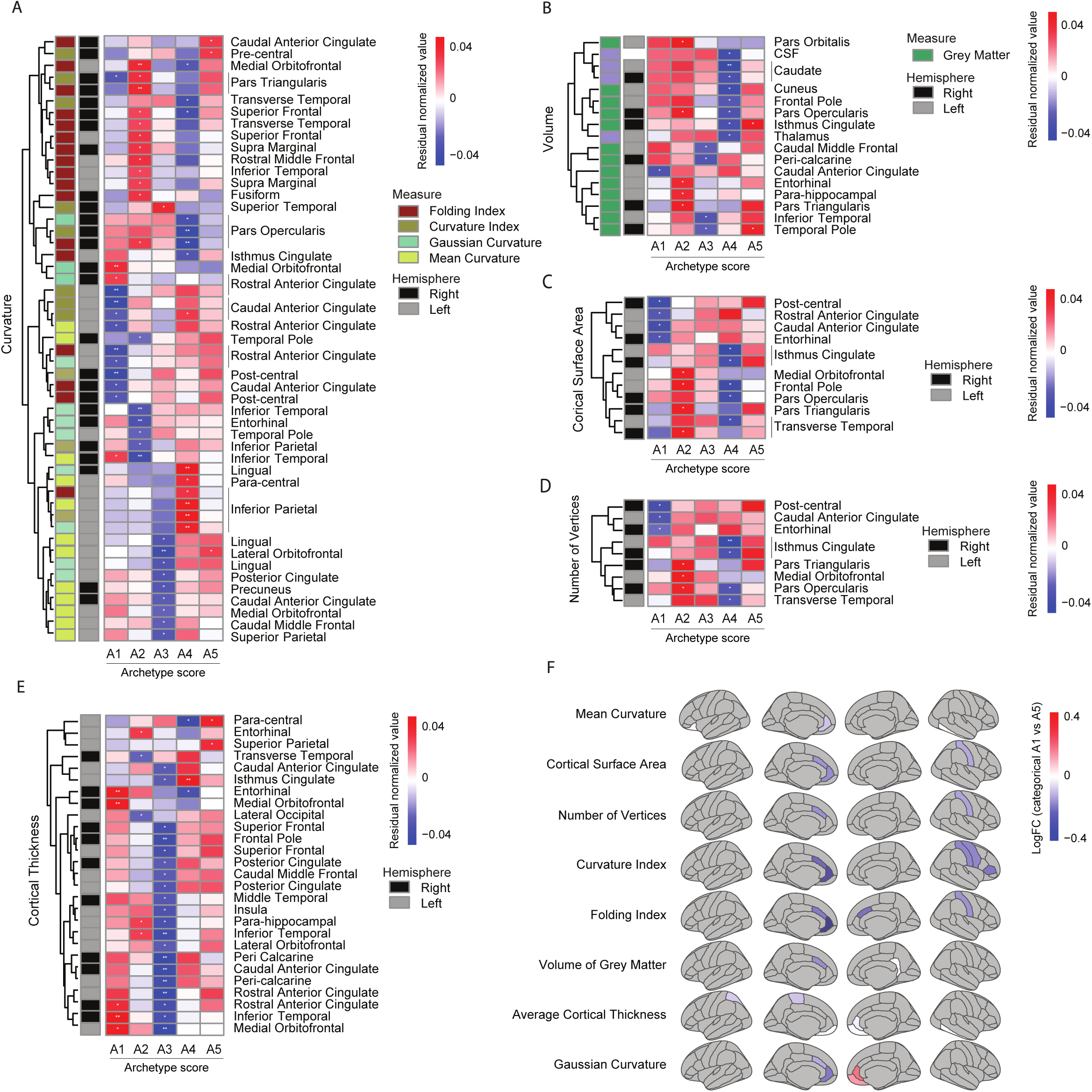
Neuroarchitectural correlates of psychometric archetypes scores. **A)** Archetype-wise differences in residual-normalized brain morphometry across **A)** curvature; **B)** Volume; **C)** surface area; **D)** number of vertices; and **E)** cortical thickness. **F)** Log fold-change in structural metrics between categorical A1 and A5 archetypes. Regional differences were observed in areas implicated in emotional and cognitive processing, including anterior cingulate, orbitofrontal cortex, entorhinal cortex, and insula.

The risk-associated A1 archetype score was characterized by reduced cortical surface area in the post-central, entorhinal, and anterior cingulate regions, decreased gray matter volume in the caudal anterior cingulate, and a lower number of vertices in these same regions (**Figure 3B-D**). Cortical thickness was increased covering the cingulate, orbitofrontal, and entorhinal cortex (**Figure 3E**), while curvature was increased in the orbitofrontal cortex, but decreased in the post-central and cingulate cortex (**Figure 3F**). In contrast, the A5 archetype score showed opposing patterns in many of these features, particularly in the post-central and cingulate cortex (**Figure 3A-E**). Directly comparing fold changes in measures between the A1 and A5 archetypes similarly revealed the most pronounced differences in entorhinal, prefrontal, and temporal regions, including the orbitofrontal cortex and cingulate subregions (**Figure 3F** and **Table S10**).

### Archetype scores are associated with distinct circulating metabolite signatures

To identify potential circulating biomarkers differentiating the five archetypes, we analyzed fasting-state plasma metabolites using ordinary least squares regression. Following thorough quality control procedures, we retained quantitative data for 433 metabolite features, of which 124 were successfully annotated. Despite the relatively limited number of participants with available metabolomic data, we identified 68 nominally significant metabolite-archetype associations (p < 0.05) (**Figure 4A** and **Table S11**). However, none of these associations remained statistically significant after correcting for multiple testing using the Benjamini-Hochberg FDR procedure (q < 0.05).

**Figure 4.**
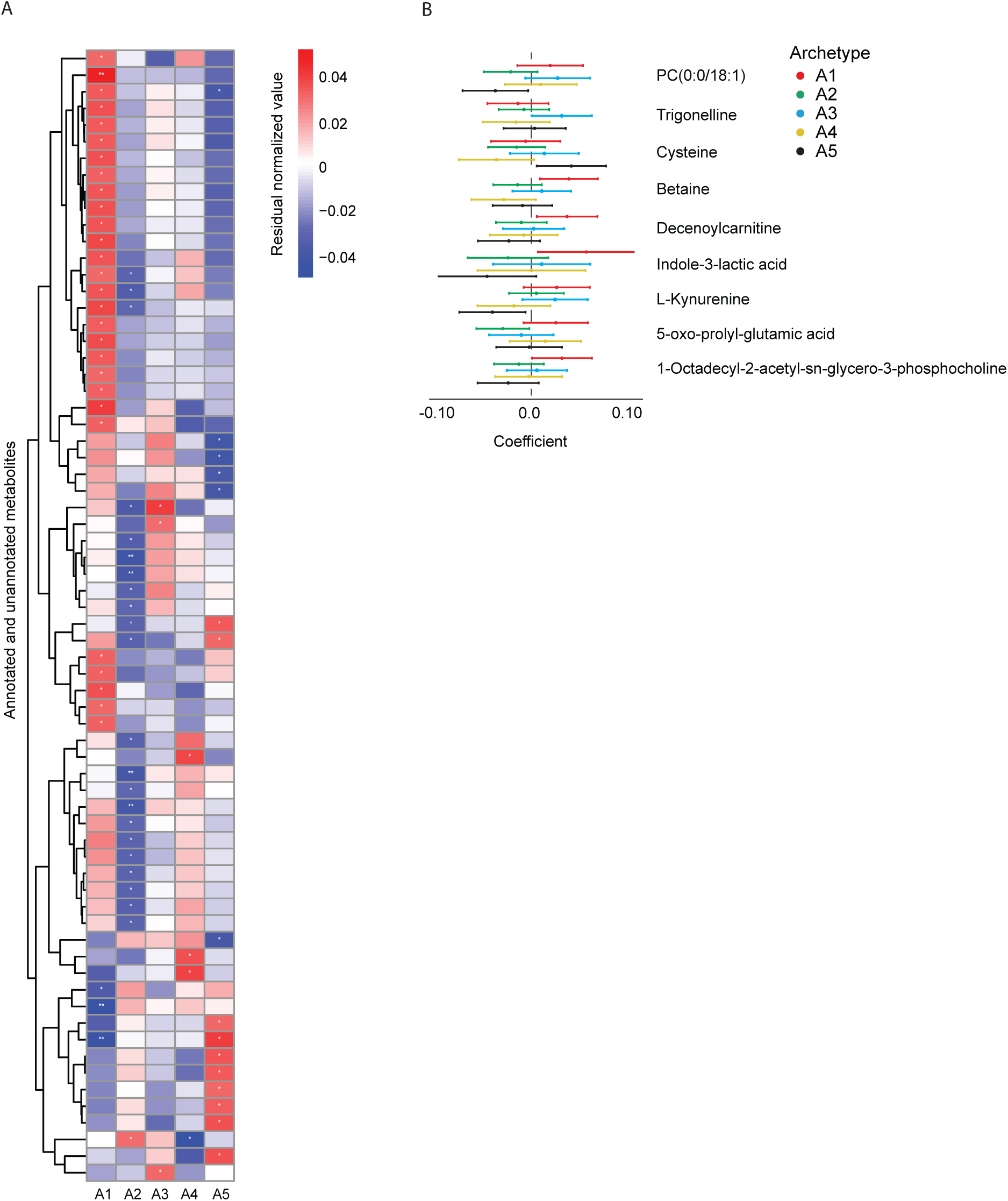
Archetype-specific differences in circulating metabolites. Residual-normalized abundances of metabolites across archetypes. **A)** All measured metabolites. **B)** Differentially abundant annotated metabolites. A1 is characterized by elevated levels of stress- and inflammation-related metabolites such as betaine, kynurenine, and indole-3-lactic acid, while A5 shows increased levels of cysteine and phospholipids associated with neuroprotection. Feature ID of metabolites are: Betaine: 415; Cysteine: 417; Decenoylcarnitine: 4044; Indole-3-lactic acid: 2162; L-Kynurenine: 1801; Trigonelline: 498; PC(0:0/18:1): 7451; 1-Octadecyl-2-acetyl-sn-glycero-3-phosphocholine: 10368; 5-oxo-prolyl-glutamic acid: 1188.

Among the annotated metabolites, A1 scores showed positive associations with betaine, decenoylcarnitine, indole-3-lactic acid, and 1-octadecyl-2-acetyl-sn-glycero-3-phosphocholine, an unknown indole derivative, and a possible sulfenyl compound (**Figure 4B** and **Table S11**). Strikingly, for most metabolites with nominal associations, coefficients were in opposite directions for A1 and A5, highlighting a mirrored metabolic profile across the vulnerability–resilience continuum. This included kynurenine, which showed a nominal negative association with A5 score and a positive association with A1 score. In contrast, cysteine was nominally positively associated with A5 scores but negatively associated with A1 scores.

While the annotated metabolites primarily highlighted associations with A1 and A5 scores, broader inspection of the full metabolomic feature set revealed numerous nominally significant associations across the intermediate archetype scores (A2-A4) as well (**Figure 4A-B**), underscoring the complex and multidimensional biological distinctions between archetypal profiles.

### Cross-modal integration of PGS, metabolomics, and MRI features with archetypes

To further explore the biological underpinnings of the extreme archetypes (A1 and A5), we constructed a correlation-based visualization integrating polygenic scores (PGSs), fasting-state plasma metabolites, and structural MRI features **(Figure 5A**). Rather than relying solely on the univariate associations presented earlier, we examined pairwise Spearman correlations to highlight both direct links between features and archetype scores as well as inter-feature correlations across modalities. This approach does not constitute formal network analysis but instead provides an intuitive representation of how genetic, neuroimaging, and metabolic features jointly contribute to archetype-specific biological signatures.

**Figure 5.**
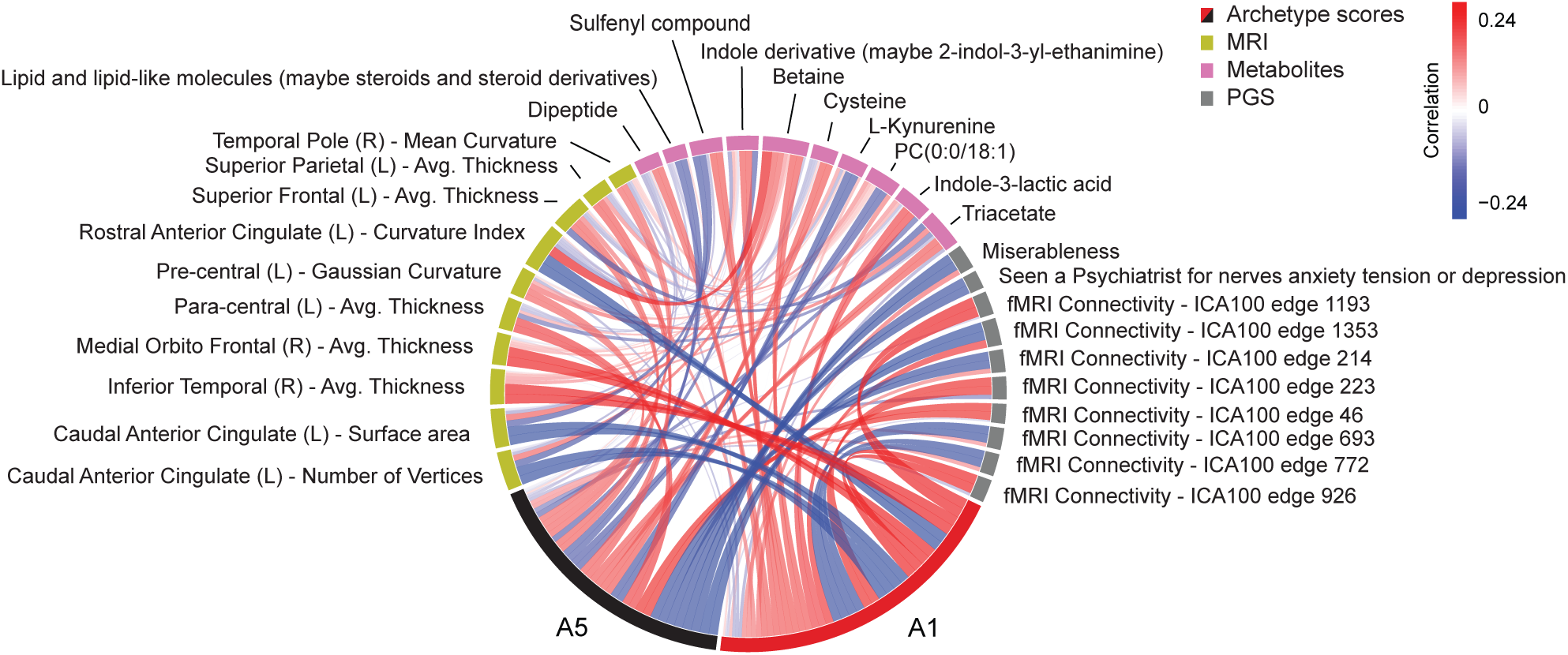
Integrated correlation-based network visualization of archetype-associated features across modalities. Multimodal correlation-based network visualization showing correlations between A1 and A5 archetype scores and their top associated features from brain imaging, polygenic scores, and metabolomics. The graph highlights cross-domain relationships, including shared associations with specific cortical features (e.g., anterior cingulate and inferior temporal cortex), polygenic risk (e.g., miserableness, fMRI connectivity), and metabolites (e.g., betaine, cysteine, kynurenine). Features are organized to emphasize contrasting patterns of vulnerability (A1) and resilience (A5). Feature ID of metabolites are: Betaine: 415; Cysteine: 417; Dipeptide: 1821; Indole-3-lactic acid: 2162; Indole derivative (maybe 2-indol-3-yl-ethanimine): 2171; Lipid and lipid-like molecules (maybe steroids and steroid derivatives): 10340; L-Kynurenine: 1801; PC(0:0/18:1): 7451; Sulfenyl compound: 974; Triacetate: 347

After correcting for multiple testing using the Benjamini–Hochberg FDR (α = 0.05), a total of 34 associations remained statistically significant (**Table S12**). These included several robust, cross-modal links involving archetype scores and features from all three data layers. For example, A1 scores were significantly positively correlated with cortical thickness in the right inferior temporal gyrus (ρ = 0.24, FDR-adjusted p = 0.008) and the right medial orbitofrontal cortex (ρ = 0.23, FDR-adjusted p = 0.008). A5 scores showed a significant negative correlation with the PGS for “miserableness” (ρ = –0.22, FDR-adjusted p = 0.008), while A1 scores showed a nominal positive association with the same score (ρ = 0.13, FDR-adjusted p = 0.101). Similarly, a PGS reflecting resting-state fMRI connectivity (ICA100-1353) was positively associated with A1 scores (ρ = 0.19, FDR-adjusted p = 0.015) and negatively with A5 scores (ρ = –0.22, FDR-adjusted p = 0.008).

Beyond these archetype–feature links, one significant cross-modal association emerged between a metabolite and an MRI trait: Betaine, which was nominally associated with A1 scores (ρ = 0.16, p = 0.034, FDR-adjusted p = 0.090), showed a significant positive correlation with cortical curvature in the left rostral anterior cingulate cortex (ρ = 0.24, FDR-adjusted p = 0.008). Notably, this cortical curvature feature itself was negatively associated with A1 scores (ρ = –0.24, FDR-adjusted p = 0.007). Collectively, this illustrates how correlation-based integration can reveal coherent multimodal biological signatures of mental health variation, reinforcing the interconnected nature of the features shaping archetypal risk and resilience.

### Five-Year Follow-Up: Archetype Stability and Mental Health Outcomes

To assess the long-term relevance of baseline psychometric archetypes, we recontacted participants approximately five years after their initial enrollment. A total of 214 individuals completed the MHQ2 questionnaire, providing detailed self-reported data on current symptoms, psychiatric diagnoses, and psychosocial functioning.

We first examined whether baseline archetype scores were associated with MHQ2 outcomes five years later (**Figure 6A**). A1 scores were strongly negatively correlated with resilience (Brief Resilience Scale, r = – 0.42, FDR p < 1e–6) and positively correlated with loneliness (UCLA, r = 0.35, FDR p = 0.00003), depression severity (PHQ-9, r = 0.33, FDR p = 0.00005), and anxiety (GAD-7, r = 0.22, FDR p = 0.0035). Moreover, A1 scores were associated with an increased likelihood of melancholic depression diagnosis (r = 0.25, FDR p = 0.0078), consistent with a stable risk profile over time. A5 scores showed an opposing pattern, with lower loneliness (UCLA, r = −0.21, FDR p = 0.026), lower depression scores (PHQ-9, r = - 0.20, FDR p = 0.026), and higher resilience (Brief Resilience Scale, r = 0.22, FDR p = 0.01), although associations were generally weaker. We then examined point-biserial correlations between archetype scores and MHQ2 binary diagnostic and experience-based outcomes (**Figure 6A**). Here, findings included associations between A2 scores and cannabis exposure (r = 0.33, FDR p = 0.00038), A3 scores and childhood trauma (r = 0.27, FDR p = 0.005) and GAD-derived anxiety (r = 0.22, FDR p = 0.0178), and between A4 scores and reduced prevalence of panic attacks (r = −0.27, FDR p = 0.005) and GAD-derived anxiety (r = −0.17, FDR p = 0.076). Collectively, these findings suggest that the archetype profiles identified at baseline not only relate to current mental health symptom burden but also align with lifetime mental health history and adverse psychological experiences. Overall, the results indicate a high degree of stability in the psychological and clinical relevance of archetypes over time.

**Figure 6.**
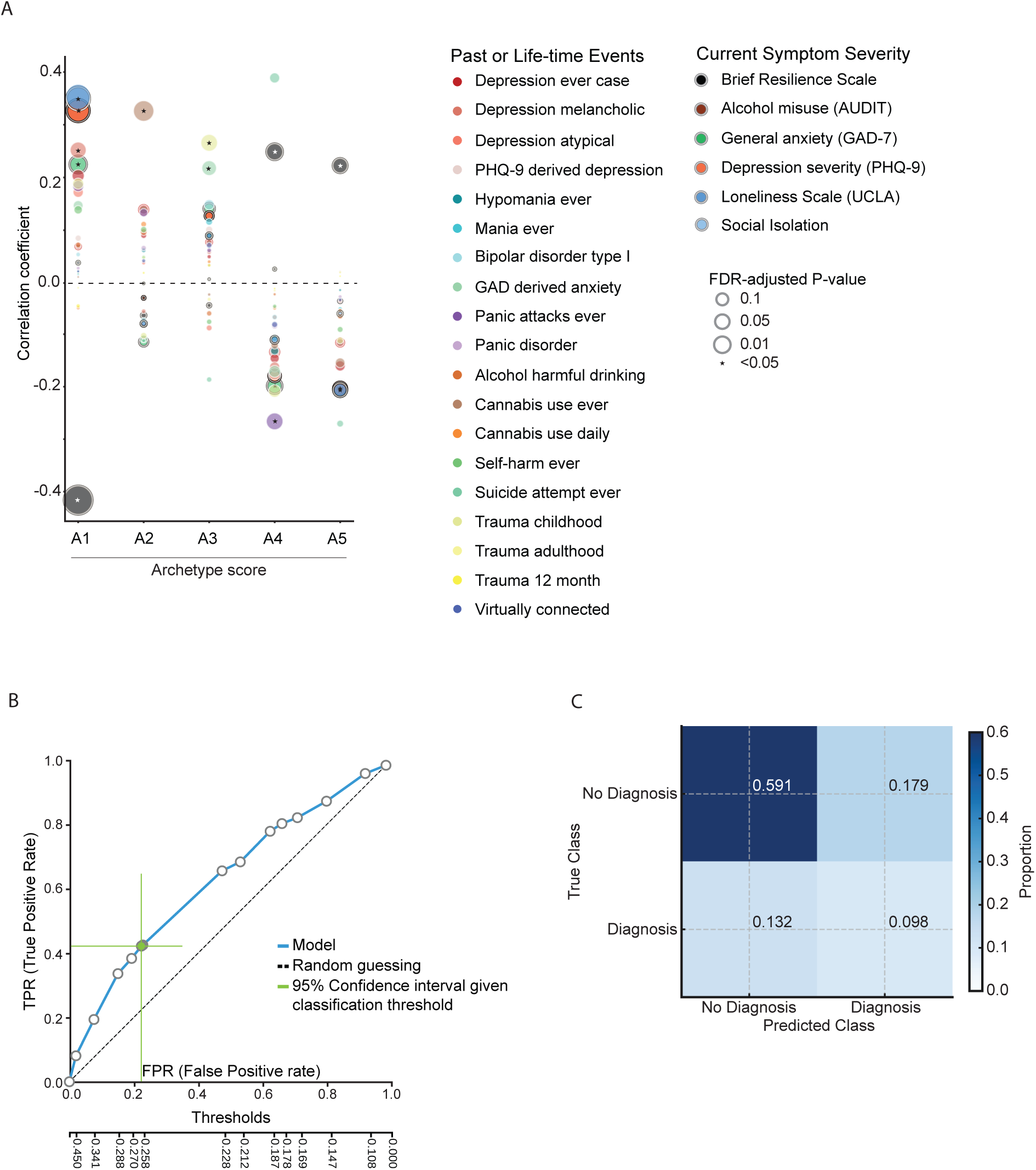
Archetype profiles are associated with distinct symptom patterns and predict future psychiatric diagnoses. **A)** Correlations between baseline archetype scores (A1-A5) and a range of current symptoms, lifetime psychiatric traits, trauma exposure, and psychosocial outcomes measured at five-year follow-up. Colored bars indicate significant associations (FDR-adjusted p < 0.05); effect sizes are represented as Pearson correlation coefficients. Archetype A3 showed strong associations with anxiety, depressive symptoms, and childhood trauma, whereas A1 was associated with a broader range of psychopathology and adverse life experiences. **B**) PR AUC illustrating the performance of a support vector machine trained to predict new-onset psychiatric diagnoses at five-year follow-up based on the top 5 features from each of the biological modalities as well as psychometrics most strongly associated with A1. The support sector machine was of type C-SVC with a polynomial kernel and hyper-parameters: cost = 0.001, gamma = 0.01, degree = 2. Green line indicates 95% confidence interval around the true positive rate across classification thresholds, where the classification threshold was determined by balanced accuracy. **C**) Confusion matrix for diagnosis prediction in the training set, showing the proportion of true and predicted cases across diagnostic outcome classes for the best performing model, where the classification threshold was set by balanced accuracy. The model achieved moderate discrimination between all-time cases and non-cases, with the highest classification accuracy for individuals without diagnoses.

### Predicting Future Mental Health Diagnoses Using Baseline Biosignatures

To evaluate whether psychometric archetypes predict mental health outcomes, we examined self-reported psychiatric diagnoses across categorical archetypal groups obtained approximately 5 years after baseline. In total, 74 participants reported at least one diagnosis at baseline or follow-up, including 40 newly diagnosed since baseline (**Table 2** and **Figure S15**). The risk-associated A1 archetype group showed the highest prevalence (14/22; 64%), including 7 new cases, while the resilient A5 archetype group had the lowest (5/30; 17%). The categorical mixed group, reflecting its larger size, accounted for the highest absolute number of cases (24/116; 21%), including 12 new cases. Interestingly, A3 showed the highest proportion of incident diagnosis, with 9 new cases since baseline. Comorbidity concentrated in the A1 (6/22) and the mixed group (11/116), with only one A5 individual reporting multiple diagnoses. Reported conditions were heterogeneous but most often included depression and ADHD, particularly in A1 and the mixed group. No participants reported schizophrenia or bipolar disorder. ASD and PTSD were confined to A3 and mixed groups, with eating and anxiety disorders showing sparse distribution across archetypes.

**Table 2.**
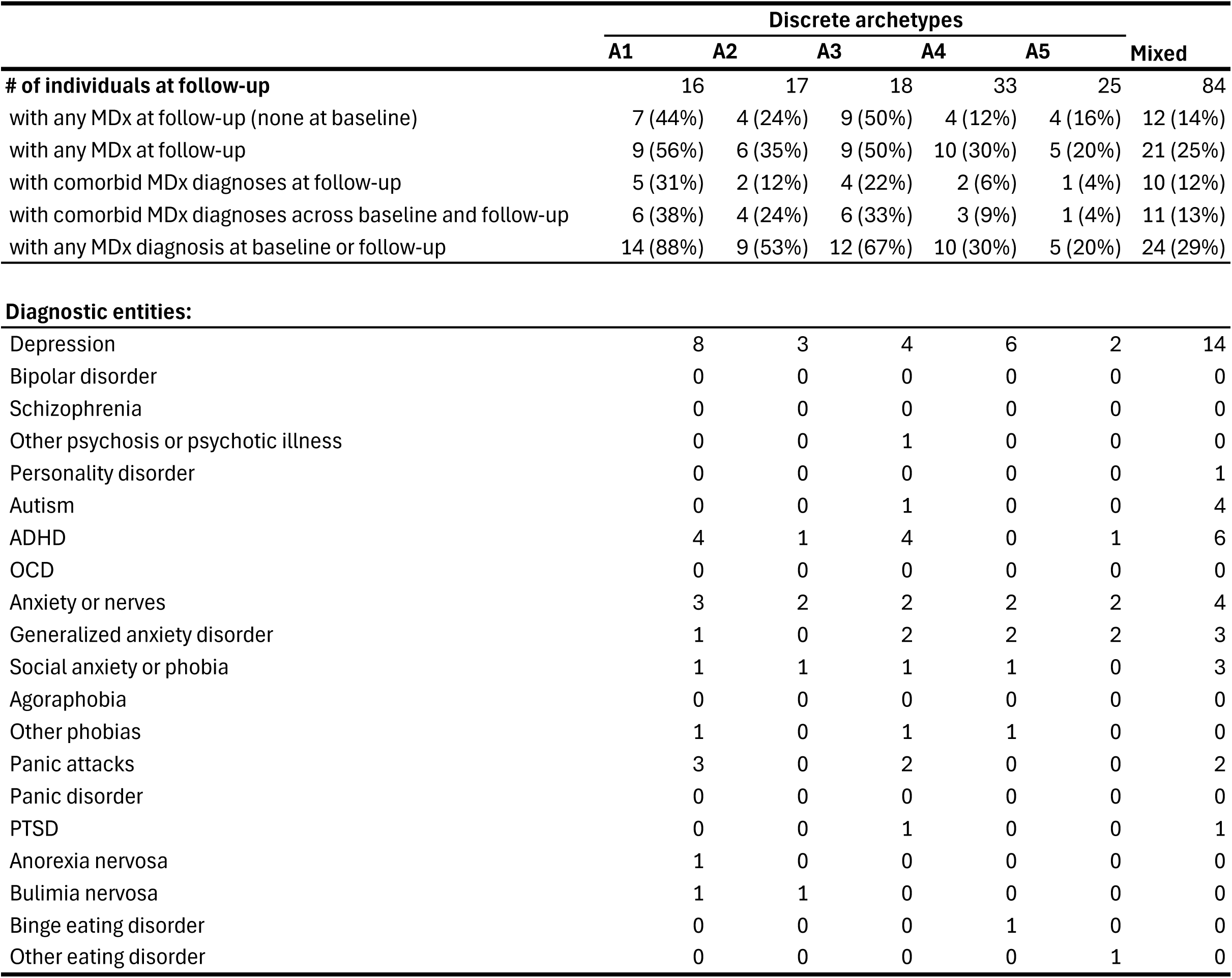
Distribution of self-reported diagnoses across discrete archetypes at follow-up.

|  | Discrete archetypes |  |  |  |  |  |
| --- | --- | --- | --- | --- | --- | --- |
|  | A1 | A2 | A3 | A4 | A5 | Mixed |
| **# of individuals at follow-up** | 16 | 17 | 18 | 33 | 25 | 84 |
| with any MDx at follow-up (none at baseline) | 7 (44%) | 4 (24%) | 9 (50%) | 4 (12%) | 4 (16%) | 12 (14%) |
| with any MDx at follow-up | 9 (56%) | 6 (35%) | 9 (50%) | 10 (30%) | 5 (20%) | 21 (25%) |
| with comorbid MDx diagnoses at follow-up | 5 (31%) | 2 (12%) | 4 (22%) | 2 (6%) | 1 (4%) | 10 (12%) |
| with comorbid MDx diagnoses across baseline and follow-up | 6 (38%) | 4 (24%) | 6 (33%) | 3 (9%) | 1 (4%) | 11 (13%) |
| with any MDx diagnosis at baseline or follow-up | 14 (88%) | 9 (53%) | 12 (67%) | 10 (30%) | 5 (20%) | 24 (29%) |
| **Diagnostic entities:** |  |  |  |  |  |  |
| Depression | 8 | 3 | 4 | 6 | 2 | 14 |
| Bipolar disorder | 0 | 0 | 0 | 0 | 0 | 0 |
| Schizophrenia | 0 | 0 | 0 | 0 | 0 | 0 |
| Other psychosis or psychotic illness | 0 | 0 | 1 | 0 | 0 | 0 |
| Personality disorder | 0 | 0 | 0 | 0 | 0 | 1 |
| Autism | 0 | 0 | 1 | 0 | 0 | 4 |
| ADHD | 4 | 1 | 4 | 0 | 1 | 6 |
| OCD | 0 | 0 | 0 | 0 | 0 | 0 |
| Anxiety or nerves | 3 | 2 | 2 | 2 | 2 | 4 |
| Generalized anxiety disorder | 1 | 0 | 2 | 2 | 2 | 3 |
| Social anxiety or phobia | 1 | 1 | 1 | 1 | 0 | 3 |
| Agoraphobia | 0 | 0 | 0 | 0 | 0 | 0 |
| Other phobias | 1 | 0 | 1 | 1 | 0 | 0 |
| Panic attacks | 3 | 0 | 2 | 0 | 0 | 2 |
| Panic disorder | 0 | 0 | 0 | 0 | 0 | 0 |
| PTSD | 0 | 0 | 1 | 0 | 0 | 1 |
| Anorexia nervosa | 1 | 0 | 0 | 0 | 0 | 0 |
| Bulimia nervosa | 1 | 1 | 0 | 0 | 0 | 0 |
| Binge eating disorder | 0 | 0 | 0 | 1 | 0 | 0 |
| Other eating disorder | 0 | 0 | 0 | 0 | 1 | 0 |

To assess the potential of archetype-related features for predicting later mental health outcomes, nine supervised classification models were trained on baseline data to identify individuals with a self-reported diagnosis at follow-up. Two feature selection strategies were applied: an archetype-guided approach using top A1 or A1/A5 features from each modality (6 models) and an agnostic approach using full-dataset feature selection via LASSO or SES (3 models). Both were trained on psychometric-only, biology-only (PGSs, imaging, metabolites), and combined data. The training set included 78 subjects, and the test set 19.

Models based on A1- and A5-guided features outperformed agnostic approaches, but all models reached statistical significance compared to baseline (**Table 3**). The best-performing model, an A1-derived SVM combining psychometric and biological data, achieved a PR AUC of 0.801 in internal cross-validation (**Figure 6B**). Overall, the models guided only by A1 features performed marginally better than those guided by both A1 and A5 features. The optimal psychometry-only model, again an A1-derived SVM, performed nearly as well, with an area under the curve of 0.782. Comparably, the optimal biology-only model, an A1-derived Ridge Logistic Regression, reached 0.780. The confusion matrix from the top-performing model indicated that the model correctly classified 59.1% of participants without and 9.8% with a diagnosis (false positives = 17.9%; false negatives = 13.2%) (**Figure 6C**). A similar pattern was observed for the rest of the models based on A1- and A5-guided features, although they tended to have higher false positive rates (**Figure S16D-H**). In contrast, models based on agnostic feature selection performed relatively worse (**Figure S16A-C** and **Table 3**). The best-performing model in this group, which used psychometric-only data, achieved a PR AUC of 0.737, with comparable misclassification rates. It should be noted that their 95% confidence intervals did overlap with the A1- and A5-guided models, although the agnostic model’s intervals were generally 0.05 lower.

**Table 3.**
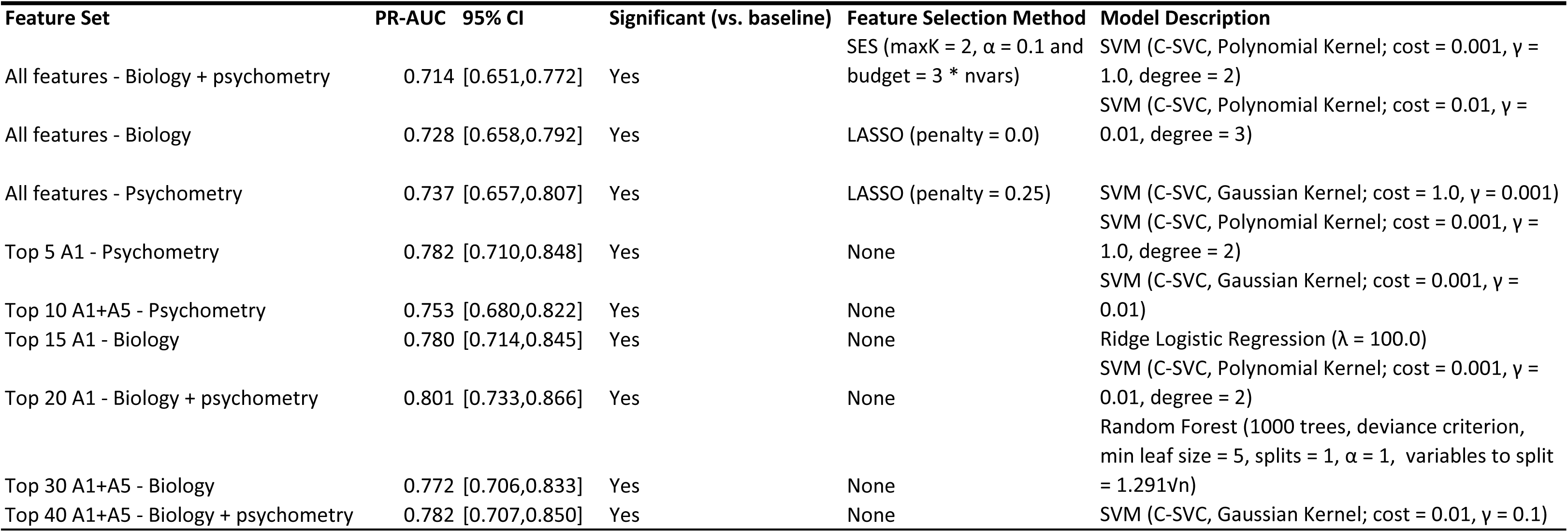
Predictive performance of prediction participants with any MDx at follow-up in training and modeling details for tested feature sets.

To evaluate generalizability, we tested the best-performing model, the A1-derived model combining psychometry and biology, on a held-out test sample. Performance declined modestly, achieving a PR AUC of 0.700. With the classification threshold set to 0.259, derived from balanced accuracy, the model did not predict any participants as diagnosed, correctly identifying 0.0% without and 21.1% with diagnoses (false positives = 78.9%; false negatives = 0%). When applying a threshold of 0.450, determined by overall accuracy, the model correctly identified 36.8% of non-diagnosed and 21.1% of diagnosed participants (false positives = 63.2%; false negatives = 5.3%).

These findings demonstrate that archetype-informed feature selection enables interpretable, biologically grounded prediction of mental health outcomes. Despite modest reductions in performance in the independent sample, likely attributable to sample limitations and phenotypic heterogeneity, vulnerability- and resilience-related features remained informative for future mental health risk. Further validation in larger cohorts is warranted to refine and strengthen predictive accuracy.

## DISCUSSION

This study introduces a novel framework for mental health stratification by identifying psychometric archetypes in a general population sample and linking them to multimodal biological signatures. To our knowledge, it is among the most comprehensive community-based investigations integrating psychometric, neuroanatomical, genetic, and metabolic data. Rather than relying on case–control comparisons, we applied a dimensional framework to young adults within the typical age of onset for psychiatric disorders, capturing the continuum of psychological traits and vulnerability profiles. This approach enables characterization of subclinical conditions and heterogeneous experiences in individuals who may not meet diagnostic criteria but nevertheless carry substantial psychological burden and are often on trajectories toward a clinical condition. In this way, the study provides a proof-of-concept for biology-informed risk stratification. Our identification of five distinct psychometric archetypes underscores the continuous and multifaceted nature of mental health-related traits. These archetypes reflect a broad spectrum of cognitive, emotional, and behavioral patterns, each of which is differentially associated with mental health outcomes and underpinned by coherent biological variation spanning genetic profiles, structural brain metrics, and circulating metabolites. In particular, A1 and A5 represent the extremes of a vulnerability-resilience continuum, each characterized by distinct and opposing biological profiles across genetic, neuroimaging, and metabolic domains.

Archetype A1, marked by high neuroticism and low extraversion, is associated with elevated broad psychiatric polygenic risk as well as a higher likelihood of psychiatric diagnoses and comorbidity at baseline. These associations are consistent with the extensive literature linking neuroticism to common mental disorders^123^. However, A1 also captures unique combinations of traits with limited overlap with traditional personality dimensions, such as elevated impulsivity (BIS), anhedonia (SHAPS), alexithymia (TAS), negative thinking (CES-D), and emotional reactivity (PSS). These contribute to a broader vulnerability profile. Neuroanatomically, A1 score is associated with increased cortical thickness in medial orbitofrontal and inferior temporal areas, regions implicated in emotional regulation and mood^124^. While cortical thinning is frequently reported in psychiatric case-control studies^125^, particularly in BP and schizophrenia (SZ), there is accumulating evidence that increased cortical thickness may be a feature of early, subclinical, or genetically predisposed states. For instance, recent large-scale population studies have shown that higher polygenic risk for SZ is associated with greater cortical thickness in typically developing youth^126^. Similarly, increased thickness in medial and lateral prefrontal cortices has been observed in untreated or early-stage MDD^127^. These patterns are thought to reflect compensatory or maladaptive remodeling that precedes cortical thinning in later stages of illness. In addition, A1 shows reduced surface area and gray matter volume in the cingulate and post-central cortex, which support emotional processing and awareness^128,129^, and have been linked to depression and anxiety^130^. In our data, structural cortical changes in A1 may thus reflect disrupted cognitive-affective integration, representing a neural correlate of early mental health vulnerability. As structural brain measures were not corrected for total brain or intracranial volume, this may influence interpretation. However, because such differences are commonly observed across sexes^131^, and we did not observe substantial sex bias in our archetypes, the impact is expected to be minimal.

Metabolically, the A1 score was associated with elevated levels of betaine, kynurenine, indole-3-lactic acid, and 9-decenoylcarnitine, all previously implicated in neuropsychiatric pathophysiology. Betaine participates in methylation and homocysteine metabolism, processes essential for brain function and linked to psychopathology in both preclinical^132^ and clinical research^133,134^. 9-Decenoylcarnitine, an acylcarnitine associated with mitochondrial function and cholinergic signaling, has been found to be altered in SZ^135^. Indole-3-lactic acid and kynurenine are both derived from tryptophan in the gut and intestines and linked to neurodevelopment, with systemic levels found to be altered in several psychiatric disorders^136–139^. Kynurenine stems mainly from endogenous biosynthesis^140,141^, whereas indole-3-lactic acid is largely of bacterial origin, particularly Bifidobacterium species that metabolize dietary tryptophan^142^. The observed elevations in both metabolites may therefore reflect a combined shift toward the kynurenine pathway and altered gut–brain axis signaling, consistent with previous reports of microbial involvement in psychiatric disorders^142–144^.

Conversely, A5 is defined by low neuroticism, high extraversion, along with heightened interoceptive awareness, visual imagery capacity, sleep regularity, and adaptive coping strategies, consistent with a more resilient and self-regulated cognitive–emotional profile^145–147^. It is negatively associated with psychiatric PGSs and positively associated with polygenic resilience traits. Neurostructural markers include greater cortical folding and surface area in fronto-cingulate regions, patterns that have been associated with cognitive flexibility, emotional regulation, and psychological resilience in prior studies^148^. Greater gyrification and surface expansion in these areas are thought to reflect enhanced integrative capacity and adaptability, consistent with findings from structural imaging studies of healthy individuals with resilience to adversity^148^. Metabolically, A5 score is associated with signatures suggestive of protective or homeostatic regulation, including elevated cysteine, an amino acid involved in antioxidant defense and glutathione synthesis^149^. Cysteine availability is a critical determinant of brain redox status, and its pharmacological precursor, N-acetylcysteine (NAC), is under investigation in neurological and psychiatric disorders, where it has shown promise in alleviating symptoms through antioxidant and glutamatergic mechanisms^150,151^.

Archetype A3 presents an instructive case that underscores the complexity of risk pathways to psychopathology. Despite minimal familial loading, A3 was associated with elevated levels of childhood trauma, high baseline depressive symptoms (CES-D), and a substantial number of new diagnoses at follow-up. Notably, this archetype also showed a broad pattern of structural brain differences, particularly in cortical thickness, suggesting that biological alterations are nonetheless present. This raises the possibility that A3 reflects an environmentally mediated risk trajectory, where early adversity becomes biologically embedded through long-term effects on brain development. Understanding such divergent trajectories may help guide targeted prevention and intervention strategies.

The delineation of these archetypes highlights how combinations of cognitive, emotional, and behavioral traits converge with multi-modal biology to shape mental health outcomes. Our findings extend prior biomarker research by embedding biological correlates within a dimensional, population-based stratification model. The biological coherence of A1 and A5 supports their utility as intermediate phenotypes and candidate risk/protective endotypes.

Critically, the predictive validity of the archetypes was tested using MHQ2 data collected five years after baseline. A1 and A5 scores remained strongly associated with symptom severity and diagnostic outcomes, demonstrating trait-like stability and real-world relevance. These results suggest that psychometric archetypes can be used for risk assessment by identifying individuals at long-term risk even in the absence of a clinical diagnosis at baseline. When these at-risk individuals have been identified, potential preventive measures can be employed to impede further mental health decline.

We note that the presented network analysis is a correlation-based visualization intended to highlight cross-modal patterns. Future work should employ formal network methods, such as community detection and centrality analyses, to uncover organizational structure beyond pairwise associations and to improve predictive accuracy, which will require larger samples.

Importantly, our archetypes demonstrated predictive validity: A1 and A5 scores remained strongly associated with symptom severity and diagnostic outcomes five years after baseline, indicating trait-like stability and clinical relevance. Archetype-guided models built on top features outperformed unconstrained feature selection from the full dataset, underscoring the advantage of psychologically informed stratification. Predictive performance declined slightly in the held-out test data, which is expected given the modest sample size and the complexity of psychiatric outcomes. Despite this, we still achieved a fairly high PR AUC of 0.70 in the test set, comparable to or better than similar studies in the field^161–163^. Thereby, the models retained the ability to distinguish between individuals who received and didn’t receive a diagnosis at the five-year follow-up. This supports the potential of archetype-informed modeling as a scalable tool for early identification of individuals at risk for mental health disorders.

### Perspectives

This study challenges the traditional binary distinction between illness and non-illness by adopting a dimensional framework grounded in psychological archetypes. Rather than pursuing isolated biomarkers within single data layers, we advocate for an integrative approach that reflects the multifaceted and biologically embedded nature of mental health. In this proof-of-concept study, we demonstrate that archetype-informed models can identify meaningful psychological and biological patterns associated with future psychiatric outcomes, offering a scalable and interpretable strategy for early risk detection.

Future research should build on this foundation by refining archetype definitions, increasing the granularity of biological features, and rigorously testing clinical utility in larger and more diverse populations. Extending archetyping to predict treatment response may further strengthen its translational potential. Importantly, applying this approach prospectively across broader age ranges, clinical contexts, and health systems will be key to evaluating its practical utility.

In summary, we propose a population-level framework for understanding individual differences in mental health that is grounded in psychological structure and supported by multi-layered biology. This approach offers a path toward more precise, integrated, and preventive strategies in psychiatric research and mental health screening.

## Data Availability

Data cannot be shared publicly because it is part of an ongoing study and is thus considered unanonymised under Danish law even if pseudonymized. Researchers who wish to access the data may contact Dr. Kristian Sandberg at The Center of Functionally Integrative Neuroscience and/or The Technology Transfer Office at Aarhus University, Denmark, to make a data sharing contract. After permission has been given by the relevant data committee, data will be made available to the researchers.

## Acknowledgments

This manuscript is based on work from COST Action CA18106 (The Neural Architecture of Consciousness), supported by COST (European Cooperation in Science and Technology). The study was supported by grants from The Lundbeck Foundation (ADB and PQ), The Jascha Foundation (PQ), Direktør Kurt Bønnelycke og Hustrufru Grethe Bønnelyckes Fond (PQ), Oda and Hans Svenningsens Fond (PQ), the Carlsberg Foundation (CF22-0132 to TLK), and The Danish Foundation for Research in Neurology (KS). We extend our special thanks to Alexander Fjældsted, Francesca Fardo, Inga Griskova-Bulanova, Martin Dresler, and Henrique Fernandes for their contributions to identifying the questionnaires utilized in this research. We thank Claude Bajada for providing MRI analysis scripts/processed data. We would like to thank GenomeDK and Aarhus University for providing computational resources and support that contributed to these research results. We also wish to acknowledge Sierra Lyn Wittrup Olsen for her meticulous assistance with DNA extraction.

## Author contributions

Conceptualization: PQ

Methodology: KS, FO, ME, JBG, NM, DKS, ABC, PQ, JGD, JDiC, BZ, TLK, SD, JB, MGM, MY, JG, NUR, DPM

Investigation: NM, ABC, TK, SD, JGD, FO, DKS, KS

Visualization: NM, ABC, PQ

Funding acquisition: NM, KS, ADB, PQ, TLK

Project administration: PQ, KS

Supervision: KS, PQ

Writing - original draft: NM, ABC, and PQ

Writing - review & editing: All authors

