## Supplementary Material for "Psychometric archetypes reveal biological signatures of vulnerability, resilience, and future mental health"

### Identification of multimodal mental health signatures in the young population using deep phenotyping

#### Supplementary Material

### Supplementary text

#### Supplementary Methods and Materials

##### Metabolomics

*Sample preparation*

Plasma samples (100 µl) were randomly distributed over 3 96-well plates (batches). A batch of plasma was created before the sample preparation and stored at −80 °C, referred to as external control (EC) samples. Both EC samples and plate-specific pools of all plasma samples in each batch were analyzed for quality control purposes. Each plate included eight EC, four process blanks (PB, extraction in empty well), four pooled samples, and 80 analytical samples. All solvents were LCMS-grade and were purchased from Thermo Fisher Scientific (Waltham, MA, USA). 300 µL of cold methanol/acetonitrile (50/50) was added to the samples, with subsequent shaking for 15 min at 900 rpm at room temperature. Samples were then frozen at -20 °C for 1 hour and consecutively centrifuged at 4000 rpm for 30 min at 4 °C. 50 µL of extract was pipetted into a new 96-well polypropylene plate, which was then evaporated under nitrogen for 1 h at 60 L/min, at room temperature. The samples were reconstituted in 100 µL of reconstitution solution (comprised of 5% solvent B in 95% solvent A, see Metabolomics Profiling section), shaken at 600 rpm for 15 min at room temperature, and then centrifuged at 3000 rpm for 10 min at 4 °C. Afterward the samples on the plate were pooled into a single well on a deep well plate, and pipetted into the four pool positions on the plate, which was then sealed with a silicone lid and centrifuged at 3000 rpm for 5 min at 4 °C. The plate was then run on the LC-MS/MS platform. All pipetting steps were performed on a Microlab STAR automated liquid handler (Hamilton Bonaduz AG, Bonaduz, Switzerland).

*Metabolomics profiling*

The LC-MS/MS platform consisted of timsTOF Pro mass spectrometer with an Apollo II ion-source for electrospray ionization, Bruker Daltonics (Billerica, MA, US) coupled to a UHPLC Elute LC system, Bruker Daltonics (Billerica, MA, US). The chromatographic separation system included a binary pump, an autosampler with cooling function, and a column oven with temperature control. For infusion of the reference solution, used for external and internal mass calibration, an additional isocratic pump, Azura Pump P4.1S (Knauer, Berlin, Germany) was used. The analytical separation was performed on an Acquity HSS T3 (100 Å, 2.1 mm x 100 mm, 1.8 µm) column (Waters, Milford, MA, US). The mobile phase consisted of solvent A (99.8% water and 0.2% formic acid) and B (49.9% methanol, 49.9% acetonitrile, and 0.2% formic acid). The analysis started with 99% mobile phase A for 1.5 min, thereafter a linear gradient to 95% mobile phase B during 8.5 min followed by an isocratic condition at 95% mobile phase B for 2.5 min before going back to 99% mobile phase A and equilibration for 2.4 min. Total run time for each injection was 15 min and the analysis time for a full 96-well plate was approximately 25 h. Samples were maintained at +15°C in the autosampler, and 5 µL were loaded to the column with a flow rate of 0.4 mL/min and a column temperature of 40 °C.

Tandem mass spectrometric analysis on the timsTOF Pro was performed in the Q-TOF mode with TIMS off, and auto MS/MS on using the following settings: ionization mode set to positive ionization, mass range set to 20 - 1100 *m/z* and a Spectra Rate of 9 Hz (Sample time 0.11s). Source settings as Capillary: 4500 V, Nebulizer Gas: 2.2 Bar, Dry Gas flow: 10 l/min, Dry Gas temperature: 220 °C. Tune settings as follows: Funnel 1 RF and Funnel 2 RF: 200Vpp, isCID: 0 eV, Multipole RF: 60 Vpp, Deflection Delta: 60 V, Quadrupole Ion Energy: 5 eV with a low mass set to 60 *m/z*, Collision Cell Energy set to 7 eV with a pre-Pulse Storage of 5 µs. Stepping is used in Basic Mode with a Collison RF from 250 - 750 Vpp, Transfer Time 20 - 50 µs, and Timing set to 50% for both. For MS/MS only the collision energy ranges from 100% - 250% with timing set to 50% for both. Auto MS/MS was used with a predefined Cycle Time of 0.5 s, Active Exclusion was used with Exclusion after 3 Spectra, and a Release time set to 0.15 min. Dynamic MS/MS spectra acquisition was applied with a target intensity of 20 000 counts, max MS/MS spectra acquisition of 30 Hz (0.03 sec), and min MS/MS spectra acquisition of 16 Hz (0.06 sec). Sodium formate clusters were applied for instrument mass calibration and for internal recalibration of individual samples. A Precursor Exclusion list was used with Exclusion of mass range of 20-60.

*Metabolomics preprocessing*

Bruker .d files were exported to the .mzML format using ProteoWizard's MSConvert10 and subsequently preprocessed using the Ion Identity Network workflow in MZmine[1,2] (version 3.3.0). Mass lists were created by considering mass spectra with retention times of 0.4-14 minutes and retaining MS1 intensities above 3E2 and MS2 intensities above 0. The chromatogram was built through the ADAP chromatogram builder by using the following parameters, minimum group size of scans: 5, group intensity threshold: 3E2, minimum highest intensity: 9.0E2, and *m/z* tolerance: 0.002 *m/z* or 5 ppm. The chromatogram was smoothed with a filter width of 5. The local minimum search algorithm was used for deconvolution with parameters set to, chromatographic threshold: 85%, minimum RT range (min): 0.05, minimum relative height: 0%, minimum absolute height: 9.0E2, min ratio of peak top/edge: 2.2, peak duration range (min): 0.05-0.5. The peaks were deisotoped by using the isotopic peak grouper function, with parameters set to, *m/z* tolerance: 0.002 *m/z* or 5 ppm, retention time tolerance: 0.15 min, monotonic shape: on, maximum charge: 2, representative isotope: most intense. Peaks from all samples were aligned, by using the join aligner function with parameters set to, *m/z* tolerance: 0.002 *m/z* or 5 ppm, retention time tolerance: 0.15 min, weight for *m/z*: 75, weight for retention time: 25. Rows were then filtered using the duplicate peak filter with the new average filter mode and *m/z* tolerance set to 0.001 *m/z* or 5 ppm and RT tolerance 0.03 min. Gap-filling was performed using the same mz and RT range gap filler, with a *m/z* tolerance of 0.002 *m/z* or 5ppm and an RT tolerance of 0.03 minutes. The metaCorrelate function was used to find correlating peak shapes with parameters set to, RT tolerance: 0.1 min, min. height: 9.0E2, noise level: 5E2, min samples in all: 2 (abs), min samples in group: 0 (abs), min %-intensity overlap: 60%, exclude estimated features (gap-filled): on. Parameters for the correlation grouping were set as follows, min data points: 5, min data points on edge: 2, measure: Pearson, min feature shape correlation: 85%. Ion identity networking parameters were set to, *m/z* tolerance: 0.002 *m/z* or 5 ppm, check: one feature, min. height: 9.0E2E3 with ion identity library parameters set to, MS mode: positive, maximum charge: 2, maximum molecules/cluster: 2, adducts: M+H, M+Na, M+K, modifications: M-H2O, M-NH3. Further ion identity networks were added with *m/z* tolerance: 0.002 *m/z* or 5 ppm, min. height: 9.0E2, and ion identity library parameters set to, MS mode: positive, maximum charge: 2, maximum molecules/cluster: 6, adducts: M+H, M+Na, modifications: M-H2O, M-2H2O, M-3H2O, M-4H2O, M-5H2O, and *m/z* tolerance: 0.002 *m/z* or 5 ppm, min. height: 1E3, and annotation refinement on with parameters set to, delete smaller networks: link threshold: 4, delete networks without monomer: on, and ion identity library parameters set to MS mode: positive, maximum charge: 2, maximum molecules/cluster: 2, adducts: M+H, M+Na, M+K, modifications: M-H2O, M-NH3. Finally, two feature tables were exported in the .csv format. One feature table contains all extracted mass spectral features, and another feature table filtered for mass spectral features with associated fragmentation spectra (MS2). An aggregated list of MS2 fragmentation spectra was exported in the .mgf format and submitted to ion identity feature-based mass spectral molecular networking through the Global Natural Products Social Molecular Networking Platform (GNPS)[3,4].

Before statistical analysis, connected ion adducts were merged and mass spectral feature signals with a relative intensity less than 5 times the mean relative intensity in all paper blank samples were removed. Metabolite features present in less than 25% of the samples were removed and features present in fewer than 75% were treated as binary variables (present or absent). This resulted in a final dataset with a total of 1076 metabolite features measured, among which 433 features were continuous and 643 were binary variables. Missing values for metabolite features with continuous measurements were further subjected to imputation and batch correction procedures. Among the 433 metabolite features, 214 (49%) had less than 5% missing values. Missing values were imputed using missForest[5], with the maximum number of iterations set to 10 and the number of trees to 100. Batch correction was performed by centering and univariance scaling of each metabolite per batch.

*Quality control procedures*

Quality control procedures for metabolite profiling are divided into three main categories, system suitability test (SST), batch evaluation, and post-processing quality control.

In the SST, the mass spectral and chromatographic performance was evaluated prior to each batch by injecting two different standard samples. Standard sample A consisted of leucine enkephalin (1.8 µM in 50/50: H2O/ACN) and standard sample B consisted of a mix of amino acids and acylcarnitines in 50/50: H2O/ACN (Cambridge Isotope Laboratories, Tewksbury, MA, USA). System suitability was evaluated based on retention time deviation (<0.2 min), mass accuracy (<2 ppm), and relative standard deviation (<20%) for all compounds in both standard samples A and B. Batch evaluation was performed by monitoring sixteen quality control metabolites in pooled sample extracts, EC samples and paper blanks. Potential carry-over is controlled by ensuring that quality control metabolites are not present in the paper-blank samples. Mass spectral and chromatographic performance is evaluated by monitoring retention time deviation (<0.2 min), mass accuracy (<2 ppm), and coefficient of variation (<20%) in EC samples and pooled sample extracts. Data for batch evaluation is presented in **Table S13**. Feature picking for the SST and batch evaluation was performed in Metaboscape (Bruker, Billerica, MA, United States).

*Metabolite identification*

To annotate mass spectral features to putative chemical structures, a mass spectral molecular network was created through the GNPS Platform (http://gnps.ucsd.edu) using the ion identity feature-based molecular networking workflow (https://ccms-ucsd.github.io/GNPSDocumentation/fbmn-iin/)[1,3,4]. The data was filtered by removing all MS/MS fragment ions within +/- 17 Da of the precursor *m/z*. MS/MS spectra were window-filtered by choosing only the top 6 fragment ions in the +/- 50 Da window throughout the spectrum. The precursor ion mass tolerance was set to 0.02 Da and an MS/MS fragment ion tolerance of 0.02 Da. A network was then created where edges were filtered to have a cosine score above 0.7 and more than 4 matched peaks. Further, edges between two nodes were kept in the network if and only if each of the nodes appeared in each other's respective top 10 most similar nodes. Finally, the maximum size of a molecular family was set to 100, and the lowest-scoring edges were removed from molecular families until the molecular family size was below this threshold. The spectra in the network were then searched against all GNPS' spectral libraries. The library spectra were filtered in the same manner as the input data. All matches kept between network spectra and library spectra were required to have a score above 0.7 and at least 4 matched peaks. Furthermore, *in silico* structural annotation prediction was performed using Sirius+CSI:FingerID[6]. Chemical class annotations were performed using deep neural networks in CANOPUS[7] and followed the ClassyFire chemical ontology[8].

### Supplementary Tables

See **Extended Data**

### Supplementary Figures

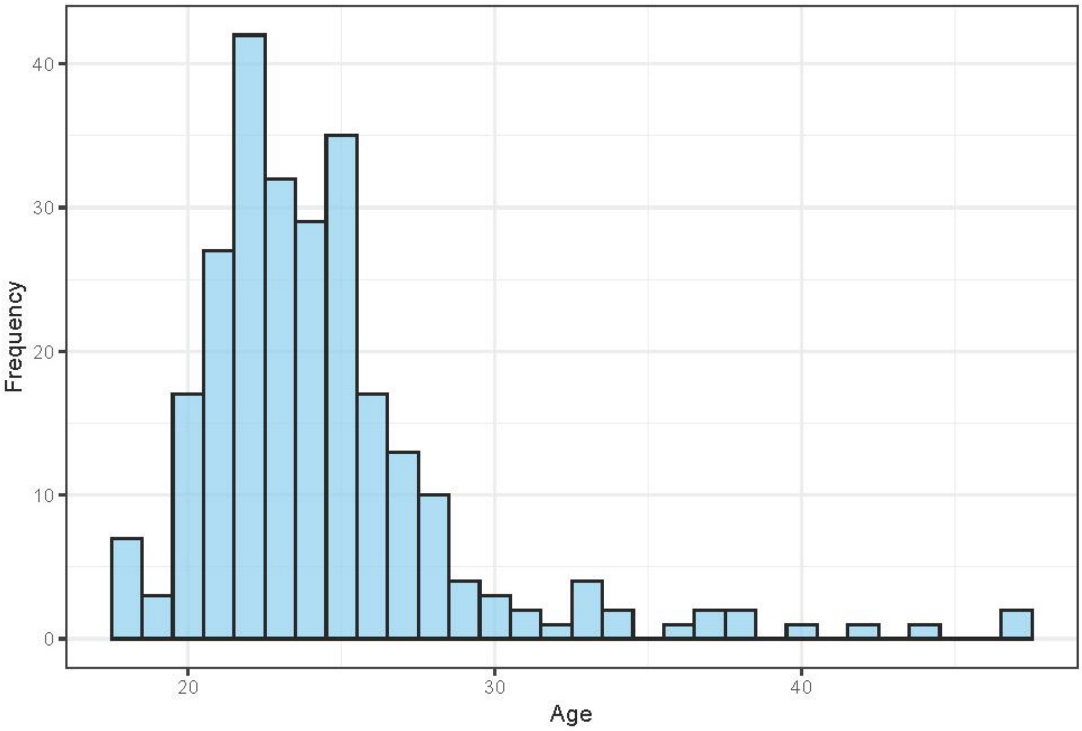

**Figure S1** | Age distribution of the sample. Histogram showing the age distribution of the study participants. The x-axis represents age in years, and the y-axis shows the frequency of individuals within each age group.

A

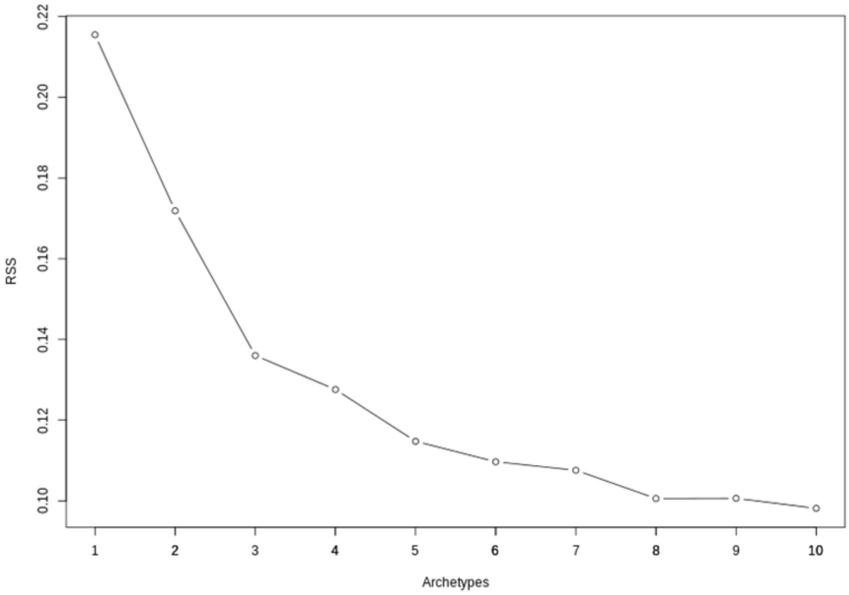

B

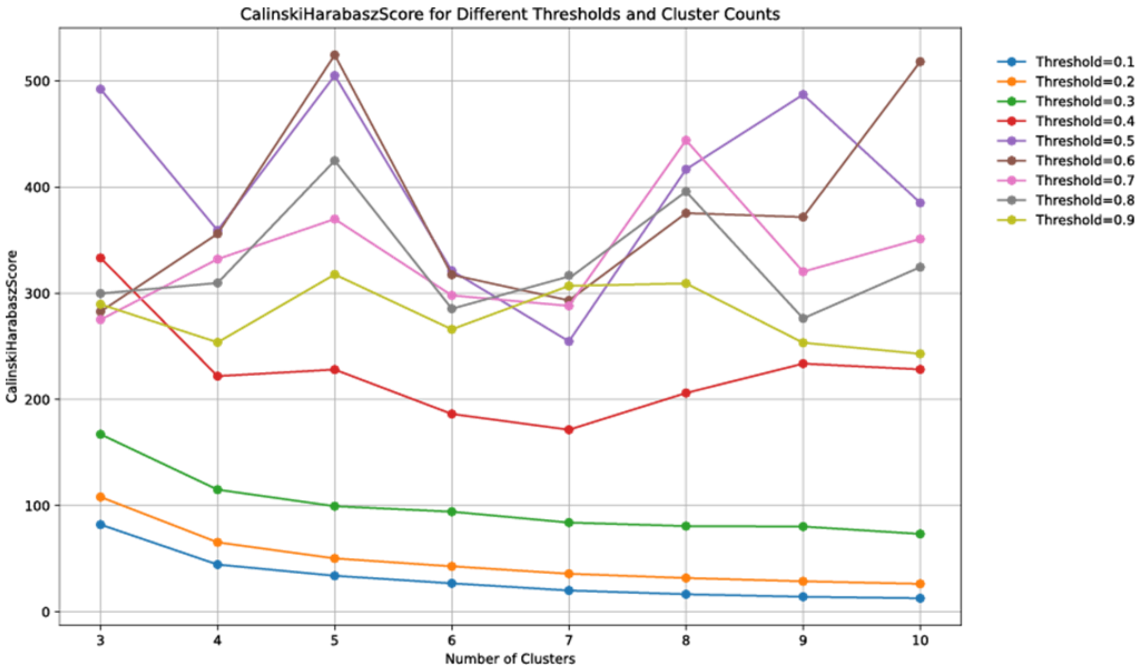

**Figure S2 |** Model Selection and Cluster Validation for Archetype Analysis. **(A)** Scree plot of the minimized residual sum of squares (RSS) for k archetypes (k = 1 to 10) across 100 restarts of the archetype algorithm. The plot shows a plateau in intra-cluster variance reduction at k = 5. **(B)** The Calinski-Harabasz index supports this, indicating similar model fitness at thresholds of 0.5 and 0.6.

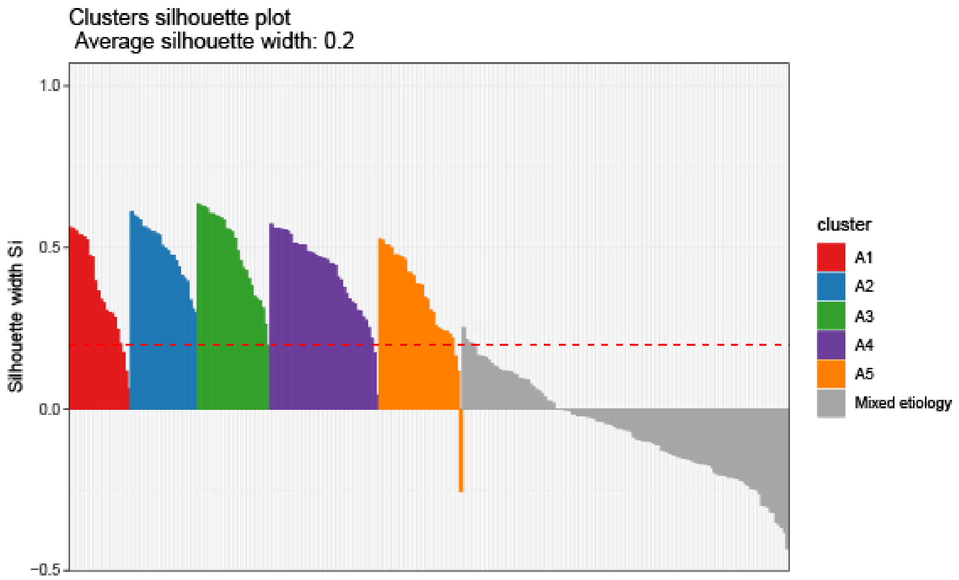

**Figure S3** | Silhouette Analysis of Clustering for Discrete Archetype Memberships. Silhouette analysis of clustering for individuals with discrete archetype and the mixed archetype group. The analysis reveals that individuals with discrete archetype (greater than 0.5) for each of the five archetypes are well-clustered, whereas the mixed archetype group does not form a cohesive cluster.

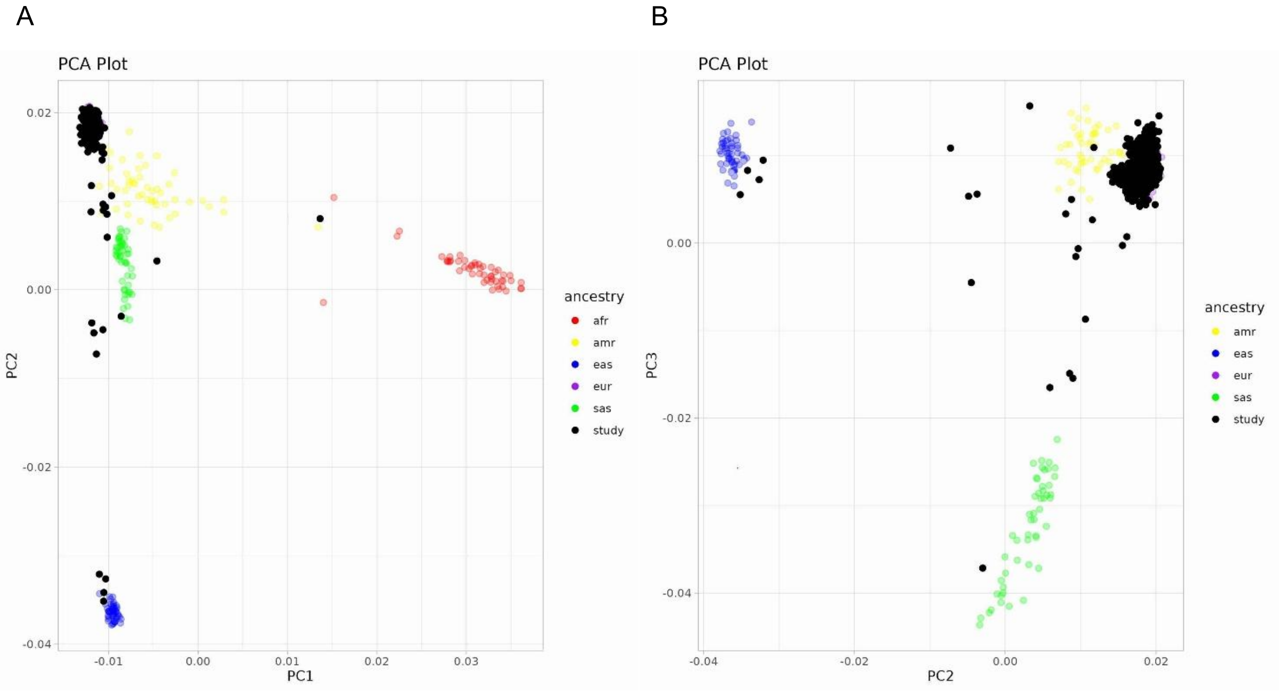

**Figure S4** | Principal Component Analysis of Genetic Populations. Plots of the first three principal components from LDAK (**A**: PC1 and PC2, **B**: PC2 and PC3). Solid black circles represent 265 individuals from the Danish subset of CA18106 that donated DNA for the study. Samples from the 1000 genome project belonging to various genetic populations are presented with different colored circles: red = African (AFR), yellow = Ad Mixed American (AMR), blue = East Asian (EAS), purple = European (EUR), and green = South Asian (SAS).

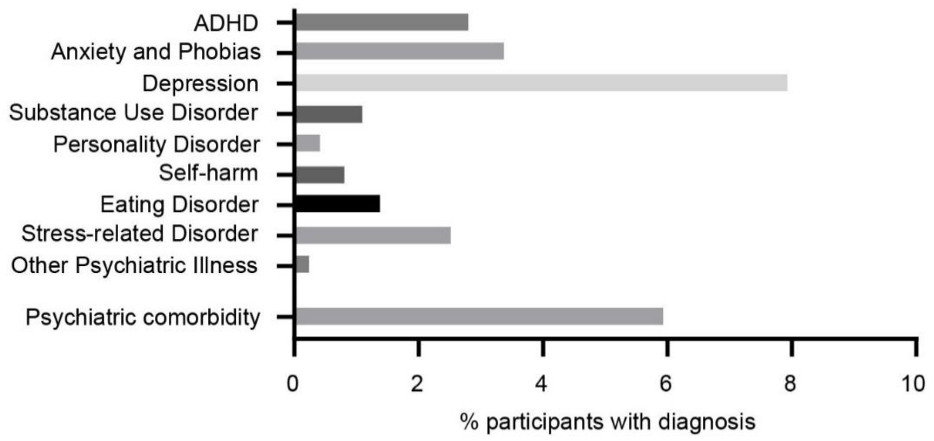

**Figure S5 |** Prevalence of self-reported MDx diagnoses within the sample. The y-axis represents different MDx diagnoses, while the x-axis shows the proportion or number of individuals reporting each diagnosis.

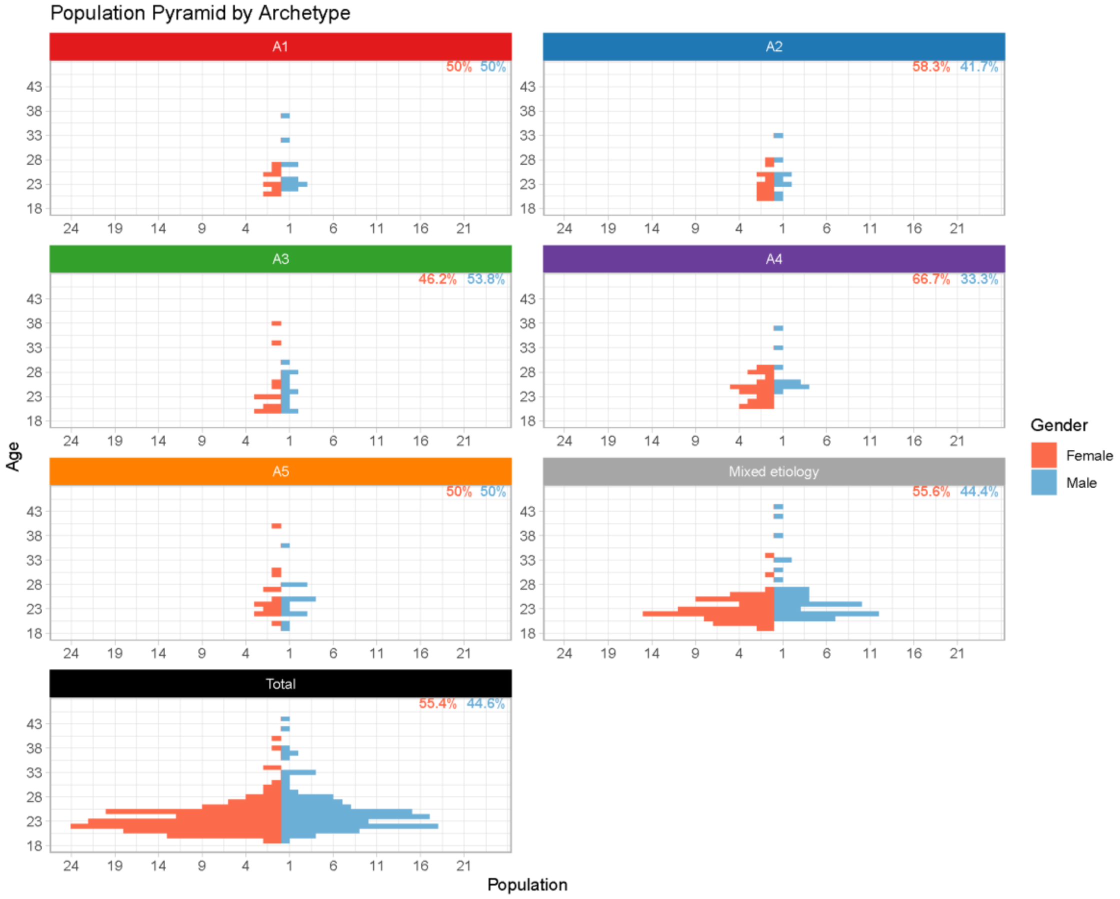

**Figure S6** | Population Pyramid of Age Distribution by Discrete Archetype and Gender. This population pyramid displays the age distribution of males and females across each discrete archetype, as well as for the total cohort. The y-axis represents age groups, and the x-axis shows the number of individuals, separated by gender.

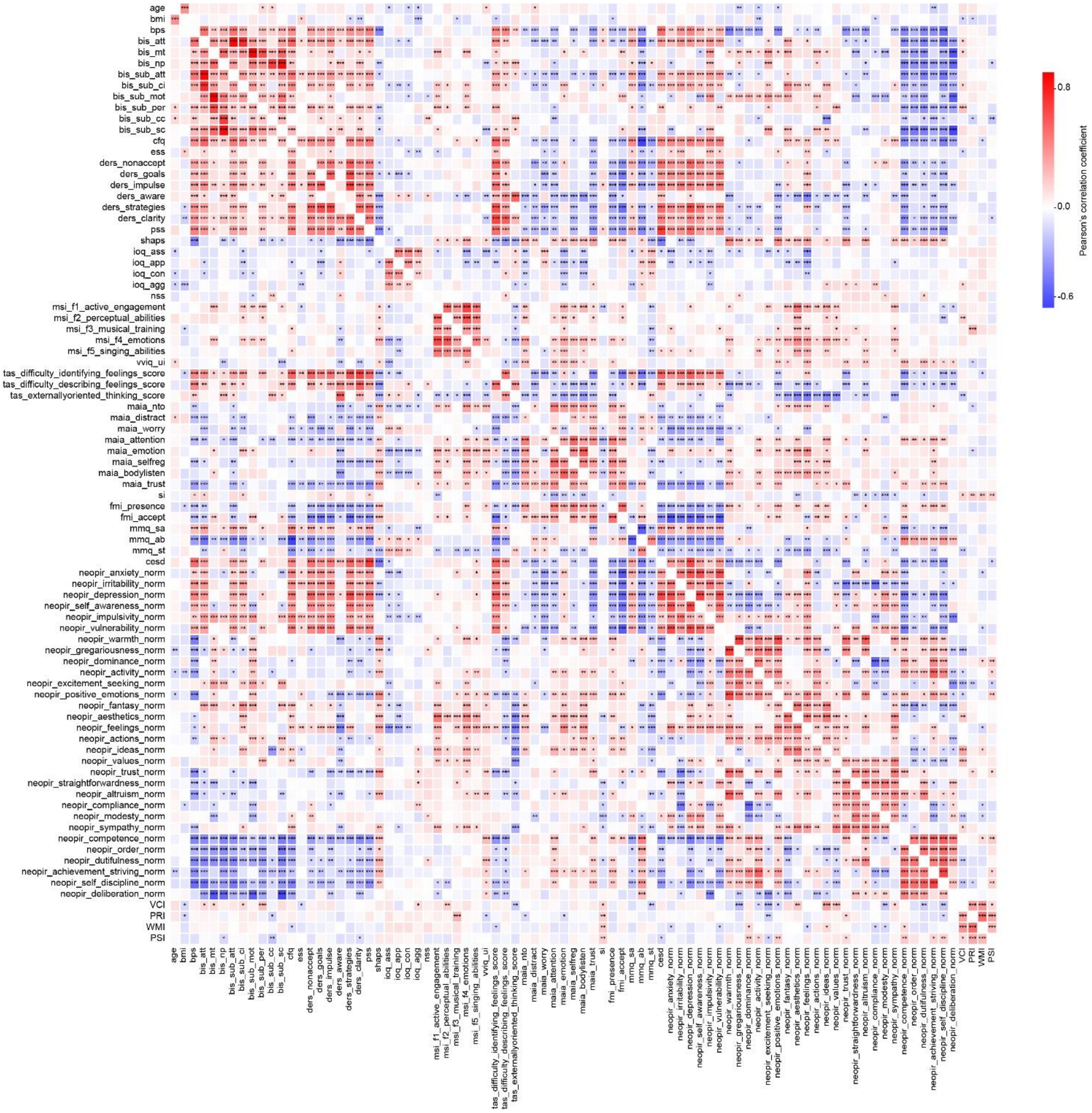

**Figure S7 |** Correlation Heatmap of Psychometric Variables. This heatmap displays Pearson's correlation coefficients between psychometric variables, with shades of red and blue indicating the strength and direction of the correlations. Significant correlations are marked with asterisks: * for p<0.05, ** for p<0.01, and *** for p<0.001. Darker shades represent stronger correlations, with red indicating positive correlations and blue indicating negative correlations.

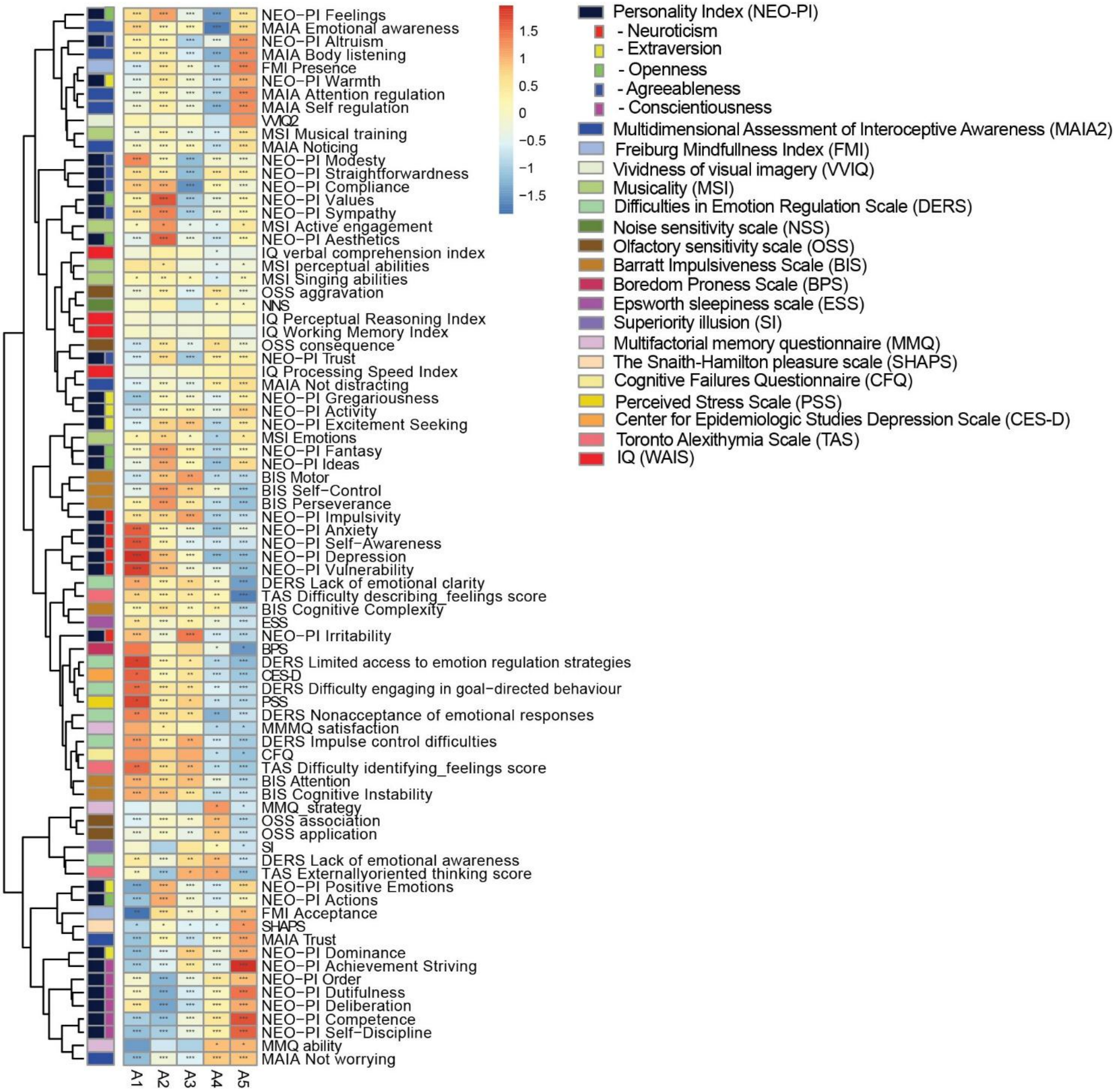

**Figure S8 |** Heatmap of associations between archetype scores and clustering variables. This heatmap shows the associations between archetype scores and clustering variables, with shades of red and blue indicating the strength and direction of the associations. Darker shades of red represent stronger positive associations, while darker shades of blue indicate stronger negative associations. The clustering variables are indicated in the legend and are color-coded to match the heatmap. Significant correlations are marked with asterisks: * for p<0.05, ** for p<0.01, and *** for p<0.001.

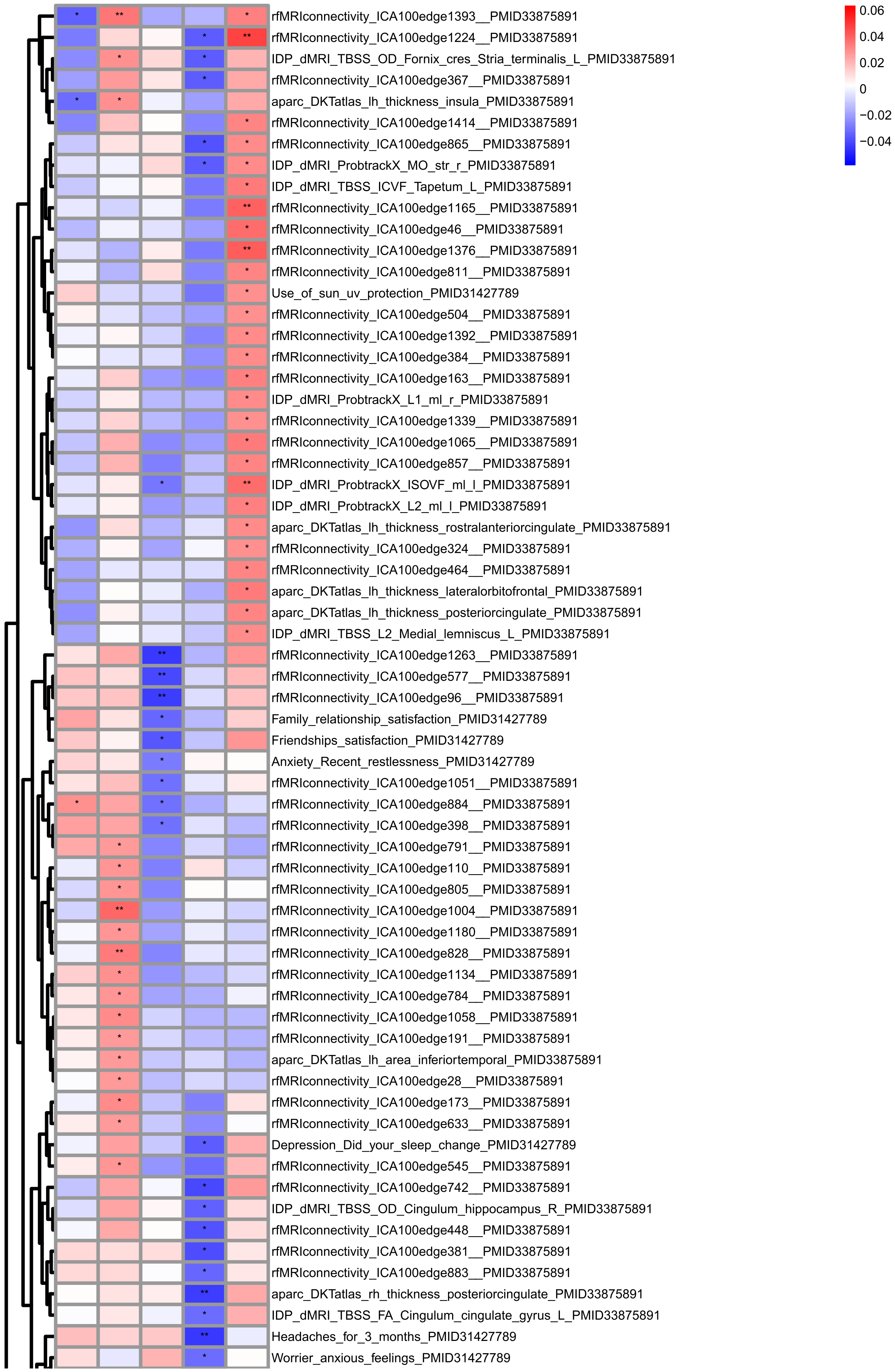
0.06
0.04
0.02
0
−0.02
−0.04
rfMRIconnectivity_ICA100edge1393__PMID33875891
rfMRIconnectivity_ICA100edge1224__PMID33875891
IDP_dMRI_TBSS_OD_Fornix_cres_Stria_terminalis_L_PMID33875891
rfMRIconnectivity_ICA100edge367__PMID33875891
aparc_DKTatlas_lh_thickness_insula_PMID33875891
rfMRIconnectivity_ICA100edge1414__PMID33875891
rfMRIconnectivity_ICA100edge865__PMID33875891
IDP_dMRI_ProbtrackX_MO_str_r_PMID33875891
IDP_dMRI_TBSS_ICVF_Tapetum_L_PMID33875891
rfMRIconnectivity_ICA100edge1165__PMID33875891
rfMRIconnectivity_ICA100edge46__PMID33875891
rfMRIconnectivity_ICA100edge1376__PMID33875891
rfMRIconnectivity_ICA100edge811__PMID33875891
Use_of_sun_uv_protection_PMID31427789
rfMRIconnectivity_ICA100edge504__PMID33875891
rfMRIconnectivity_ICA100edge1392__PMID33875891
rfMRIconnectivity_ICA100edge384__PMID33875891
rfMRIconnectivity_ICA100edge163__PMID33875891
IDP_dMRI_ProbtrackX_L1_ml_r_PMID33875891
rfMRIconnectivity_ICA100edge1339__PMID33875891
rfMRIconnectivity_ICA100edge1065__PMID33875891
rfMRIconnectivity_ICA100edge857__PMID33875891
IDP_dMRI_ProbtrackX_ISOVF_ml_l_PMID33875891
IDP_dMRI_ProbtrackX_L2_ml_l_PMID33875891
aparc_DKTatlas_lh_thickness_rostralanteriorcingulate_PMID33875891
rfMRIconnectivity_ICA100edge324__PMID33875891
rfMRIconnectivity_ICA100edge464__PMID33875891
aparc_DKTatlas_lh_thickness_lateralorbitofrontal_PMID33875891
aparc_DKTatlas_lh_thickness_posteriorcingulate_PMID33875891
IDP_dMRI_TBSS_L2_Medial_lemniscus_L_PMID33875891
rfMRIconnectivity_ICA100edge1263__PMID33875891
rfMRIconnectivity_ICA100edge577__PMID33875891
rfMRIconnectivity_ICA100edge96__PMID33875891
Family_relationship_satisfaction_PMID31427789
Friendships_satisfaction_PMID31427789
Anxiety_Recent_restlessness_PMID31427789
rfMRIconnectivity_ICA100edge1051__PMID33875891
rfMRIconnectivity_ICA100edge884__PMID33875891
rfMRIconnectivity_ICA100edge398__PMID33875891
rfMRIconnectivity_ICA100edge791__PMID33875891
rfMRIconnectivity_ICA100edge110__PMID33875891
rfMRIconnectivity_ICA100edge805__PMID33875891
rfMRIconnectivity_ICA100edge1004__PMID33875891
rfMRIconnectivity_ICA100edge1180__PMID33875891
rfMRIconnectivity_ICA100edge828__PMID33875891
rfMRIconnectivity_ICA100edge1134__PMID33875891
rfMRIconnectivity_ICA100edge784__PMID33875891
rfMRIconnectivity_ICA100edge1058__PMID33875891
rfMRIconnectivity_ICA100edge191__PMID33875891
aparc_DKTatlas_lh_area_inferiortemporal_PMID33875891
rfMRIconnectivity_ICA100edge28__PMID33875891
rfMRIconnectivity_ICA100edge173__PMID33875891
rfMRIconnectivity_ICA100edge633__PMID33875891
Depression_Did_your_sleep_change_PMID31427789
rfMRIconnectivity_ICA100edge545__PMID33875891
rfMRIconnectivity_ICA100edge742__PMID33875891
IDP_dMRI_TBSS_OD_Cingulum_hippocampus_R_PMID33875891
rfMRIconnectivity_ICA100edge448__PMID33875891
rfMRIconnectivity_ICA100edge381__PMID33875891
rfMRIconnectivity_ICA100edge883__PMID33875891
aparc_DKTatlas_rh_thickness_posteriorcingulate_PMID33875891
IDP_dMRI_TBSS_FA_Cingulum_cingulate_gyrus_L_PMID33875891
Headaches_for_3_months_PMID31427789
Worrier_anxious_feelings_PMID31427789

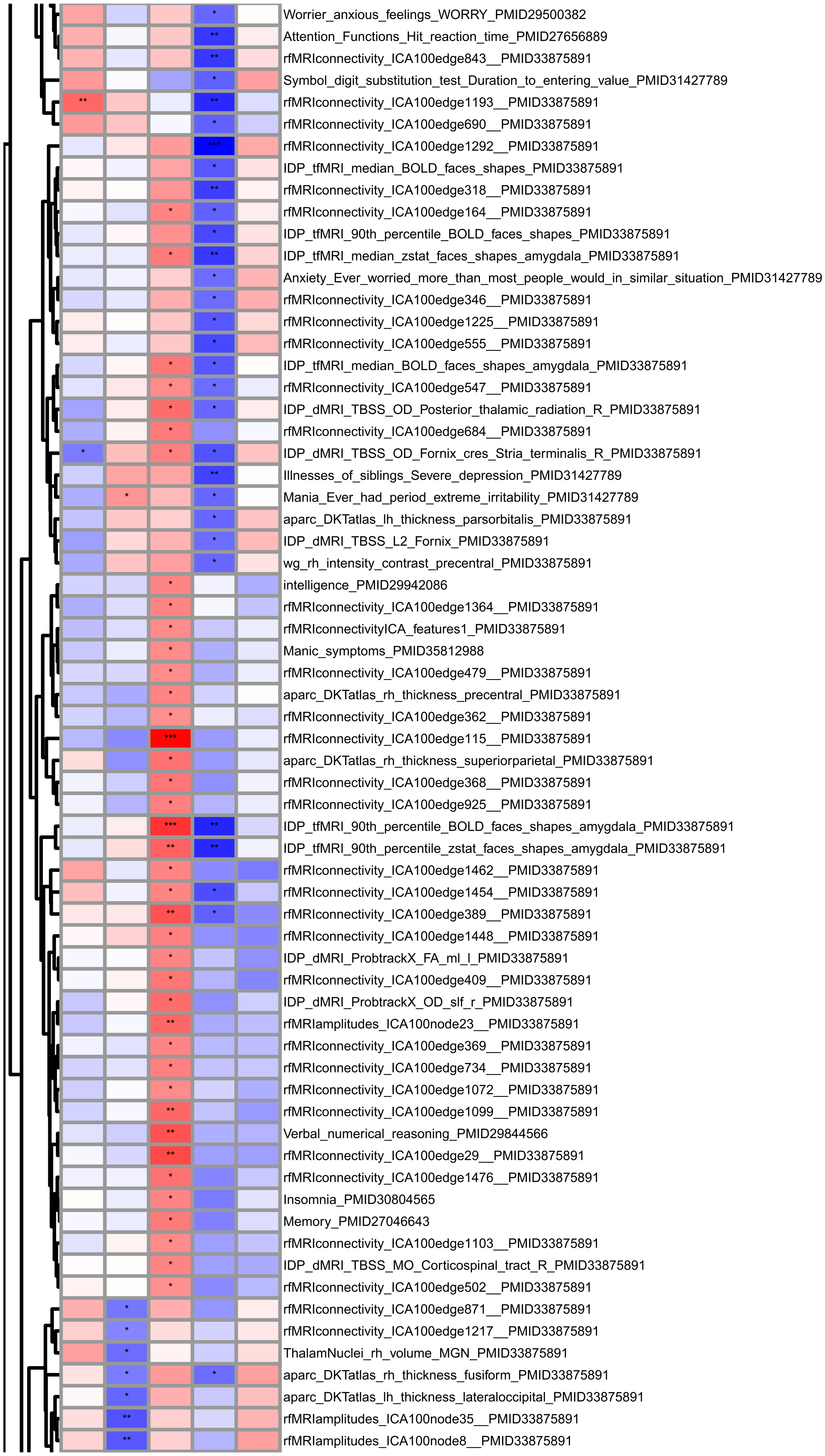
Worrier_anxious_feelings_WORRY_PMID29500382
Attention_Functions_Hit_reaction_time_PMID27656889
rfMRIconnectivity_ICA100edge843__PMID33875891
Symbol_digit_substitution_test_Duration_to_entering_value_PMID31427789
rfMRIconnectivity_ICA100edge1193__PMID33875891
rfMRIconnectivity_ICA100edge690__PMID33875891
rfMRIconnectivity_ICA100edge1292__PMID33875891
IDP_tfMRI_median_BOLD_faces_shapes_PMID33875891
rfMRIconnectivity_ICA100edge318__PMID33875891
rfMRIconnectivity_ICA100edge164__PMID33875891
IDP_tfMRI_90th_percentile_BOLD_faces_shapes_PMID33875891
IDP_tfMRI_median_zstat_faces_shapes_amygdala_PMID33875891
Anxiety_Ever_worried_more_than_most_people_would_in_similar_situation_PMID31427789
rfMRIconnectivity_ICA100edge346__PMID33875891
rfMRIconnectivity_ICA100edge1225__PMID33875891
rfMRIconnectivity_ICA100edge555__PMID33875891
IDP_tfMRI_median_BOLD_faces_shapes_amygdala_PMID33875891
rfMRIconnectivity_ICA100edge547__PMID33875891
IDP_dMRI_TBSS_OD_Posterior_thalamic_radiation_R_PMID33875891
rfMRIconnectivity_ICA100edge684__PMID33875891
IDP_dMRI_TBSS_OD_Fornix_cres_Stria_terminalis_R_PMID33875891
Illnesses_of_siblings_Severe_depression_PMID31427789
Mania_Ever_had_period_extreme_irritability_PMID31427789
aparc_DKTatlas_lh_thickness_parsorbitalis_PMID33875891
IDP_dMRI_TBSS_L2_Fornix_PMID33875891
wg_rh_intensity_contrast_precentral_PMID33875891
intelligence_PMID29942086
rfMRIconnectivity_ICA100edge1364__PMID33875891
rfMRIconnectivityICA_features1_PMID33875891
Manic_symptoms_PMID35812988
rfMRIconnectivity_ICA100edge479__PMID33875891
aparc_DKTatlas_rh_thickness_precentral_PMID33875891
rfMRIconnectivity_ICA100edge362__PMID33875891
rfMRIconnectivity_ICA100edge115__PMID33875891
aparc_DKTatlas_rh_thickness_superiorparietal_PMID33875891
rfMRIconnectivity_ICA100edge368__PMID33875891
rfMRIconnectivity_ICA100edge925__PMID33875891
IDP_tfMRI_90th_percentile_BOLD_faces_shapes_amygdala_PMID33875891
IDP_tfMRI_90th_percentile_zstat_faces_shapes_amygdala_PMID33875891
rfMRIconnectivity_ICA100edge1462__PMID33875891
rfMRIconnectivity_ICA100edge1454__PMID33875891
rfMRIconnectivity_ICA100edge389__PMID33875891
rfMRIconnectivity_ICA100edge1448__PMID33875891
IDP_dMRI_ProbtrackX_FA_ml_l_PMID33875891
rfMRIconnectivity_ICA100edge409__PMID33875891
IDP_dMRI_ProbtrackX_OD_slf_r_PMID33875891
rfMRIamplitudes_ICA100node23__PMID33875891
rfMRIconnectivity_ICA100edge369__PMID33875891
rfMRIconnectivity_ICA100edge734__PMID33875891
rfMRIconnectivity_ICA100edge1072__PMID33875891
rfMRIconnectivity_ICA100edge1099__PMID33875891
Verbal_numerical_reasoning_PMID29844566
rfMRIconnectivity_ICA100edge29__PMID33875891
rfMRIconnectivity_ICA100edge1476__PMID33875891
Insomnia_PMID30804565
Memory_PMID27046643
rfMRIconnectivity_ICA100edge1103__PMID33875891
IDP_dMRI_TBSS_MO_Corticospinal_tract_R_PMID33875891
rfMRIconnectivity_ICA100edge502__PMID33875891
rfMRIconnectivity_ICA100edge871__PMID33875891
rfMRIconnectivity_ICA100edge1217__PMID33875891
ThalamNuclei_rh_volume_MGN_PMID33875891
aparc_DKTatlas_rh_thickness_fusiform_PMID33875891
aparc_DKTatlas_lh_thickness_lateraloccipital_PMID33875891
rfMRIamplitudes_ICA100node35__PMID33875891
rfMRIamplitudes_ICA100node8__PMID33875891

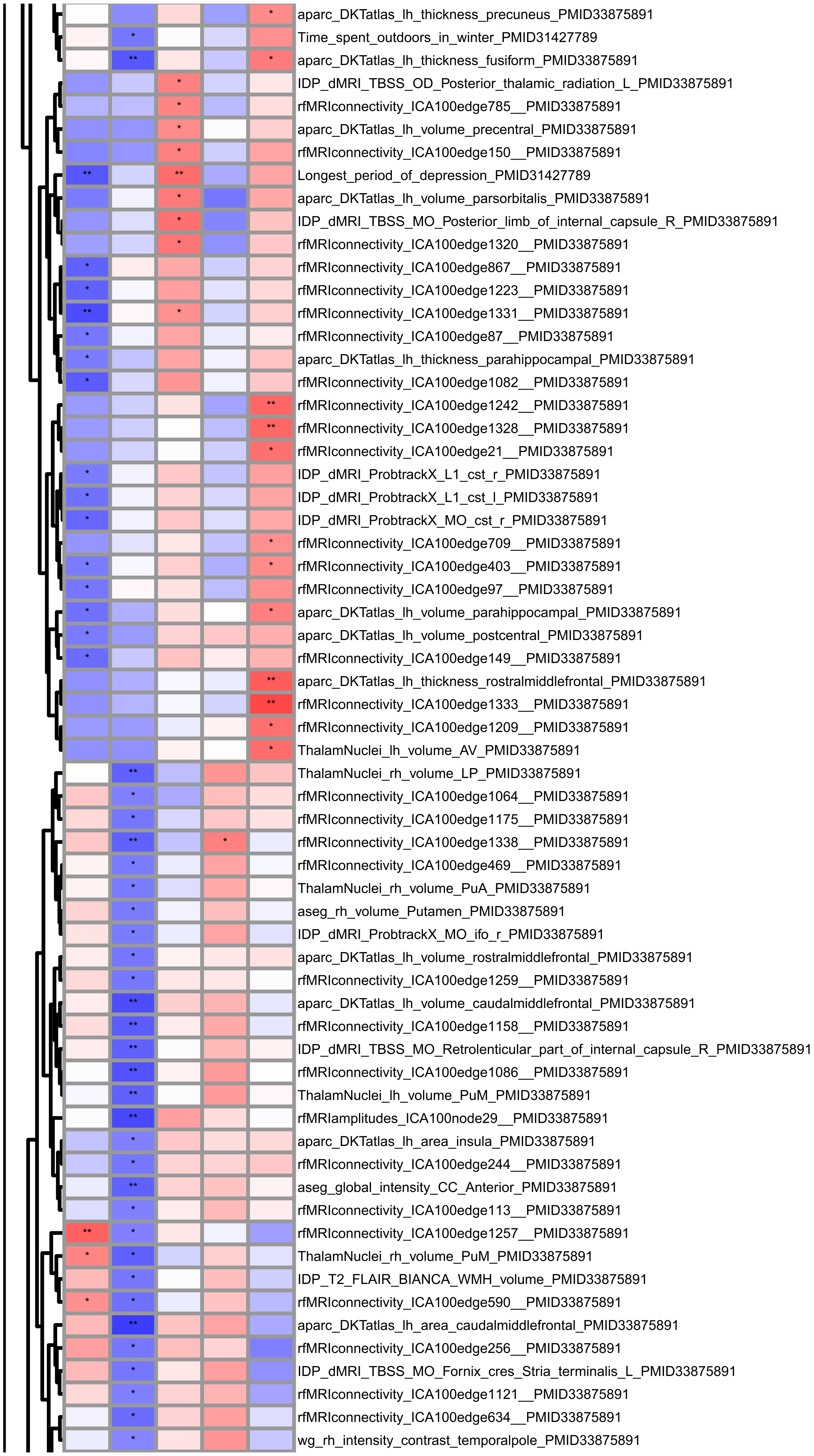
aparc_DKTatlas_lh_thickness_precuneus_PMID33875891
Time_spent_outdoors_in_winter_PMID31427789
aparc_DKTatlas_lh_thickness_fusiform_PMID33875891
IDP_dMRI_TBSS_OD_Posterior_thalamic_radiation_L_PMID33875891
rfMRIconnectivity_ICA100edge785__PMID33875891
aparc_DKTatlas_lh_volume_precentral_PMID33875891
rfMRIconnectivity_ICA100edge150__PMID33875891
Longest_period_of_depression_PMID31427789
aparc_DKTatlas_lh_volume_parsorbitalis_PMID33875891
IDP_dMRI_TBSS_MO_Posterior_limb_of_internal_capsule_R_PMID33875891
rfMRIconnectivity_ICA100edge1320__PMID33875891
rfMRIconnectivity_ICA100edge867__PMID33875891
rfMRIconnectivity_ICA100edge1223__PMID33875891
rfMRIconnectivity_ICA100edge1331__PMID33875891
rfMRIconnectivity_ICA100edge87__PMID33875891
aparc_DKTatlas_lh_thickness_parahippocampal_PMID33875891
rfMRIconnectivity_ICA100edge1082__PMID33875891
rfMRIconnectivity_ICA100edge1242__PMID33875891
rfMRIconnectivity_ICA100edge1328__PMID33875891
rfMRIconnectivity_ICA100edge21__PMID33875891
IDP_dMRI_ProbtrackX_L1_cst_r_PMID33875891
IDP_dMRI_ProbtrackX_L1_cst_l_PMID33875891
IDP_dMRI_ProbtrackX_MO_cst_r_PMID33875891
rfMRIconnectivity_ICA100edge709__PMID33875891
rfMRIconnectivity_ICA100edge403__PMID33875891
rfMRIconnectivity_ICA100edge97__PMID33875891
aparc_DKTatlas_lh_volume_parahippocampal_PMID33875891
aparc_DKTatlas_lh_volume_postcentral_PMID33875891
rfMRIconnectivity_ICA100edge149__PMID33875891
aparc_DKTatlas_lh_thickness_rostralmiddlefrontal_PMID33875891
rfMRIconnectivity_ICA100edge1333__PMID33875891
rfMRIconnectivity_ICA100edge1209__PMID33875891
ThalamNuclei_lh_volume_AV_PMID33875891
ThalamNuclei_rh_volume_LP_PMID33875891
rfMRIconnectivity_ICA100edge1064__PMID33875891
rfMRIconnectivity_ICA100edge1175__PMID33875891
rfMRIconnectivity_ICA100edge1338__PMID33875891
rfMRIconnectivity_ICA100edge469__PMID33875891
ThalamNuclei_rh_volume_PuA_PMID33875891
aseg_rh_volume_Putamen_PMID33875891
IDP_dMRI_ProbtrackX_MO_ifo_r_PMID33875891
aparc_DKTatlas_lh_volume_rostralmiddlefrontal_PMID33875891
rfMRIconnectivity_ICA100edge1259__PMID33875891
aparc_DKTatlas_lh_volume_caudalmiddlefrontal_PMID33875891
rfMRIconnectivity_ICA100edge1158__PMID33875891
IDP_dMRI_TBSS_MO_Retrolenticular_part_of_internal_capsule_R_PMID33875891
rfMRIconnectivity_ICA100edge1086__PMID33875891
ThalamNuclei_lh_volume_PuM_PMID33875891
rfMRIamplitudes_ICA100node29__PMID33875891
aparc_DKTatlas_lh_area_insula_PMID33875891
rfMRIconnectivity_ICA100edge244__PMID33875891
aseg_global_intensity_CC_Anterior_PMID33875891
rfMRIconnectivity_ICA100edge113__PMID33875891
rfMRIconnectivity_ICA100edge1257__PMID33875891
ThalamNuclei_rh_volume_PuM_PMID33875891
IDP_T2_FLAIR_BIANCA_WMH_volume_PMID33875891
rfMRIconnectivity_ICA100edge590__PMID33875891
aparc_DKTatlas_lh_area_caudalmiddlefrontal_PMID33875891
rfMRIconnectivity_ICA100edge256__PMID33875891
IDP_dMRI_TBSS_MO_Fornix_cres_Stria_terminalis_L_PMID33875891
rfMRIconnectivity_ICA100edge1121__PMID33875891
rfMRIconnectivity_ICA100edge634__PMID33875891
wg_rh_intensity_contrast_temporalpole_PMID33875891

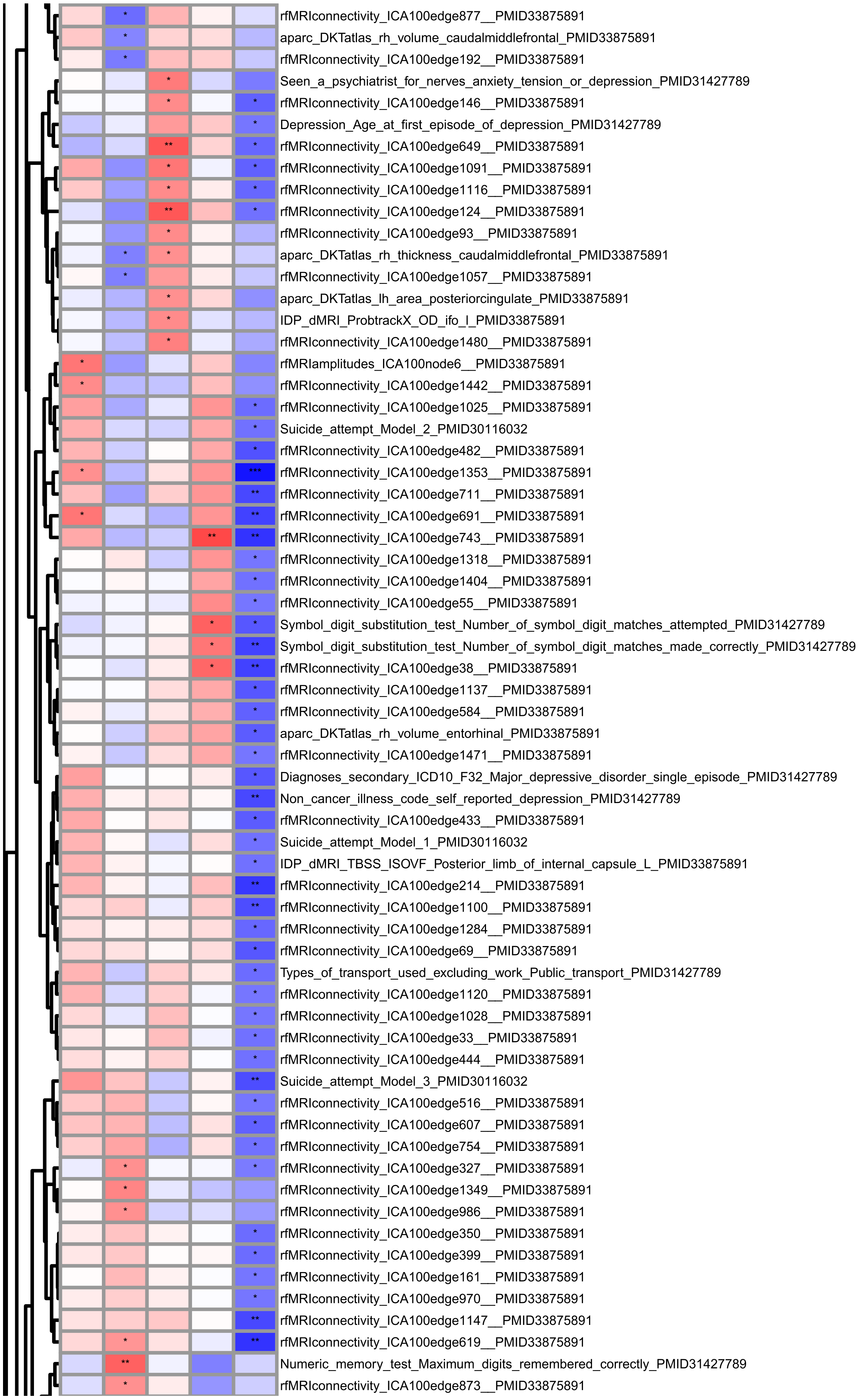
rfMRIconnectivity_ICA100edge877__PMID33875891
aparc_DKTatlas_rh_volume_caudalmiddlefrontal_PMID33875891
rfMRIconnectivity_ICA100edge192__PMID33875891
Seen_a_psychiatrist_for_nerves_anxiety_tension_or_depression_PMID31427789
rfMRIconnectivity_ICA100edge146__PMID33875891
Depression_Age_at_first_episode_of_depression_PMID31427789
rfMRIconnectivity_ICA100edge649__PMID33875891
rfMRIconnectivity_ICA100edge1091__PMID33875891
rfMRIconnectivity_ICA100edge1116__PMID33875891
rfMRIconnectivity_ICA100edge124__PMID33875891
rfMRIconnectivity_ICA100edge93__PMID33875891
aparc_DKTatlas_rh_thickness_caudalmiddlefrontal_PMID33875891
rfMRIconnectivity_ICA100edge1057__PMID33875891
aparc_DKTatlas_lh_area_posteriorcingulate_PMID33875891
IDP_dMRI_ProbtrackX_OD_ifo_l_PMID33875891
rfMRIconnectivity_ICA100edge1480__PMID33875891
rfMRIamplitudes_ICA100node6__PMID33875891
rfMRIconnectivity_ICA100edge1442__PMID33875891
rfMRIconnectivity_ICA100edge1025__PMID33875891
Suicide_attempt_Model_2_PMID30116032
rfMRIconnectivity_ICA100edge482__PMID33875891
rfMRIconnectivity_ICA100edge1353__PMID33875891
rfMRIconnectivity_ICA100edge711__PMID33875891
rfMRIconnectivity_ICA100edge691__PMID33875891
rfMRIconnectivity_ICA100edge743__PMID33875891
rfMRIconnectivity_ICA100edge1318__PMID33875891
rfMRIconnectivity_ICA100edge1404__PMID33875891
rfMRIconnectivity_ICA100edge55__PMID33875891
Symbol_digit_substitution_test_Number_of_symbol_digit_matches_attempted_PMID31427789
Symbol_digit_substitution_test_Number_of_symbol_digit_matches_made_correctly_PMID31427789
rfMRIconnectivity_ICA100edge38__PMID33875891
rfMRIconnectivity_ICA100edge1137__PMID33875891
rfMRIconnectivity_ICA100edge584__PMID33875891
aparc_DKTatlas_rh_volume_entorhinal_PMID33875891
rfMRIconnectivity_ICA100edge1471__PMID33875891
Diagnoses_secondary_ICD10_F32_Major_depressive_disorder_single_episode_PMID31427789
Non_cancer_illness_code_self_reported_depression_PMID31427789
rfMRIconnectivity_ICA100edge433__PMID33875891
Suicide_attempt_Model_1_PMID30116032
IDP_dMRI_TBSS_ISOVF_Posterior_limb_of_internal_capsule_L_PMID33875891
rfMRIconnectivity_ICA100edge214__PMID33875891
rfMRIconnectivity_ICA100edge1100__PMID33875891
rfMRIconnectivity_ICA100edge1284__PMID33875891
rfMRIconnectivity_ICA100edge69__PMID33875891
Types_of_transport_used_excluding_work_Public_transport_PMID31427789
rfMRIconnectivity_ICA100edge1120__PMID33875891
rfMRIconnectivity_ICA100edge1028__PMID33875891
rfMRIconnectivity_ICA100edge33__PMID33875891
rfMRIconnectivity_ICA100edge444__PMID33875891
Suicide_attempt_Model_3_PMID30116032
rfMRIconnectivity_ICA100edge516__PMID33875891
rfMRIconnectivity_ICA100edge607__PMID33875891
rfMRIconnectivity_ICA100edge754__PMID33875891
rfMRIconnectivity_ICA100edge327__PMID33875891
rfMRIconnectivity_ICA100edge1349__PMID33875891
rfMRIconnectivity_ICA100edge986__PMID33875891
rfMRIconnectivity_ICA100edge350__PMID33875891
rfMRIconnectivity_ICA100edge399__PMID33875891
rfMRIconnectivity_ICA100edge161__PMID33875891
rfMRIconnectivity_ICA100edge970__PMID33875891
rfMRIconnectivity_ICA100edge1147__PMID33875891
rfMRIconnectivity_ICA100edge619__PMID33875891
Numeric_memory_test_Maximum_digits_remembered_correctly_PMID31427789
rfMRIconnectivity_ICA100edge873__PMID33875891

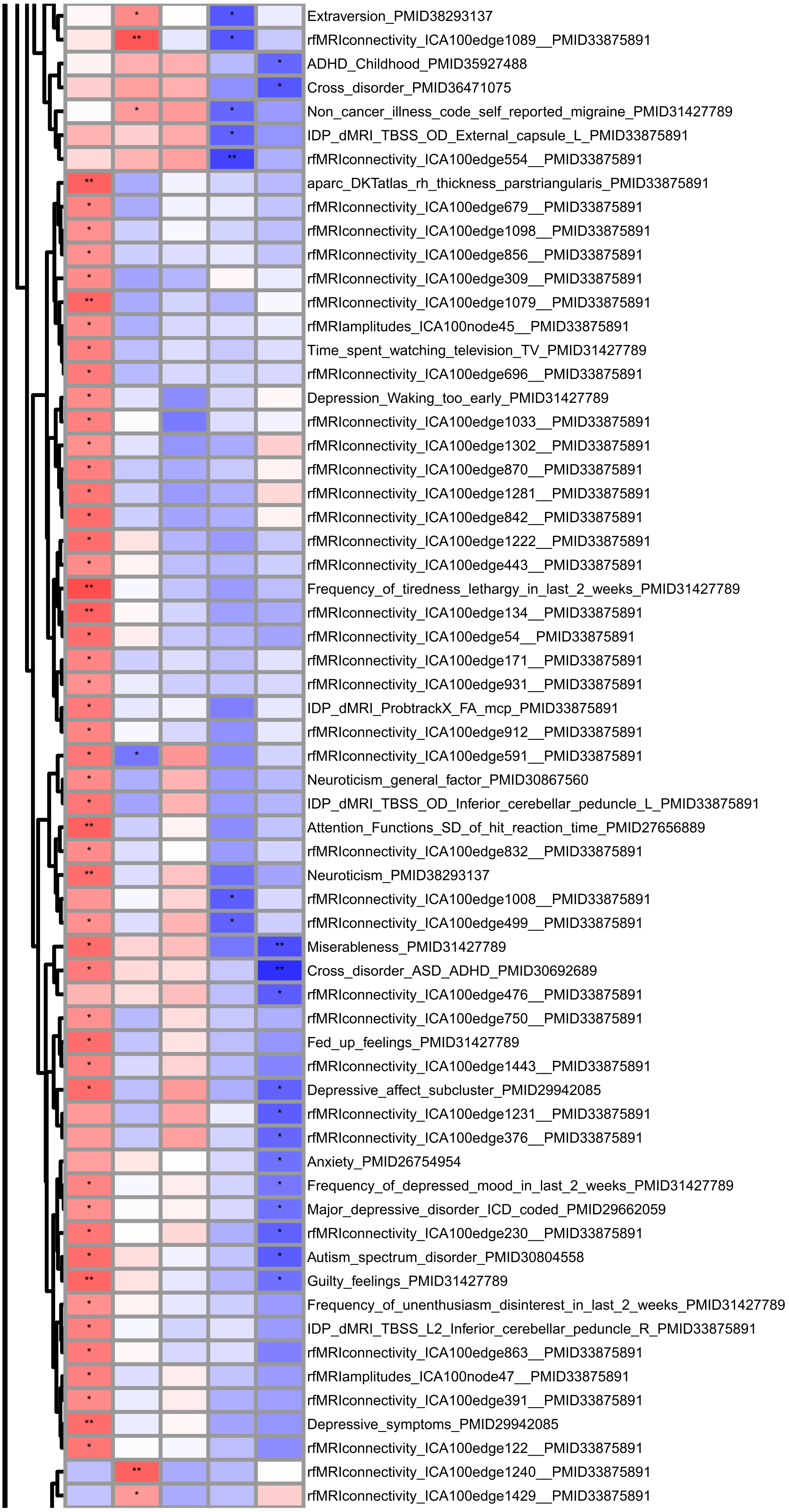
Extraversion_PMID38293137
rfMRIconnectivity_ICA100edge1089__PMID33875891
ADHD_Childhood_PMID35927488
Cross_disorder_PMID36471075
Non_cancer_illness_code_self_reported_migraine_PMID31427789
IDP_dMRI_TBSS_OD_External_capsule_L_PMID33875891
rfMRIconnectivity_ICA100edge554__PMID33875891
aparc_DKTatlas_rh_thickness_parstriangularis_PMID33875891
rfMRIconnectivity_ICA100edge679__PMID33875891
rfMRIconnectivity_ICA100edge1098__PMID33875891
rfMRIconnectivity_ICA100edge856__PMID33875891
rfMRIconnectivity_ICA100edge309__PMID33875891
rfMRIconnectivity_ICA100edge1079__PMID33875891
rfMRIamplitudes_ICA100node45__PMID33875891
Time_spent_watching_television_TV_PMID31427789
rfMRIconnectivity_ICA100edge696__PMID33875891
Depression_Waking_too_early_PMID31427789
rfMRIconnectivity_ICA100edge1033__PMID33875891
rfMRIconnectivity_ICA100edge1302__PMID33875891
rfMRIconnectivity_ICA100edge870__PMID33875891
rfMRIconnectivity_ICA100edge1281__PMID33875891
rfMRIconnectivity_ICA100edge842__PMID33875891
rfMRIconnectivity_ICA100edge1222__PMID33875891
rfMRIconnectivity_ICA100edge443__PMID33875891
Frequency_of_tiredness_lethargy_in_last_2_weeks_PMID31427789
rfMRIconnectivity_ICA100edge134__PMID33875891
rfMRIconnectivity_ICA100edge54__PMID33875891
rfMRIconnectivity_ICA100edge171__PMID33875891
rfMRIconnectivity_ICA100edge931__PMID33875891
IDP_dMRI_ProbtrackX_FA_mcp_PMID33875891
rfMRIconnectivity_ICA100edge912__PMID33875891
rfMRIconnectivity_ICA100edge591__PMID33875891
Neuroticism_general_factor_PMID30867560
IDP_dMRI_TBSS_OD_Inferior_cerebellar_peduncle_L_PMID33875891
Attention_Functions_SD_of_hit_reaction_time_PMID27656889
rfMRIconnectivity_ICA100edge832__PMID33875891
Neuroticism_PMID38293137
rfMRIconnectivity_ICA100edge1008__PMID33875891
rfMRIconnectivity_ICA100edge499__PMID33875891
Miserableness_PMID31427789
Cross_disorder_ASD_ADHD_PMID30692689
rfMRIconnectivity_ICA100edge476__PMID33875891
rfMRIconnectivity_ICA100edge750__PMID33875891
Fed_up_feelings_PMID31427789
rfMRIconnectivity_ICA100edge1443__PMID33875891
Depressive_affect_subcluster_PMID29942085
rfMRIconnectivity_ICA100edge1231__PMID33875891
rfMRIconnectivity_ICA100edge376__PMID33875891
Anxiety_PMID26754954
Frequency_of_depressed_mood_in_last_2_weeks_PMID31427789
Major_depressive_disorder_ICD_coded_PMID29662059
rfMRIconnectivity_ICA100edge230__PMID33875891
Autism_spectrum_disorder_PMID30804558
Guilty_feelings_PMID31427789
Frequency_of_unenthusiasm_disinterest_in_last_2_weeks_PMID31427789
IDP_dMRI_TBSS_L2_Inferior_cerebellar_peduncle_R_PMID33875891
rfMRIconnectivity_ICA100edge863__PMID33875891
rfMRIamplitudes_ICA100node47__PMID33875891
rfMRIconnectivity_ICA100edge391__PMID33875891
Depressive_symptoms_PMID29942085
rfMRIconnectivity_ICA100edge122__PMID33875891
rfMRIconnectivity_ICA100edge1240__PMID33875891
rfMRIconnectivity_ICA100edge1429__PMID33875891

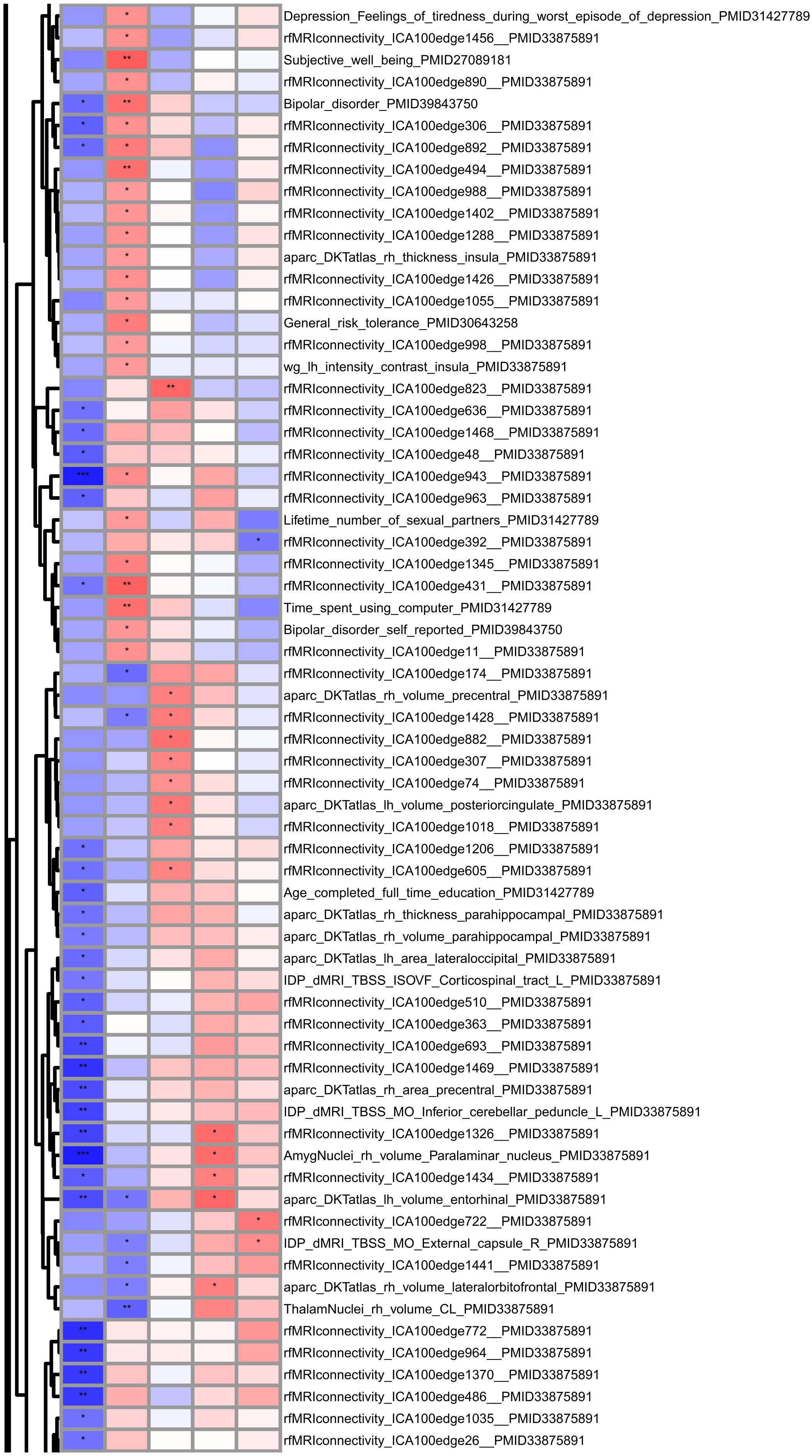
Depression_Feelings_of_tiredness_during_worst_episode_of_depression_PMID31427789
rfMRIconnectivity_ICA100edge1456__PMID33875891
Subjective_well_being_PMID27089181
rfMRIconnectivity_ICA100edge890__PMID33875891
Bipolar_disorder_PMID39843750
rfMRIconnectivity_ICA100edge306__PMID33875891
rfMRIconnectivity_ICA100edge892__PMID33875891
rfMRIconnectivity_ICA100edge494__PMID33875891
rfMRIconnectivity_ICA100edge988__PMID33875891
rfMRIconnectivity_ICA100edge1402__PMID33875891
rfMRIconnectivity_ICA100edge1288__PMID33875891
aparc_DKTatlas_rh_thickness_insula_PMID33875891
rfMRIconnectivity_ICA100edge1426__PMID33875891
rfMRIconnectivity_ICA100edge1055__PMID33875891
General_risk_tolerance_PMID30643258
rfMRIconnectivity_ICA100edge998__PMID33875891
wg_lh_intensity_contrast_insula_PMID33875891
rfMRIconnectivity_ICA100edge823__PMID33875891
rfMRIconnectivity_ICA100edge636__PMID33875891
rfMRIconnectivity_ICA100edge1468__PMID33875891
rfMRIconnectivity_ICA100edge48__PMID33875891
rfMRIconnectivity_ICA100edge943__PMID33875891
rfMRIconnectivity_ICA100edge963__PMID33875891
Lifetime_number_of_sexual_partners_PMID31427789
rfMRIconnectivity_ICA100edge392__PMID33875891
rfMRIconnectivity_ICA100edge1345__PMID33875891
rfMRIconnectivity_ICA100edge431__PMID33875891
Time_spent_using_computer_PMID31427789
Bipolar_disorder_self_reported_PMID39843750
rfMRIconnectivity_ICA100edge11__PMID33875891
rfMRIconnectivity_ICA100edge174__PMID33875891
aparc_DKTatlas_rh_volume_precentral_PMID33875891
rfMRIconnectivity_ICA100edge1428__PMID33875891
rfMRIconnectivity_ICA100edge882__PMID33875891
rfMRIconnectivity_ICA100edge307__PMID33875891
rfMRIconnectivity_ICA100edge74__PMID33875891
aparc_DKTatlas_lh_volume_posteriorcingulate_PMID33875891
rfMRIconnectivity_ICA100edge1018__PMID33875891
rfMRIconnectivity_ICA100edge1206__PMID33875891
rfMRIconnectivity_ICA100edge605__PMID33875891
Age_completed_full_time_education_PMID31427789
aparc_DKTatlas_rh_thickness_parahippocampal_PMID33875891
aparc_DKTatlas_rh_volume_parahippocampal_PMID33875891
aparc_DKTatlas_lh_area_lateraloccipital_PMID33875891
IDP_dMRI_TBSS_ISOVF_Corticospinal_tract_L_PMID33875891
rfMRIconnectivity_ICA100edge510__PMID33875891
rfMRIconnectivity_ICA100edge363__PMID33875891
rfMRIconnectivity_ICA100edge693__PMID33875891
rfMRIconnectivity_ICA100edge1469__PMID33875891
aparc_DKTatlas_rh_area_precentral_PMID33875891
IDP_dMRI_TBSS_MO_Inferior_cerebellar_peduncle_L_PMID33875891
rfMRIconnectivity_ICA100edge1326__PMID33875891
AmygNuclei_rh_volume_Paralaminar_nucleus_PMID33875891
rfMRIconnectivity_ICA100edge1434__PMID33875891
aparc_DKTatlas_lh_volume_entorhinal_PMID33875891
rfMRIconnectivity_ICA100edge722__PMID33875891
IDP_dMRI_TBSS_MO_External_capsule_R_PMID33875891
rfMRIconnectivity_ICA100edge1441__PMID33875891
aparc_DKTatlas_rh_volume_lateralorbitofrontal_PMID33875891
ThalamNuclei_rh_volume_CL_PMID33875891
rfMRIconnectivity_ICA100edge772__PMID33875891
rfMRIconnectivity_ICA100edge964__PMID33875891
rfMRIconnectivity_ICA100edge1370__PMID33875891
rfMRIconnectivity_ICA100edge486__PMID33875891
rfMRIconnectivity_ICA100edge1035__PMID33875891
rfMRIconnectivity_ICA100edge26__PMID33875891

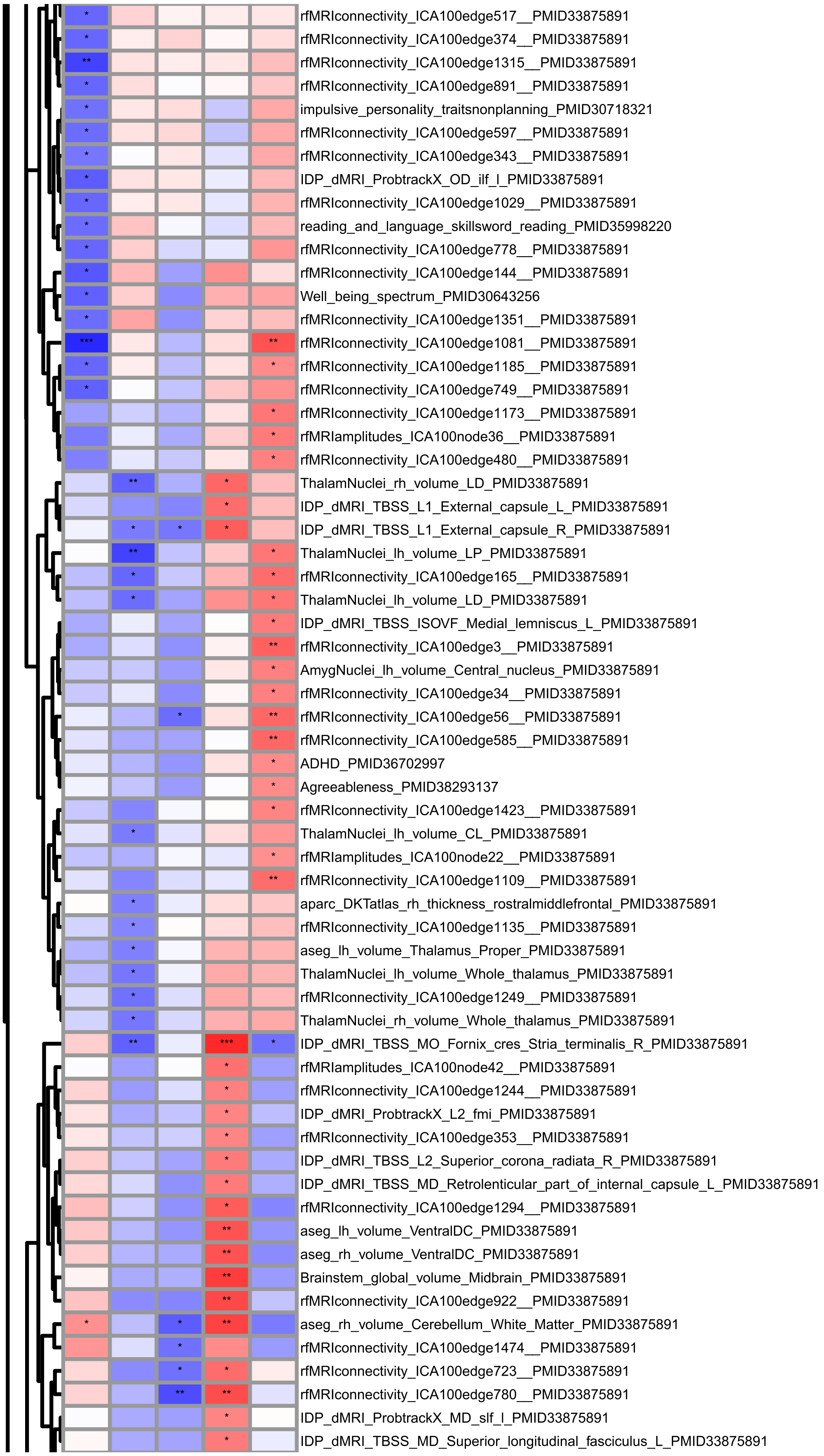
rfMRIconnectivity_ICA100edge517__PMID33875891
rfMRIconnectivity_ICA100edge374__PMID33875891
rfMRIconnectivity_ICA100edge1315__PMID33875891
rfMRIconnectivity_ICA100edge891__PMID33875891
impulsive_personality_traitsnonplanning_PMID30718321
rfMRIconnectivity_ICA100edge597__PMID33875891
rfMRIconnectivity_ICA100edge343__PMID33875891
IDP_dMRI_ProbtrackX_OD_ilf_l_PMID33875891
rfMRIconnectivity_ICA100edge1029__PMID33875891
reading_and_language_skillsword_reading_PMID35998220
rfMRIconnectivity_ICA100edge778__PMID33875891
rfMRIconnectivity_ICA100edge144__PMID33875891
Well_being_spectrum_PMID30643256
rfMRIconnectivity_ICA100edge1351__PMID33875891
rfMRIconnectivity_ICA100edge1081__PMID33875891
rfMRIconnectivity_ICA100edge1185__PMID33875891
rfMRIconnectivity_ICA100edge749__PMID33875891
rfMRIconnectivity_ICA100edge1173__PMID33875891
rfMRIamplitudes_ICA100node36__PMID33875891
rfMRIconnectivity_ICA100edge480__PMID33875891
ThalamNuclei_rh_volume_LD_PMID33875891
IDP_dMRI_TBSS_L1_External_capsule_L_PMID33875891
IDP_dMRI_TBSS_L1_External_capsule_R_PMID33875891
ThalamNuclei_lh_volume_LP_PMID33875891
rfMRIconnectivity_ICA100edge165__PMID33875891
ThalamNuclei_lh_volume_LD_PMID33875891
IDP_dMRI_TBSS_ISOVF_Medial_lemniscus_L_PMID33875891
rfMRIconnectivity_ICA100edge3__PMID33875891
AmygNuclei_lh_volume_Central_nucleus_PMID33875891
rfMRIconnectivity_ICA100edge34__PMID33875891
rfMRIconnectivity_ICA100edge56__PMID33875891
rfMRIconnectivity_ICA100edge585__PMID33875891
ADHD_PMID36702997
Agreeableness_PMID38293137
rfMRIconnectivity_ICA100edge1423__PMID33875891
ThalamNuclei_lh_volume_CL_PMID33875891
rfMRIamplitudes_ICA100node22__PMID33875891
rfMRIconnectivity_ICA100edge1109__PMID33875891
aparc_DKTatlas_rh_thickness_rostralmiddlefrontal_PMID33875891
rfMRIconnectivity_ICA100edge1135__PMID33875891
aseg_lh_volume_Thalamus_Proper_PMID33875891
ThalamNuclei_lh_volume_Whole_thalamus_PMID33875891
rfMRIconnectivity_ICA100edge1249__PMID33875891
ThalamNuclei_rh_volume_Whole_thalamus_PMID33875891
IDP_dMRI_TBSS_MO_Fornix_cres_Stria_terminalis_R_PMID33875891
rfMRIamplitudes_ICA100node42__PMID33875891
rfMRIconnectivity_ICA100edge1244__PMID33875891
IDP_dMRI_ProbtrackX_L2_fmi_PMID33875891
rfMRIconnectivity_ICA100edge353__PMID33875891
IDP_dMRI_TBSS_L2_Superior_corona_radiata_R_PMID33875891
IDP_dMRI_TBSS_MD_Retrolenticular_part_of_internal_capsule_L_PMID33875891
rfMRIconnectivity_ICA100edge1294__PMID33875891
aseg_lh_volume_VentralDC_PMID33875891
aseg_rh_volume_VentralDC_PMID33875891
Brainstem_global_volume_Midbrain_PMID33875891
rfMRIconnectivity_ICA100edge922__PMID33875891
aseg_rh_volume_Cerebellum_White_Matter_PMID33875891
rfMRIconnectivity_ICA100edge1474__PMID33875891
rfMRIconnectivity_ICA100edge723__PMID33875891
rfMRIconnectivity_ICA100edge780__PMID33875891
IDP_dMRI_ProbtrackX_MD_slf_l_PMID33875891
IDP_dMRI_TBSS_MD_Superior_longitudinal_fasciculus_L_PMID33875891

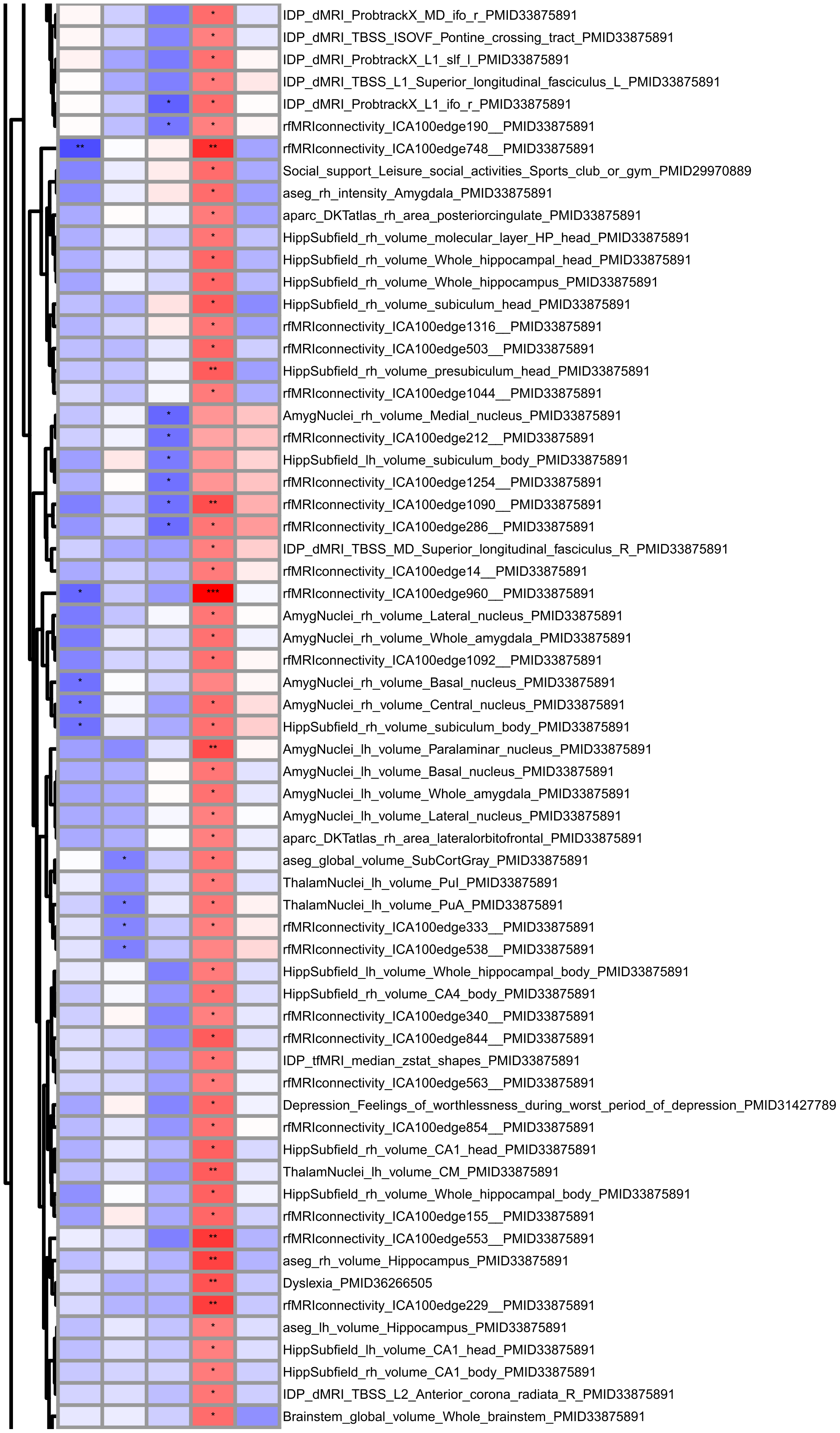
IDP_dMRI_ProbtrackX_MD_ifo_r_PMID33875891
IDP_dMRI_TBSS_ISOVF_Pontine_crossing_tract_PMID33875891
IDP_dMRI_ProbtrackX_L1_slf_l_PMID33875891
IDP_dMRI_TBSS_L1_Superior_longitudinal_fasciculus_L_PMID33875891
IDP_dMRI_ProbtrackX_L1_ifo_r_PMID33875891
rfMRIconnectivity_ICA100edge190__PMID33875891
rfMRIconnectivity_ICA100edge748__PMID33875891
Social_support_Leisure_social_activities_Sports_club_or_gym_PMID29970889
aseg_rh_intensity_Amygdala_PMID33875891
aparc_DKTatlas_rh_area_posteriorcingulate_PMID33875891
HippSubfield_rh_volume_molecular_layer_HP_head_PMID33875891
HippSubfield_rh_volume_Whole_hippocampal_head_PMID33875891
HippSubfield_rh_volume_Whole_hippocampus_PMID33875891
HippSubfield_rh_volume_subiculum_head_PMID33875891
rfMRIconnectivity_ICA100edge1316__PMID33875891
rfMRIconnectivity_ICA100edge503__PMID33875891
HippSubfield_rh_volume_presubiculum_head_PMID33875891
rfMRIconnectivity_ICA100edge1044__PMID33875891
AmygNuclei_rh_volume_Medial_nucleus_PMID33875891
rfMRIconnectivity_ICA100edge212__PMID33875891
HippSubfield_lh_volume_subiculum_body_PMID33875891
rfMRIconnectivity_ICA100edge1254__PMID33875891
rfMRIconnectivity_ICA100edge1090__PMID33875891
rfMRIconnectivity_ICA100edge286__PMID33875891
IDP_dMRI_TBSS_MD_Superior_longitudinal_fasciculus_R_PMID33875891
rfMRIconnectivity_ICA100edge14__PMID33875891
rfMRIconnectivity_ICA100edge960__PMID33875891
AmygNuclei_rh_volume_Lateral_nucleus_PMID33875891
AmygNuclei_rh_volume_Whole_amygdala_PMID33875891
rfMRIconnectivity_ICA100edge1092__PMID33875891
AmygNuclei_rh_volume_Basal_nucleus_PMID33875891
AmygNuclei_rh_volume_Central_nucleus_PMID33875891
HippSubfield_rh_volume_subiculum_body_PMID33875891
AmygNuclei_lh_volume_Paralaminar_nucleus_PMID33875891
AmygNuclei_lh_volume_Basal_nucleus_PMID33875891
AmygNuclei_lh_volume_Whole_amygdala_PMID33875891
AmygNuclei_lh_volume_Lateral_nucleus_PMID33875891
aparc_DKTatlas_rh_area_lateralorbitofrontal_PMID33875891
aseg_global_volume_SubCortGray_PMID33875891
ThalamNuclei_lh_volume_PuI_PMID33875891
ThalamNuclei_lh_volume_PuA_PMID33875891
rfMRIconnectivity_ICA100edge333__PMID33875891
rfMRIconnectivity_ICA100edge538__PMID33875891
HippSubfield_lh_volume_Whole_hippocampal_body_PMID33875891
HippSubfield_rh_volume_CA4_body_PMID33875891
rfMRIconnectivity_ICA100edge340__PMID33875891
rfMRIconnectivity_ICA100edge844__PMID33875891
IDP_tfMRI_median_zstat_shapes_PMID33875891
rfMRIconnectivity_ICA100edge563__PMID33875891
Depression_Feelings_of_worthlessness_during_worst_period_of_depression_PMID31427789
rfMRIconnectivity_ICA100edge854__PMID33875891
HippSubfield_rh_volume_CA1_head_PMID33875891
ThalamNuclei_lh_volume_CM_PMID33875891
HippSubfield_rh_volume_Whole_hippocampal_body_PMID33875891
rfMRIconnectivity_ICA100edge155__PMID33875891
rfMRIconnectivity_ICA100edge553__PMID33875891
aseg_rh_volume_Hippocampus_PMID33875891
Dyslexia_PMID36266505
rfMRIconnectivity_ICA100edge229__PMID33875891
aseg_lh_volume_Hippocampus_PMID33875891
HippSubfield_lh_volume_CA1_head_PMID33875891
HippSubfield_rh_volume_CA1_body_PMID33875891
IDP_dMRI_TBSS_L2_Anterior_corona_radiata_R_PMID33875891
Brainstem_global_volume_Whole_brainstem_PMID33875891

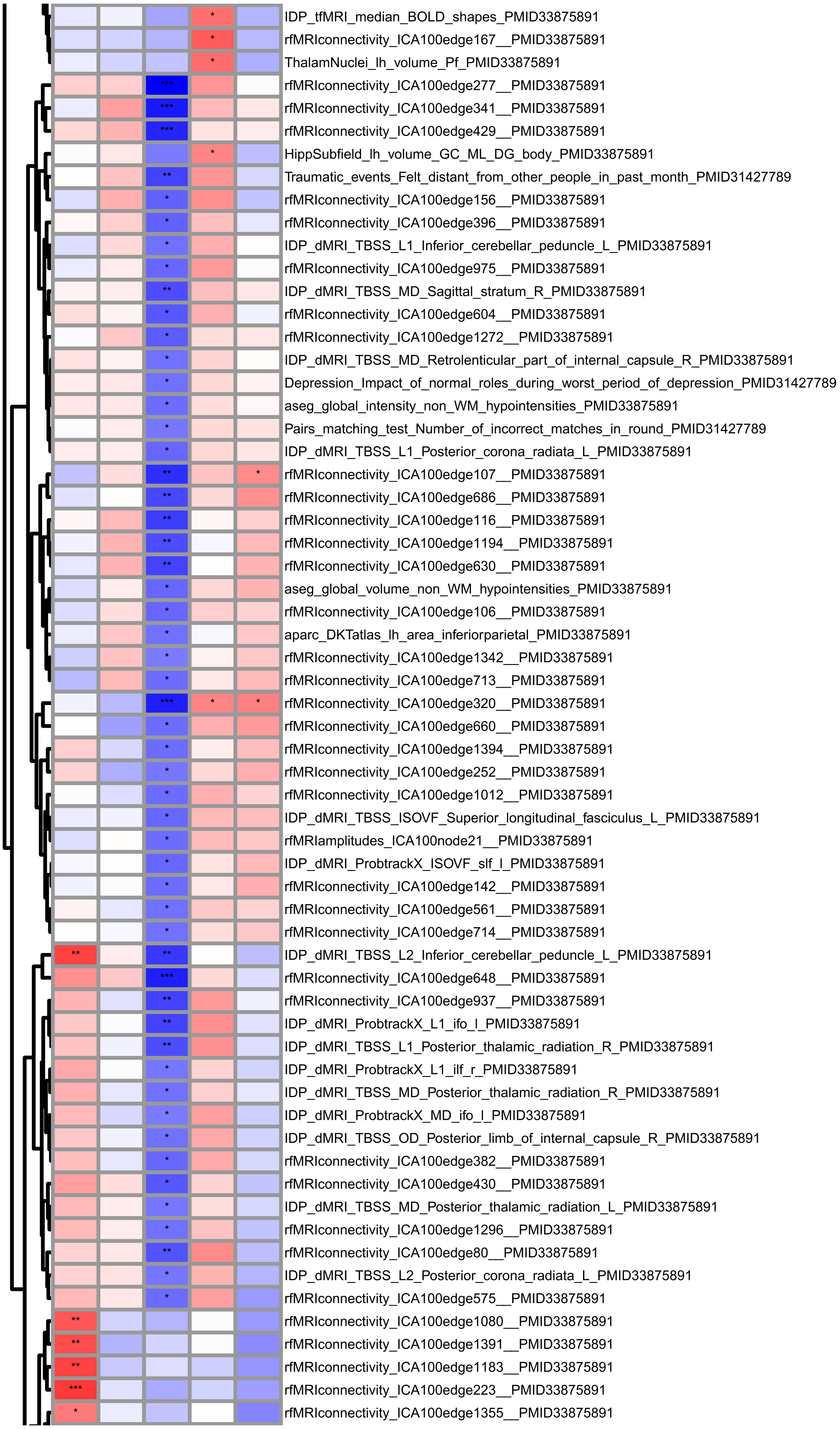
IDP_tfMRI_median_BOLD_shapes_PMID33875891
rfMRIconnectivity_ICA100edge167__PMID33875891
ThalamNuclei_lh_volume_Pf_PMID33875891
rfMRIconnectivity_ICA100edge277__PMID33875891
rfMRIconnectivity_ICA100edge341__PMID33875891
rfMRIconnectivity_ICA100edge429__PMID33875891
HippSubfield_lh_volume_GC_ML_DG_body_PMID33875891
Traumatic_events_Felt_distant_from_other_people_in_past_month_PMID31427789
rfMRIconnectivity_ICA100edge156__PMID33875891
rfMRIconnectivity_ICA100edge396__PMID33875891
IDP_dMRI_TBSS_L1_Inferior_cerebellar_peduncle_L_PMID33875891
rfMRIconnectivity_ICA100edge975__PMID33875891
IDP_dMRI_TBSS_MD_Sagittal_stratum_R_PMID33875891
rfMRIconnectivity_ICA100edge604__PMID33875891
rfMRIconnectivity_ICA100edge1272__PMID33875891
IDP_dMRI_TBSS_MD_Retrolenticular_part_of_internal_capsule_R_PMID33875891
Depression_Impact_of_normal_roles_during_worst_period_of_depression_PMID31427789
aseg_global_intensity_non_WM_hypointensities_PMID33875891
Pairs_matching_test_Number_of_incorrect_matches_in_round_PMID31427789
IDP_dMRI_TBSS_L1_Posterior_corona_radiata_L_PMID33875891
rfMRIconnectivity_ICA100edge107__PMID33875891
rfMRIconnectivity_ICA100edge686__PMID33875891
rfMRIconnectivity_ICA100edge116__PMID33875891
rfMRIconnectivity_ICA100edge1194__PMID33875891
rfMRIconnectivity_ICA100edge630__PMID33875891
aseg_global_volume_non_WM_hypointensities_PMID33875891
rfMRIconnectivity_ICA100edge106__PMID33875891
aparc_DKTatlas_lh_area_inferiorparietal_PMID33875891
rfMRIconnectivity_ICA100edge1342__PMID33875891
rfMRIconnectivity_ICA100edge713__PMID33875891
rfMRIconnectivity_ICA100edge320__PMID33875891
rfMRIconnectivity_ICA100edge660__PMID33875891
rfMRIconnectivity_ICA100edge1394__PMID33875891
rfMRIconnectivity_ICA100edge252__PMID33875891
rfMRIconnectivity_ICA100edge1012__PMID33875891
IDP_dMRI_TBSS_ISOVF_Superior_longitudinal_fasciculus_L_PMID33875891
rfMRIamplitudes_ICA100node21__PMID33875891
IDP_dMRI_ProbtrackX_ISOVF_slf_l_PMID33875891
rfMRIconnectivity_ICA100edge142__PMID33875891
rfMRIconnectivity_ICA100edge561__PMID33875891
rfMRIconnectivity_ICA100edge714__PMID33875891
IDP_dMRI_TBSS_L2_Inferior_cerebellar_peduncle_L_PMID33875891
rfMRIconnectivity_ICA100edge648__PMID33875891
rfMRIconnectivity_ICA100edge937__PMID33875891
IDP_dMRI_ProbtrackX_L1_ifo_l_PMID33875891
IDP_dMRI_TBSS_L1_Posterior_thalamic_radiation_R_PMID33875891
IDP_dMRI_ProbtrackX_L1_ilf_r_PMID33875891
IDP_dMRI_TBSS_MD_Posterior_thalamic_radiation_R_PMID33875891
IDP_dMRI_ProbtrackX_MD_ifo_l_PMID33875891
IDP_dMRI_TBSS_OD_Posterior_limb_of_internal_capsule_R_PMID33875891
rfMRIconnectivity_ICA100edge382__PMID33875891
rfMRIconnectivity_ICA100edge430__PMID33875891
IDP_dMRI_TBSS_MD_Posterior_thalamic_radiation_L_PMID33875891
rfMRIconnectivity_ICA100edge1296__PMID33875891
rfMRIconnectivity_ICA100edge80__PMID33875891
IDP_dMRI_TBSS_L2_Posterior_corona_radiata_L_PMID33875891
rfMRIconnectivity_ICA100edge575__PMID33875891
rfMRIconnectivity_ICA100edge1080__PMID33875891
rfMRIconnectivity_ICA100edge1391__PMID33875891
rfMRIconnectivity_ICA100edge1183__PMID33875891
rfMRIconnectivity_ICA100edge223__PMID33875891
rfMRIconnectivity_ICA100edge1355__PMID33875891

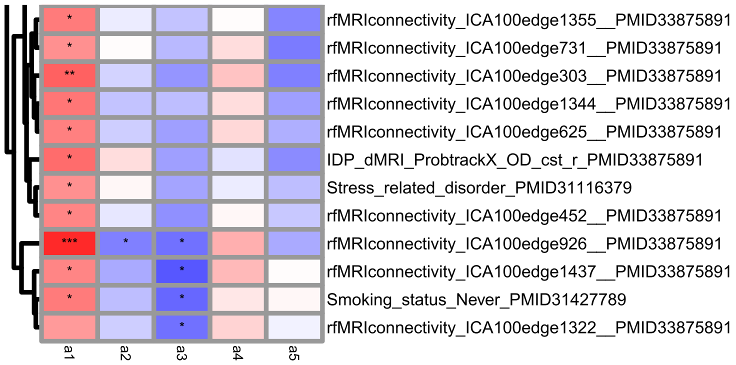

**Figure S9 |** Heatmap of linear regression coefficients for polygenic scores (PGSs) across archetypes scores. The x-axis shows each archetype score, and the y-axis lists PGSs with nominally significant associations ($p < 0.05$). Colors represent standardized regression coefficients: red for positive associations and blue for negative associations. Significant associations are marked with "*".

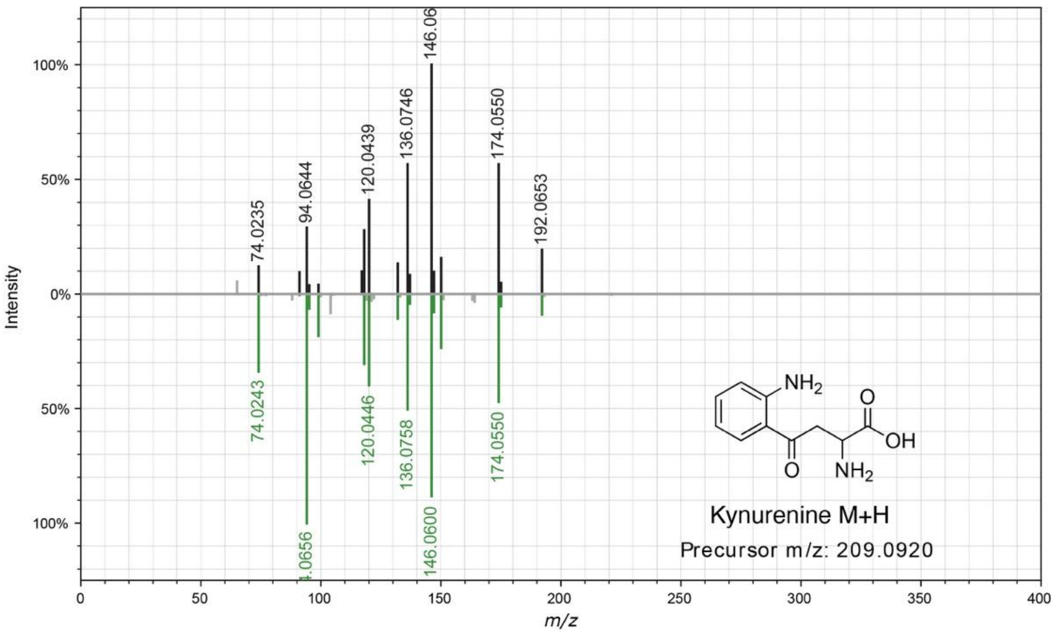

**Figure S10 |** Spectral mirror plot of kynurenine (Feature ID 1801) retrieved from GNPS. Spectral similarity scores (cosine score) are shown in Table S11. Data were extracted using the metabolomics USI tool (http://metabolomics-usi.ucsd.edu/).

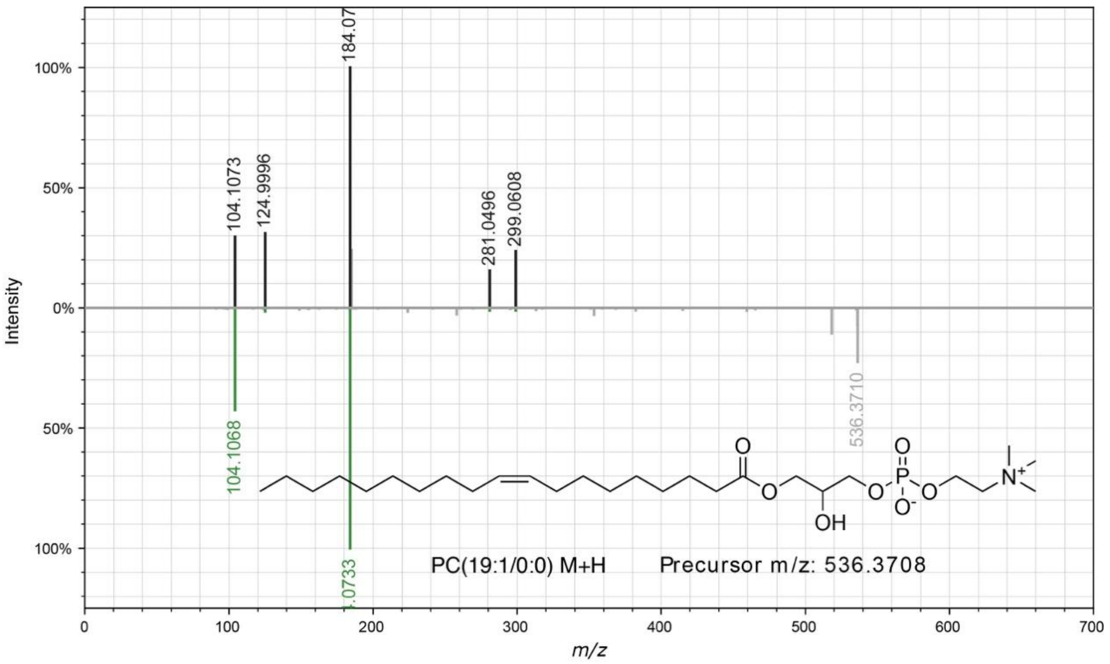

**Figure S11 |** Spectral mirror plot of PC(19:1/0:0) (Feature ID 8287) retrieved from GNPS. Spectral similarity scores (cosine score) are shown in Table S11. Data were extracted using the metabolomics USI tool (http://metabolomics-usi.ucsd.edu/).

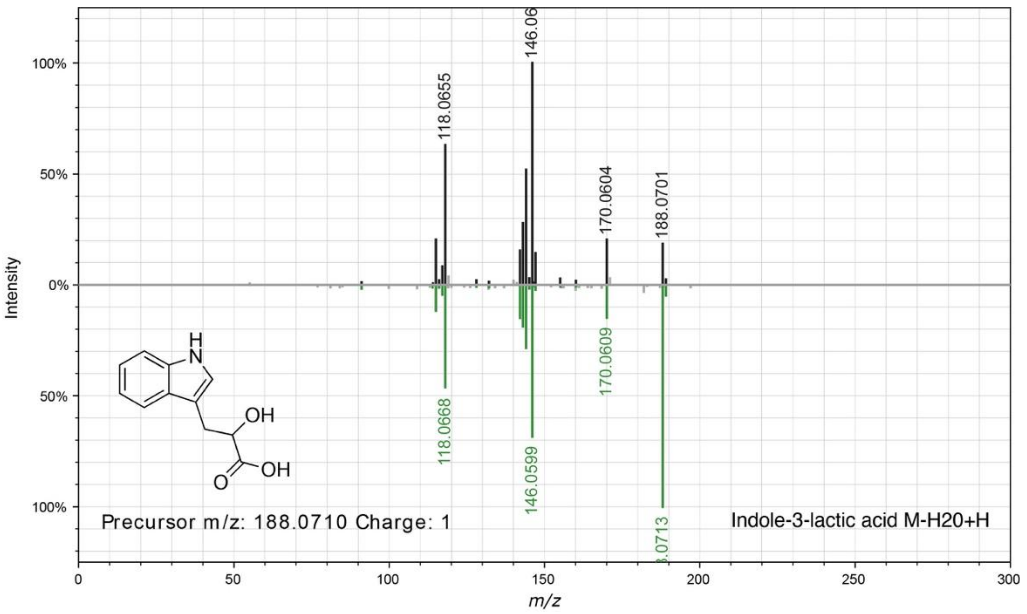

**Figure S12 |** Spectral mirror plot of indole-3-lactic acid (Feature ID 2162) retrieved from GNPS. Spectral similarity scores (cosine score) are shown in Table S11. Data were extracted using the metabolomics USI tool (http://metabolomics-usi.ucsd.edu/).

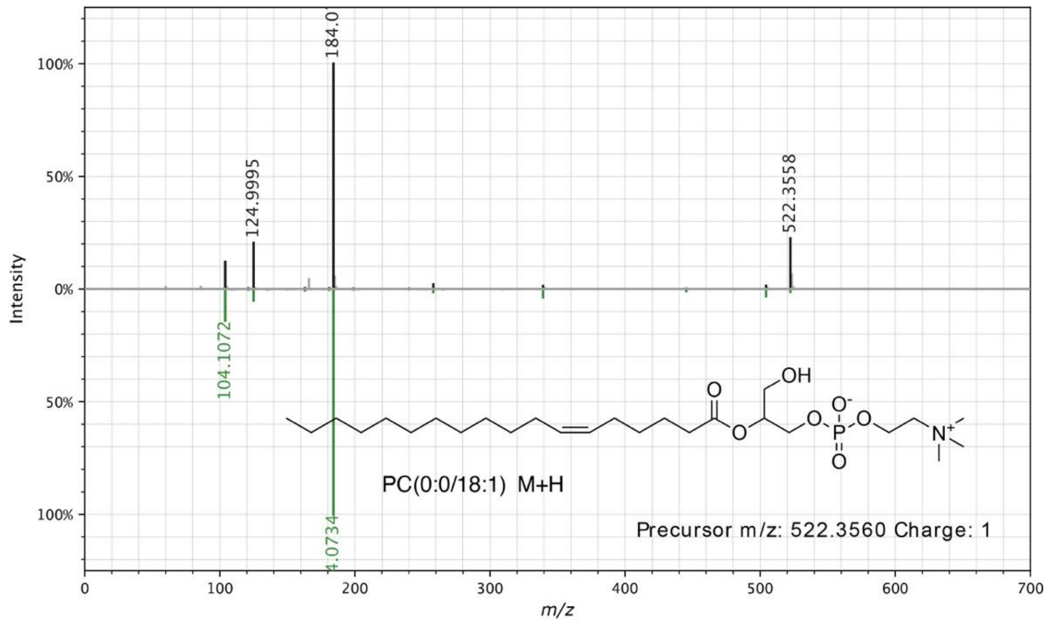

**Figure S13 |** Spectral mirror plot of PC(0:0/18:1) (Feature ID 7451) retrieved from GNPS. Spectral similarity scores (cosine score) are shown in Table S11. Data were extracted using the metabolomics USI tool (http://metabolomics-usi.ucsd.edu/).

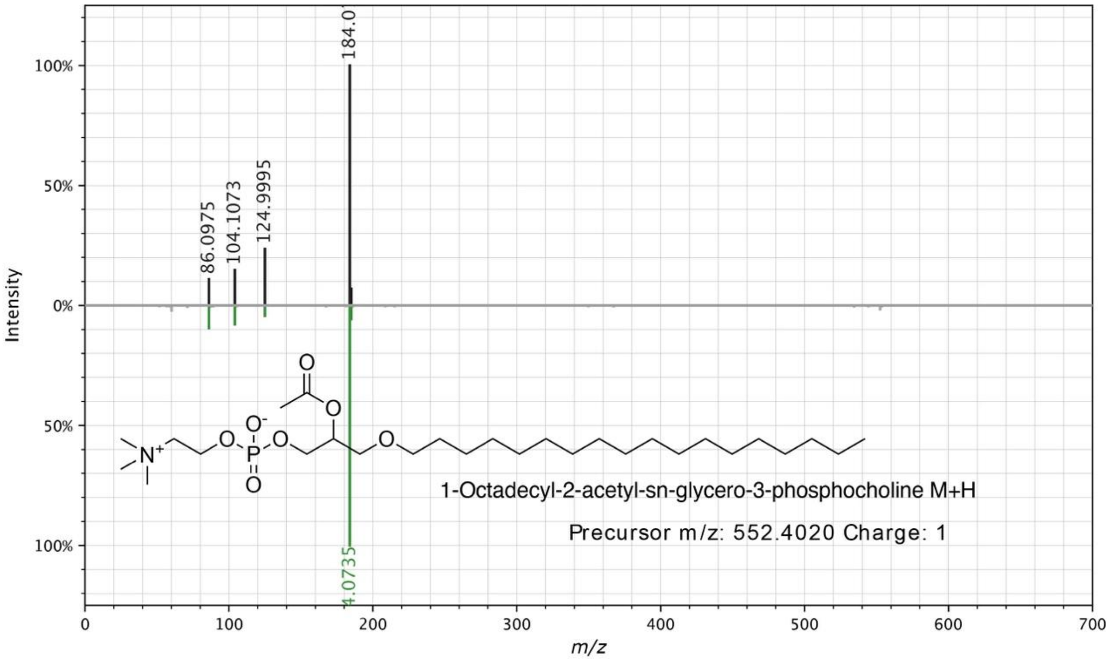

**Figure S14 |** Spectral mirror plot of 1-octadecyl-2-acetyl-sn-glycero-3-phosphocholine (Feature ID 10368) retrieved from GNPS. Spectral similarity scores (cosine score) are shown in Table S11. Data were extracted using the metabolomics USI tool (http://metabolomics-usi.ucsd.edu/).

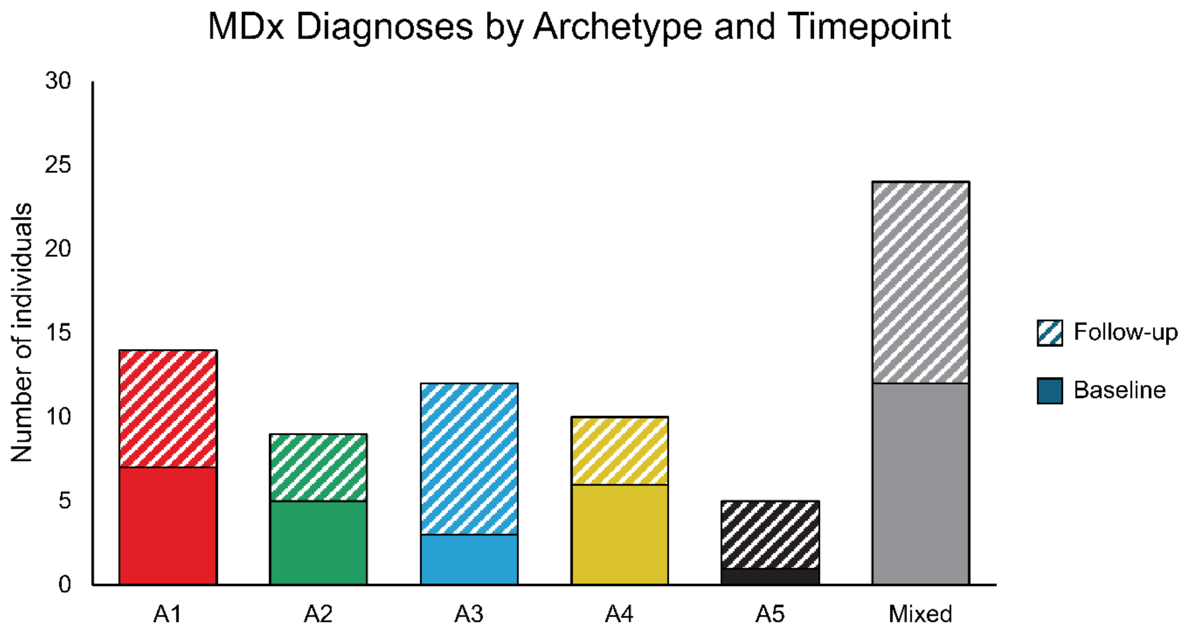

**Figure S15 |** MDx diagnoses count at baseline and follow-up in each discrete archetype. The x-axis shows each discrete archetype and the mixed group, whereas the y-axis shows the number of individuals with a self-reported MDx diagnosis. Solid colors: responses at baseline; striped colors: responses at follow-up.

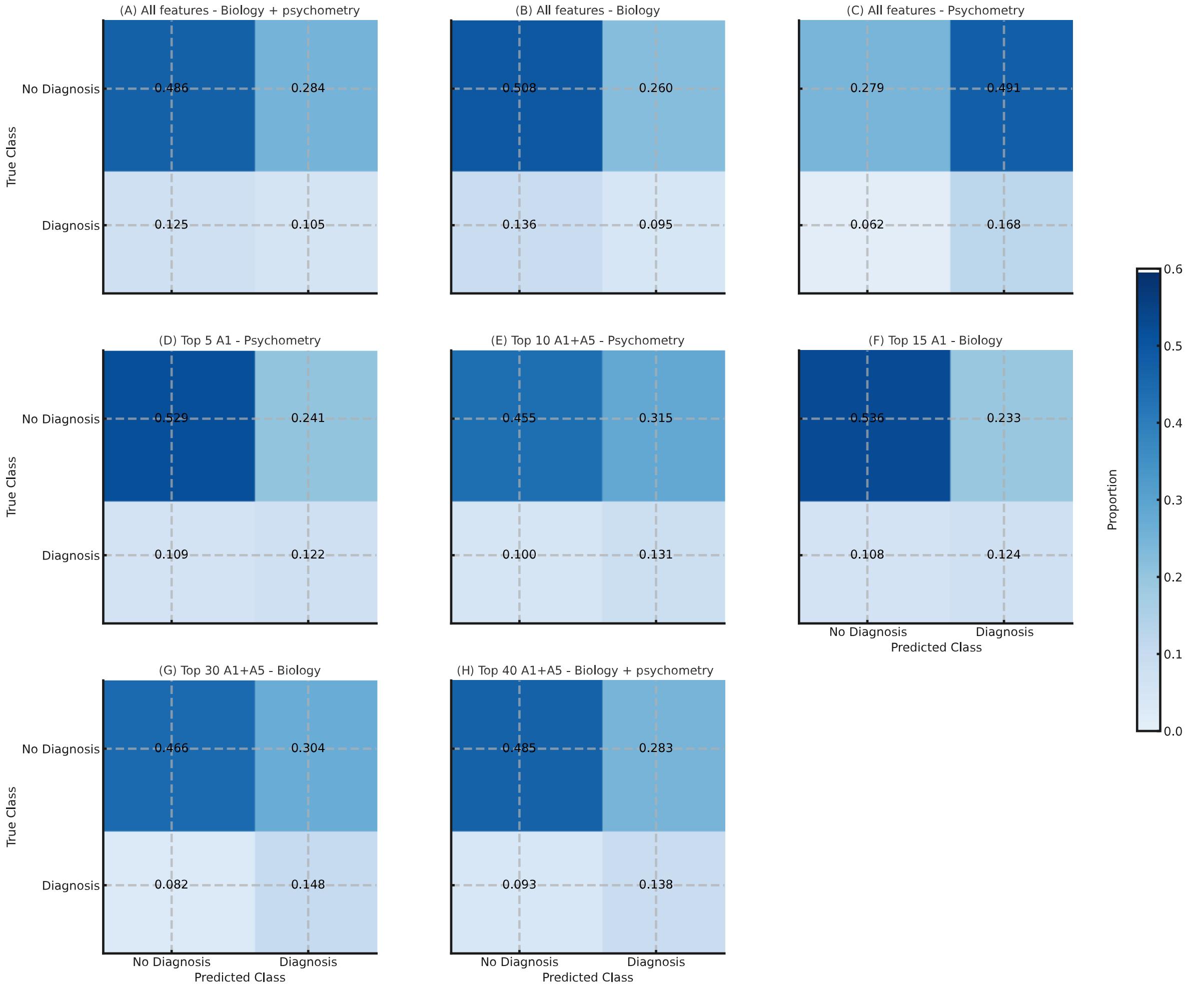

**Figure S16 |** Confusion matrices across feature sets.

Confusion matrices show predicted vs. true class proportions for eight feature configurations. Rows indicate true class ("No Diagnosis", "Diagnosis"); columns show predicted class. Cell values represent the proportion of the test set in each outcome. Panels correspond to: (**A**) All features - Biology + Psychometry; (**B**) All features - Biology; (**C**) All features - Psychometry; (**D**) Top 5 A1 score – Psychometry; (**E**) Top 10 A1+A5 scores – Psychometry; (**F**) Top 15 A1 score - Biology; (**G**) Top 30 A1+A5 scores - Biology; (**H**) Top 40 A1+A5 scores – Biology + psychometry
